# Control, Fludrocortisone or Midodrine for the treatment of Orthostatic Hypotension (CONFORM-OH): Results from an internal pilot randomised controlled trial

**DOI:** 10.1101/2025.06.06.25329129

**Authors:** Helen Mossop, Sarah Al-Ashmori, Tumi Sotire, Emma Clark, Gillian Watson, Miles D Witham, Luke Vale, Naomi McGregor, Julia Phillipson, James M. S. Wason, Alison J. Yarnall, Steve Parry, Helen Hancock, Rose Anne Kenny, James Frith

**Affiliations:** Population Health Sciences Institute, Newcastle University, Newcastle-Upon-Tyne, UK; Health Economics Group, Population Health Sciences Institute, Newcastle University, Newcastle-Upon-Tyne, UK; Newcastle Clinical Trials Unit, Newcastle University, Newcastle-Upon-Tyne, UK; AGE Research Group, Translational and Clinical Research Institute, Faculty of Medical Sciences, Newcastle University, Newcastle, UK; NIHR Newcastle Biomedical Research Centre, Newcastle upon Tyne Hospitals NHS Foundation Trust, Cumbria, Northumberland, Tyne and Wear NHS Foundation Trust and Newcastle University, Newcastle, UK; Translational and Clinical Research Institute, Newcastle University and Newcastle upon Tyne NHS Foundation Trust; The Newcastle upon Tyne Hospitals NHS Trust, Newcastle upon Tyne, UK; Trinity College Dublin; Population Health Sciences Institute, Newcastle University & Falls and Syncope Service, Newcastle upon Tyne Hospital NHS Trust

**Keywords:** Orthostatic hypotension, postural hypotension, orthostatic intolerance, randomized controlled trial, pilot study, feasibility study, midodrine, fludrocortisone, conservative treatment

## Abstract

**Background:** Orthostatic hypotension (OH) is a common debilitating condition characterised by a significant drop in blood pressure (BP) on standing upright. Adults with OH are typically offered non-pharmacologic therapies, either alone or in combination with medication. The two most used agents are fludrocortisone and midodrine. There is a lack of good quality evidence for any of these treatments, all of which are in widespread clinical use. The aim of this internal pilot trial was to evaluate recruitment, attrition, treatment crossover and quality of outcomes.

**Methods:** The trial was designed as a pragmatic, open label, randomised, prospective, multicentre, superiority, multi-arm internal pilot. Within the 10-month pilot, a target of 64 adults with OH from 14 sites, was required to evaluate feasibility of recruitment, attrition, crossover and data collection.

Participants were randomised to one of three treatments: non-drug therapies (control), fludrocortisone plus non-drug therapies or midodrine plus non-drug therapies. Outcomes measured included symptoms, quality of life, activities of daily living, postural BP, use of health and care services, falls and safety. Participants received treatment and were followed-up for 12 months. Pre-planned criteria to progress from internal pilot were defined for recruitment, retention, crossover and outcome completion.

**Results:** Between the 3^rd^ December 2021 and 31^st^ August 2022, 13 participants were randomised from four of nine recruiting centres. Redeployment of clinical and research staff, due to COVID-19, limited the number of available sites. Participants already receiving fludrocortisone or midodrine accounted for 120 of 233 eligible participants being excluded. Due to the low sample size the rates of attrition and crossover are of limited value. Apart from the falls diaries, completion rates of the outcome measures were high. Due to low recruitment rates the pilot did not progress to the planned multi-arm multi-stage trial.

**Conclusions:** In its current design, this trial was not feasible. The main barriers to success were participants already receiving treatment and redeployment of clinical and academic staff during and after the COVID-19 pandemic.

**Trial registration:** ISRCTN 87213295, 23/07/2021, https://doi.org/10.1186/ISRCTN87213295

**Key messages regarding feasibility:** The clinical and cost effectiveness of different treatment strategies for orthostatic hypotension is unknown.

People with orthostatic hypotension tend to be older, have multiple long-term conditions and take multiple medications. There is uncertainty around recruitment of this population, attrition rates, potential crossover of treatments and completion of multiple outcome measures.

Redeployment of clinical and research staff during COVID-19 and the post-COVID period meant that trial delivery was not feasible.

Recent or current exposure to one of the trial interventions precluded a large proportion of participants being eligible.

A flexible and pragmatic trial protocol was not sufficient to overcome these barriers.

The number and timing of outcome measures appears to be appropriate and feasible.

## Background

Orthostatic hypotension (OH) is a common and disabling condition, characterised by a significant reduction in blood pressure (BP) on standing upright [1]. It is particularly prevalent in older populations and in those with chronic disease, affecting up to one in five community-dwelling older people, one in four with diabetes and one in three with Parkinson’s disease (PD) [2–4]. In the US, the presence of OH in people with PD is known to increase overall health care-related costs 2.5-fold compared to those who have PD without OH [5].

The reduction in BP during standing leads to a wide variety of symptoms including dizziness, headache, nausea, fatigue and visual disturbance; at its most severe it can result in falls and syncope [6, 7]. People with OH have a reduced quality of life due to the difficulties in performing even simple tasks which involve standing [8]. There are also longer-term sequelae, as OH is associated with an increased risk of stroke, cognitive impairment and all-cause mortality [9, 10].

Despite the high prevalence of OH, there is very little good quality evidence to support its management [11]. The UK’s National Institute for Health and Care Excellence (NICE) provides evidence summaries for fludrocortisone and midodrine to treat OH, but notes that long-term efficacy and safety is unclear [12, 13]. NICE also comments that these studies are limited by their use of disease-centred outcomes (i.e. BP) rather than patient-centred outcomes, such as symptoms and quality of life.

Following conservative measures, including physical counter manoeuvres and fluid and salt loading, the European Society of Cardiology recommends the use of midodrine or fludrocortisone for OH but notes that the quality of evidence is based on expert opinion and/or small studies and that further research is needed [14]. Similarly, systematic reviews and meta-analyses consistently describe existing evidence as poor quality, calling for more rigorous evaluation to guide clinical practice [14–18]. Both agents have side effects, but as their relative effectiveness is unclear, it is unknown whether the benefits outweigh their harms.

The medications to treat OH are inexpensive but the cost consequences of successful treatment or management of side effects are not known. No high-quality economic evaluations comparing these interventions, with each other or against non-pharmacologic therapies, have been identified. Therefore, the lack of evidence on the relative cost-effectiveness of fludrocortisone and midodrine means that there is insufficient evidence about which of these therapies represents a good use of scarce healthcare resources.

In 2018 the UK National Institute for Health and Care Research Health Technology Assessment (NIHR HTA) Programme opened a call for commissioned research on ‘Management of orthostatic hypotension (HTA 18/32)’. CONFORM-OH (CONtrol, Fludrocortisone OR Midodrine for Orthostatic Hypotension) was funded by NIHR HTA in 2019 and the protocol designed in line with the commissioning brief.

## Methods

The full protocol is available on the International Standard Randomised Controlled Trial Number website at https://doi.org/10.1186/ISRCTN87213295.

### Objectives

To address uncertainties in the study design the internal pilot evaluated the feasibility of reaching the recruitment target, the rate of attrition and the crossover rates between arms.

### Progression criteria and sample size

The aim was to open 20 sites in a staged approach, with 14 sites planned to be open by the end of the 10-month pilot. The participant sample size for the pilot was based on the monthly recruitment target required to achieve the sample size in a full trial. The estimated rate of randomisation was 0.8 participants per site per month with a target of 64 participants at 10 months.

A traffic light system was in place to judge feasibility at ten months:

- Recruitment
  - Green: 64 participants (or ≥0.8 participants per site per month)
  - Amber: 40 to 63 participants (or ≥0.5 to <0.8 participants per site per month), but with strategies identified to increase recruitment
  - Red: <40 participants (or <0.5 participants per site per month)
- Attrition
  - Green: ≤15% participants withdraw before the primary endpoint
  - Amber: 16 to 35% of participants withdraw before the primary endpoint but with strategies identified to improve retention
  - Red: >35% participants withdraw with no feasible solutions to improve
- Cross-over
  - As there were no existing data to estimate crossover rates, these were to be monitored and reviewed during the pilot with no fixed traffic-light criteria.
- Outcome data
  - The quality and completeness of outcome data were monitored to make improvements if required.

Feedback was sought from site staff during remote interviews at the end of the pilot; trial feedback questionnaires were sent to participants in the post following their last visit. Feedback was collated and summarised to identify barriers and solutions.

Progression or termination of the trial following the pilot was judged by the Trial Management Group, Data Monitoring Committee and Trial Steering Committee who made their recommendations to the Funder.

The planned sample size for the full trial can be found in the in the Statistical Analysis Plan which is available in supplementary material Document 1.

### Trial design

CONFORM-OH was designed as a pragmatic, multi-arm, multi-stage, parallel group, prospective, randomised, open label, superiority trial, with a 10-month internal pilot. Participants were randomised in a 1:1:1 ratio to conservative management, conservative management plus fludrocortisone or conservative management plus midodrine. The intervention and follow-up were for 12 months.

### Setting

Participants were recruited from multiple secondary care sites across the UK NHS which included falls and syncope clinics, movement disorders services, geriatrics clinics and cardiology clinics. Fully informed, written consent was obtained by a member of research team either in person, or remotely via telephone/video with consent forms posted to site.

### Participants

#### Inclusion Criteria

- Adults aged 18 years and over
- A clinical diagnosis of symptomatic OH which was either:
  - Clinically significant where treatment was indicated quickly without a trial of lifestyle modification OR
  - Refractory to an adequate period of lifestyle modification (to be judged clinically)
- A drop in systolic blood pressure of ≥20mmHg and/or a drop in diastolic blood pressure of ≥10mmHg, within three-minutes of standing upright from a supine position (or on tilt-testing)
- A score of ≥2 on the Orthostatic Hypotension Questionnaire
- Willing and able to provide informed consent

#### Exclusion Criteria

- OH secondary to acute or reversible causes
- Use of fludrocortisone or midodrine within the last six months
- Terminal illness or life expectancy <12 months
- Supine hypertension (where the risks of treatment outweigh the benefits)
- A known allergy to study medication
- A known contra-indication to fludrocortisone or midodrine which outweighs the potential clinical benefit
- Current or planned pregnancy/breast feeding during the trial
- Inability to communicate in English
- Inability to comply with the study procedures
- Currently taking part in another clinical trial that would interfere with the outcomes of CONFORM-OH

#### Changes to planned criteria

Originally the inclusion criteria required participants to have a diagnosis of OH which was refractory to a minimum of four weeks of lifestyle modification. This criterion was specified by the funder in the commissioned call for funding. However, as a pragmatic trial and to reflect clinical practice, this was modified as above. Allowing diagnosis by tilt-table testing was an additional modification made at one site’s request.

### Interventions

#### Control

Non-pharmacologic therapies (or conservative management) included trigger avoidance, physical counter-manoeuvres, fluid and salt intake, compression hosiery and ‘culprit’ medication review. Trial specific guidance on non-drug therapies was not provided to sites, as they were encouraged to follow their usual clinical practice. To account for sites’ differing practice, the randomisation method stratified by recruiting centre and data were collected from sites to describe which conservative measures they used.

#### Fludrocortisone

Sites were requested to initiate and titrate fludrocortisone tablets as they would in their usual clinical practice. Permissible dosing ranged from 50 micrograms per day to a maximum of 400 mcg per day. There were no restrictions on the brand used.

#### Midodrine

Sites were requested to initiate and titrate the midodrine as they would in their usual clinical practice. Dosing ranges typically started at a dose of 2.5 mg three times per day orally, increasing to a maximum tolerated dose, not exceeding 10 mg three times per day. Lower starting doses (e.g. 2.5 mg twice per day) were permissible. There were no restrictions on the brand used.

#### Randomisation and allocation

Allocation was via a central, secure, web-based system (Sealed Envelope™). Participants were assigned in a 1:1:1 ratio between control and two intervention arms using a minimisation algorithm with a random element. Age (>□80 vs □≤□80 years), aetiology (neurogenic vs non-neurogenic) and site were used as stratification factors. Participants, site investigators, research staff and trial team members were aware of treatment allocation. As a pragmatic trial, study investigators followed their usual clinical practice once the participant was allocated a treatment. Changes to the treatment arm were allowed if clinically indicated.

### Outcomes

#### Feasibility outcomes

Feasibility outcomes included the total number recruited, the average recruitment rate per site per month, the proportion of participants withdrawing from the trial, the proportion of participants crossing over between trial treatments, and the proportion of outcome assessments completed out of those expected.

#### Trial outcome measures

The primary outcome of the CONFORM-OH trial was the change in OH related symptoms from baseline to six months measured using the Orthostatic Hypotension Questionnaire (OHQ)[6].

Secondary outcomes are displayed in Table 1.

#### Data collection

Questionnaires were completed by participants either during research visits, remotely over the telephone or returned via post. For the EQ-5D-5L a validated proxy version was used for those participants not able to complete the questionnaire themselves.

Postural BP was performed by a clinician during either a research, usual clinic appointment or home visit. In cases where it was not possible for the participant to attend, a recent (within six weeks) postural BP recorded in the participant’s medical records was permissible. As a contingency measure, if the BP assessment was not possible, participants were able perform their own postural BP assessment at home after being provided with a study BP monitor and instructions [19]. If participants withdrew from the study, consent was requested from participants to use routinely collected clinical data.

Adverse events were identified through participant self-report or during questioning during research visits.

#### Statistical analysis

Due to the small number of participants recruited in the internal pilot, feasibility metrics and clinical outcome data are summarised descriptively. No hypothesis testing has been performed. Continuous outcomes are reported using the mean and standard deviation (SD) and/or the median and range (minimum and maximum values). Categorical outcomes are reported as frequencies and percentages.

All clinical outcome data are reported according to randomised treatment group, following the intention-to-treat principle. To account for crossovers between intervention groups, safety data are reported according to treatment strategy received.

The OHQ and NEADL questionnaires were scored according to published scoring methods[6, 20]. Full details can be found in the Statistical Analysis Plan which is available in supplementary material Document 1.

#### Health Economics analyses

Pilot data are summarised descriptively, continuous outcomes are reported using the mean and standard deviation (SD) and/or the median and range (minimum and maximum values). Categorical outcomes are reported as frequencies and percentages. For the E-5D-5L we report responses by the descriptive framework, the EQ-5D-VAS and health state utilities.

Full details, including the analysis methods which would have been used had the trial continued past the internal pilot stage, can be found in the Health Economics Analysis Plan which is available in supplementary material Document 2.

#### COVID-19

The process of setting up the CONFORM-OH study began in November 2019, with a planned recruitment start date of May 2020. In early 2020 the study set-up process was paused as COVID-19 studies were prioritised by ethical review committees and local research sponsor. In addition, several co-investigators were redeployed to support clinical work during the pandemic. Our Patient and Public experts informed the study team that they would not be confident to attend clinical or research visits until they were certain that reliable systems were in place to keep them safe from COVID-19 in these settings. In mid-2020, the CONFORM-OH study protocol was modified to make it more feasible during the pandemic. These modifications included:

- Allowing remote consent (telephone or video link)
- Accepting routinely measured blood pressure outcomes in lieu of research visit blood pressure
- Allowing participants to measure their own postural blood pressure if research or clinic visits were not possible
- Additional questions were included in the health economics outcomes about COVID-19

## Results

### Sites

In March 2020, 41 sites had expressed an interest to recruit. Of these, 20 did not return site feasibility questionnaires and were excluded. The process of gaining sponsorship and ethical permission took nine months. The first site opened in December 2021 with a further eight sites open by May 2022. Twelve sites were in the process of being set up when the pilot phase ended. Specific sites and their screening and recruitment figures can be found in Table 2.

### Recruitment

Between the 3^rd^ December 2021 and 31^st^ August 2022, 13 participants were randomised from four of the nine recruiting centres.

Progress against the internal pilot progression criteria was evaluated by the Trial Steering Committee on 30^th^ August 2022. Nine sites had been open to recruitment for a total of 52 site months and 13 participants had been randomised. The average accrual rate was 0.25 participants per site per month, which fell well below the amber progression criteria target and into the red category. On the recommendation of the Trial Steering Committee the trial closed to recruitment on 31^st^ August 2022, one month earlier than the original planned 10-month internal pilot phase. All participants who had been recruited continued in the trial as planned.

In total, 282 patients were screened for eligibility. Forty-nine patients (17%) were found to be eligible and of those 13 (27%) were randomised. Reasons for ineligibility and eligible patients not taking part in the trial are summarised in Table 3. The main reason for ineligibility was due to recent use of fludrocortisone or midodrine within the last six months (120 of 233 ineligible patients; 52%). In most cases (116; 97%), this was due to current use of either or both study medications. A contraindication to fludrocortisone or midodrine also led to the exclusion of 25 (11%) patients; predominantly contraindications were to midodrine.

Thirty six of the 49 (73%) eligible patients did not take part in the trial. Thirty (83%) declined participation; the burden of completing trial assessments was the most common reason given (10 of 30; 33%).

### Attrition

Participant flow through the trial is provided in a CONSORT diagram in Figure 1. Of the 13 participants randomised, three were allocated to the conservative management arm (control), four were allocated to conservative management plus fludrocortisone and six were allocated to conservative management plus midodrine. Of the three participants allocated to the control arm, two (67%) withdrew from the trial, one prior to the three-month visit and one prior to the six-month visit, and one further participant died prior to the three-month visit. Both participants who withdrew from the trial agreed for routinely available data (e.g. BP measurements) to still be collected from medical records where available. One participant allocated to the midodrine arm was lost to follow-up prior to the 12-month follow-up visit. Overall, 11 (85%) participants remained in follow-up at the three-month visit, 10 (77%) participants at the six-month visit, and nine (69%) participants at the twelve-month visit.

### Treatment adherence and crossover

Of the four participants allocated to the fludrocortisone arm, one (25%) participant decided not to take their allocated study medication but continued to provide follow-up data. Of the six participants allocated to the midodrine arm, two (33%) participants discontinued their allocated treatment, one participant crossed over to fludrocortisone at the three-month visit due to side effects of midodrine and one participant stopped taking midodrine at the six-month visit as their symptoms had improved.

### Outcome measure completion

At the six-month primary outcome timepoint, data were available for ten (77%) participants; zero (0%) allocated to control, four (100%) fludrocortisone and six (100%) midodrine. Completion rates of outcome measures are summarised in Table 4. The OHQ, NEADL questionnaire, and BP measurements were available for all participants who remained in follow-up at each timepoint. No additional BP measurements were provided from medical records for either of the two participants who withdrew from the trial but allowed for counited collection of routinely available data. Falls data were collected from all participants who remained in follow-up at each timepoint, however falls diaries were only completed and returned by seven (64%) participants at the three-month visit, six (60%) participants at the six-month visit and five (56%) participants at the twelve-month visit.

The barriers to recruitment, assessment of the feasibility components and lessons learnt from the internal pilot are summarised in Table 5.

### Demographic data

Participant characteristics, medication at baseline and conservative measures advised at baseline are summarised descriptively in Table 6 and supplementary data Table S1. The average age of all participants was 74 years (SD 9.6), 7 (54%) were men, and all (100%) were of white background. Five (38%) participants had orthostatic hypotension caused by neurologic disorders, four of which were Parkinson’s disease. At baseline, the mean OHQ score was 6.4 (SD 1.5), the mean systolic postural BP drop was 40 mmHg (SD 16), and the mean diastolic BP drop was 10 mmHg (SD 16).

### Trial outcome measures

Data collected on primary and secondary clinical outcome measures are summarised descriptively in supplementary materials. No data were available for any participant allocated to the control arm at six- or 12-month follow-up.

### OHQ, activity of daily living, nadir standing blood pressure, orthostatic blood pressure drop

For each outcome measure, the change from baseline to each follow-up timepoint is summarised descriptively by randomised treatment group in supplementary data Table S2. Individual participant data are also plotted in supplementary data Figure S1.

The mean change in the OHQ score from baseline to six months was −4.5 points (N = 4; SD 2.0) in those allocated to conservative management plus fludrocortisone and −1.7 points (N = 6; SD 2.0) in those allocated to conservative management plus midodrine.

Summary data for each outcome measure at each timepoint are also provided in supplementary data Table S3. The correlation between baseline and follow-up OHQ scores is presented in supplementary data Table S4.

#### Falls

Six participants reported at least one fall during the 12-month follow-up period; one (25%) participant allocated to conservative management plus fludrocortisone and five (83%) allocated to conservative management plus midodrine (supplementary Table S5). In total, two falls were reported in the fludrocortisone arm and 46 in the midodrine arm (34 falls were reported by one participant). No syncopal events were reported by participants allocated to fludrocortisone and nine syncopal events were reported by one participant allocated to the conservative management plus midodrine arm. No data on falls or syncopal events were available for participants allocated to the control arm as the one participant who remained in follow-up at the three-month visit did not return their falls diary. Three participants reported fall-related injuries during the 12-month follow-up, summarised in supplementary Table S6.

#### Safety

Safety data are summarised in supplementary Table S7. In total, 26 adverse events were reported; four in participants allocated to conservative management, 16 in the conservative management plus fludrocortisone arm and six in the conservative management plus midodrine arm. Of those, five adverse reactions (adverse events considered to be possibly, probably or definitely related to trial treatment) were reported from three (50%) participants exposed to midodrine. All were mild in severity, however for one participant this led to a permanent discontinuation of trial treatment and a temporary interruption of treatment for a further participant. Of the 26 adverse events reported, nine were reported as serious adverse events from four participants, all were deemed to be unrelated to trial treatment. One adverse event resulted in death. Further detail is provided in the supplementary tables S8-S10.

### Health Economics

Supplementary tables S11-S30 report the summary data for the use of health and care services. These are reported by the randomised arm and for all time points. Due to the limited data, no comparisons are made between trial arms and no totals are presented for each area of resource use. For contacts with general practitioners there is a right skew in the data, with a small number of participants having a larger number of contacts. The same pattern is not clearly observed for most other areas of resource use because the use of services is very low. Similarly, the use of private health care and personal social services is likewise infrequent in every group. However, the small number of participants means firm conclusions cannot be drawn.

Supplementary tables S31 reports response rates for the EQ-5D-5L which describes pattern of responses to the health care services data. The EQ-5D-5L could be either self completed or completed by proxy. Very few participants had proxy completion for the EQ-5D-5L at any timepoint (Table S32). Tables S33-S37 provides summary data for each of the 5 dimensions of the EQ-5D –5L separately. At baseline, no participant was in the worst state of health for Mobility. Overall, the mean and median score was 1.75 and 1.5 retrospectively, indicating participants had slight problems with mobility (Table S33). For self-care, the mean score was 1.67 indicating that overall that participants had slight problems with washing or dressing themselves; similar inferences can be made from the median score which was 1 (Table S34). Responses for usual activity are shown in Table S35. At baseline, the mean score for usual activities was just over 2, and the median score was 1. The mean score indicates that participants had slight problems doing their usual activities, and the median score for usual activities indicated that participants had no problems doing them. Table S36 shows responses for the pain and discomfort dimension. This table shows that the mean and median scores were 2.5 and 3, respectively, suggesting that participants had moderate pain and discomfort. Table S37 reports responses by the anxiety and depression dimensions. The mean and median scores were 2.33 and 2, respectively. Theses scores suggest that participants were slightly anxious or depressed. Table S38 shows the responses for the EQ-5D-VAS. At baseline, the mean and median scores are 63 and 60 respectively (100 is best imaginable health and 0 is the worst possible health). The mean and median baseline EQ-5D-5L utility values which can be found in Table S39 were 0.58 and 0.62 respectively (1 is the value of full health and 0 is equivalent to dead). For all dimensions, it was unclear if the severity of responses varied over the follow-up. This is due to the small number of participants completing the EQ-5D-5L.

One participant provided EQ-5D-5L by proxy at baseline, and at 12 months, two participants provided proxy data at 6 months. Tables S40 to S45 show the responses for each dimension for proxy response. As so few data were available, summary text is not provided. Table S46 shows the responses for the EQ-5D-VAS proxy responses at baseline was 45. As so few data were available summary text is also not provided.

## Discussion

This randomised controlled internal pilot did not meet its progression criteria, suggesting the trial was not feasible as planned. The timing of the trial, originally planned to open in 2020, was unfortunate as the effects of the COVID-19 pandemic had a severe impact on research sites. Several sites were unable to recruit due to staff redeployment, either to other clinical duties, or to focus on COVID-19 clinical trials. COVID-19 also had an impact on clinical pathways. Sites noted that an increase in virtual consultations, resulted in fewer postural BP measurements and fewer diagnoses of OH. From a recruitment perspective, the biggest barrier was the eligibility criteria which excluded participants who were taking, or had recently tried the medications under investigation, this amounted to just over half of screened participants.

Of the 13 participants recruited, the primary outcome was available in ten. With such small numbers, it is difficult to estimate whether these figures would be helpful to estimate attrition in a larger study. One potential solution to reduce attrition at the primary endpoint would be to move this from six to three months. Indeed, looking at the individual results in Supplement Figure S1, it appears that most of the change seen from baseline occurs at the three-month time point, with little additional gain at six months. However, feedback from participants and from site investigators was that six months allows adequate time to titrate medication doses. At the primary outcome time point (six months), all participants from the control arm had withdrawn or died, in contrast to no withdrawals in the medication arms. Being randomised to the control arm was raised as a concern by some sites in their post-trial feedback, as some participants did not want to be allocated to control. This is an important factor to consider for future trials. However, it should be stressed that it remains unknown whether medication is superior to non-drug therapy, and this is an important question which needs answering. The participant and clinician equipoise could be addressed through novel trial design. Had the trial progressed from the pilot, the main trial was designed as an adaptive multi-arm multi-stage (MAMS) trial. An interim analysis was planned after the 200th participant had been randomised which would have allowed the study to drop an intervention arm if the interim analysis suggested it was no more effective than control. However, this was precluded by low recruitment. However, other novel designs could be considered. For example, a Personalised Randomised Controlled Trial (PRACTICAL[21]) design allows randomisation to acceptable arms only, such that if a contra-indication/exclusion to one intervention existed, the participant would not be randomised to that arm. This may address the issue of clinicians and participants not wanting to be randomised to control, or participants having a contraindication to one of the medications.

The completion rates of the secondary outcomes were excellent at all time points for symptoms, activities of daily living, and BP. There were two exceptions. One was the completion rates of falls diaries. However, this is not unusual in the field of falls-related research. One participant fed back that they would have preferred an online falls diary, and this this should be explored as an option for future trials, with the possibility of completing all the outcomes remotely. The number of outcome measures in the trial was not considered too many by most participants. In fact, feedback from participants was very positive, with no negative experiences reported by respondents in the post-trial feedback. The second exception was the reporting of BP for those that withdrew but consented to the use of routinely collected medical data, this applies to two participants. For these participants, BP measurements were recorded, while enrolled, however after withdrawing no measurements were reported. It is not clear if no measurements were available or if these were not obtained. For future trials, consideration may be needed on how to incorporate routinely collected data.

An additional uncertainty when developing the trial protocol was the expected level of crossover between intervention arms. Only one participant changed their allocated intervention arm. Unfortunately, the numbers of participants in the pilot were too low to provide robust data for future estimates, but as far as we are aware, it is the only current data available to inform this.

It is uncertain whether the trial may now be more feasible in the post-COVID-19 period. Modified clinical pathways are now established and there is a greater recognition that non-COVID-19 clinical services and research should continue even during future outbreaks (with safety measures in place). The trial was designed to be as pragmatic and flexible as possible, to match usual clinical pathways, to make recruitment and trial processes simpler for participants and clinician investigators. However, one of the limitations which became apparent was that there was considerable heterogeneity in usual practice between sites, so the study pathways were not always easily deliverable for sites. In addition, while the pragmatic nature attempted to reduce the workload for sites, there was a lack of available research support at sites to help deliver the trial in clinical settings. There may be an argument for increased funding to provide more research support, rather than depending on clinicians to deliver pragmatic clinical trials.

It is more difficult to address the exclusion criteria of currently taking treatment which led to the exclusion of large numbers of people with OH. To include these participants would require a ‘washout’ period off treatment, to measure baseline outcomes, which may be considered unacceptable by some patients and clinicians. Furthermore, their inclusion would introduce bias, as previous exposure to treatment will influence an individual’s assessment of treatment response, this is particularly relevant with the primary outcome being subjective. One solution would be to conceal treatment allocation in a placebo-controlled trial. However, participants and clinicians may be unwilling to stop treatment with the chance of being randomised to placebo. Alternatively, identifying treatment-naïve people with OH could be done in primary care. However, the coding of orthostatic hypotension in primary care is very poor (24, 973 cases among 2,911,260 records over ten years)[22]. BP screening for OH in primary care is also unlikely to be an efficient recruitment method. Not all people with OH would require medication and screening for OH does not reflect current clinical practice and may produce skewed results. This has been seen in falls intervention research where interventions have been shown to be successful in those recruited from falls services, but not in those screened in primary care[23, 24].

## Conclusions

The CONFORM-OH internal pilot showed a definitive study would not be feasible as planned. The main barriers to success were participants already receiving treatment and redeployment of clinical and academic staff during and after the COVID-19 pandemic.

## List of abbreviations

OH: orthostatic hypotension
BP: blood pressure
PD: Parkinson’s disease
NICE: National Institute for Health and Care Excellence
NIHR HTA: National Institute for Health and Care Research Health Technology Assessment
CONFORM-OH: Control, fludrocortisone or midodrine for orthostatic hypotension
OHQ: Orthostatic hypotension questionnaire
NEADL: Nottingham extended activities of daily living
MEDDRA: Medical Dictionary for Regulatory Activities
QALY: Quality adjusted life year

## Declarations

None

## Ethics approval and consent to participate

All participants provided fully informed, written consent. The study was approved by the UK NHS Health Research Authority’s Newcastle & North Tyneside 1 Research Ethics Committee (reference 21/NE/0083).

## Consent for publication

Not applicable

## Availability of data and materials

A de-identified dataset will be prepared and stored by Newcastle University. Requests for data sharing will be subject to request which should provide a clear purpose, analysis plan, how the results will be disseminated, and who the authors will be. Data transfer will be subject to completion of a Data Sharing Agreement between Newcastle University and the end users.

## Trial Registration

ISRCTN Registry: ISRCTN 87213295, registered 23^rd^ July 2021: https://www.isrctn.com/ISRCTN87213295

## Author’s contributions

All authors have contributed to writing, reviewing and revising the manuscript. JF, SP, MW, JW, HM, LV, AY and RAK secured financial support for the trial and developed the trial protocol. SAA and HM performed the statistical analysis. TS and LV performed the health economic evaluation. EC, GW, NM and HH were responsible for management of all research activity and execution including provision of trial resources. JP curated the trial database.

## Acknowledgements

The authors and investigators are grateful to the clinical research staff at the following centres: Dumfries and Galloway Royal Infirmary; Gateshead Health NHS Foundation Trust; Lewisham and Greenwich NHS Trust; Newcastle Hospitals NHS Foundation Trust; Norfolk and Norwich University Hospitals NHS Foundation Trust; Northern Care Alliance NHS Foundation Trust; Royal Devon University Healthcare NHS Foundation Trust; Royal United Hospitals Bath NHS Foundation Trust and Walsall Healthcare NHS Trust. The authors and investigators are also grateful to the following Newcastle Clinical Trials Unit staff for their help in managing and delivering the trial: Rebecca Maier, Georgina Browne, Rebecca Wilson, Michelle Bardgett, Dawn Clewes, Laura Simms, Eva Holstein, Anneka Kershaw, Laura Robertson; and to Ian Campbell, Craig Alderson and Penny Bradley for providing clinical trials pharmacy guidance and review on behalf of sponsor. The authors and investigators are also grateful to the patient and public involvement advocate Ruth Pearce and the patient expert advisors’ group. The authors and investigators are also grateful to the members of the Trial Steering Committee: Helen Roberts, Maw Pin Tan, Andrew Clegg and Brian Maxwell and the members of the Data Monitoring Committee: Roy Soiza, Babak Choodari-Oskooei and Susan Shenkin. Finally, the authors and investigators would like to thank the trial participants.

## References

1. Freeman R, Wieling W, Axelrod FB, Benditt DG, Benarroch E, Biaggioni I, Cheshire WP, Chelimsky T, Cortelli P, Gibbons CH et al: Consensus statement on the definition of orthostatic hypotension, neurally mediated syncope and the postural tachycardia syndrome. Clin Auton Res 2011, 21(2):69–72.

2. Zhou Y, Ke SJ, Qiu XP, Liu LB: Prevalence, risk factors, and prognosis of orthostatic hypotension in diabetic patients: A systematic review and meta-analysis. Medicine (Baltimore) 2017, 96(36):e8004.

3. Velseboer DC, de Haan RJ, Wieling W, Goldstein DS, de Bie RM: Prevalence of orthostatic hypotension in Parkinson’s disease: a systematic review and meta-analysis. Parkinsonism & Related Disorders 2011, 17(10):724–729.

4. Saedon NIz, Tan MP, Frith J: The Prevalence of Orthostatic Hypotension: A Systematic Review and Meta-Analysis. The Journals of Gerontology: Series A 2018:gly188–gly188.

5. Merola A, Sawyer RP, Artusi CA, Suri R, Berndt Z, Lopez-Castellanos JR, Vaughan J, Vizcarra JA, Romagnolo A, Espay AJ: Orthostatic hypotension in Parkinson disease: Impact on health care utilization. Parkinsonism Relat Disord 2018, :45–49.

6. Kaufmann H, Malamut R, Norcliffe-Kaufmann L, Rosa K, Freeman R: The Orthostatic Hypotension Questionnaire (OHQ): validation of a novel symptom assessment scale. Clin Auton Res 2012, 22(2):79–90.

7. McDonald C, Pearce M, Kerr SR, Newton J: A prospective study of the association between orthostatic hypotension and falls: definition matters. Age Ageing 2016.

8. Kim N, Park J, Hong H, Kong ID, Kang H: Orthostatic hypotension and health-related quality of life among community-living older people in Korea. Qual Life Res 2020, 29(1):303–312.

9. Angelousi A, Girerd N, Benetos A, Frimat L, Gautier S, Weryha G, Boivin JM: Association between orthostatic hypotension and cardiovascular risk, cerebrovascular risk, cognitive decline and falls as well as overall mortality: a systematic review and meta-analysis. J Hypertens 2014, 32(8):1562–1571.

10. Ricci F, Fedorowski A, Radico F, Romanello M, Tatasciore A, Di Nicola M, Zimarino M, De Caterina R: Cardiovascular morbidity and mortality related to orthostatic hypotension: a meta-analysis of prospective observational studies. Eur Heart J 2015, 36(25):1609–1617.

11. Frith J, Parry SW: New Horizons in orthostatic hypotension. Age Ageing 2017, 46(2):168– 174.

12. NICE [ESUOM20]: Postural hypotension in adults: fludrocortisone. NICE Evidence summary 2013.

13. NICE [ESNM61]: Orthostatic hypotension due to autonomic dysfunction: midodrine. NICE Evidence summary 2015.

14. Brignole M, Moya A, de Lange FJ, Deharo J-C, Elliott PM, Fanciulli A, Fedorowski A, Furlan R, Kenny RA, Martín A et al: 2018 ESC Guidelines for the diagnosis and management of syncope. Eur Heart J 2018, 39(21):1883–1948.

15. Logan IC, Witham MD: Efficacy of treatments for orthostatic hypotension: a systematic review. Age Ageing 2012, 41(5):587–594.

16. Mills PB, Fung CK, Travlos A, Krassioukov A: Nonpharmacologic management of orthostatic hypotension: a systematic review. Arch Phys Med Rehabil 2015, 96(2):366–375 e366.

17. Ong AC, Myint PK, Shepstone L, Potter JF: A systematic review of the pharmacological management of orthostatic hypotension. Int J Clin Pract 2013, 67(7):633–646.

18. Parsaik AK, Singh B, Altayar O, Mascarenhas SS, Singh SK, Erwin PJ, Murad MH: Midodrine for orthostatic hypotension: a systematic review and meta-analysis of clinical trials. J Gen Intern Med 2013, 28(11):1496–1503.

19. Gibbon JR, Parry SW, Witham MD, Yarnall A, Frith J: Feasibility, reliability and safety of self-assessed orthostatic blood pressure at home. Age Ageing 2022, 51(7).

20. Nouri FM, Lincoln NB: An extended activities of daily living scale for stroke patients. Clin Rehabil 1987, 1(4):301–305.

21. Walker AS, White IR, Turner RM, Hsu LY, Yeo TW, White NJ, Sharland M, Thwaites GE: Personalised randomised controlled trial designs-a new paradigm to define optimal treatments for carbapenem-resistant infections. Lancet Infect Dis 2021, 21(6):e175–e181. 22.

22. Bhanu C, Petersen I, Orlu M, Davis D, Walters K: Incidence of postural hypotension recorded in UK general practice: an electronic health records study. Br J Gen Pract 2023, 73(726):e9– e15.

23. Lamb SE, Bruce J, Hossain A, Ji C, Longo R, Lall R, Bojke C, Hulme C, Withers E, Finnegan S et al: Screening and Intervention to Prevent Falls and Fractures in Older People. N Engl J Med 2020, 383(19):1848–1859.

24. Gillespie LD, Robertson MC, Gillespie WJ, Sherrington C, Gates S, Clemson LM, Lamb SE: Interventions for preventing falls in older people living in the community. Cochrane Database of Systematic Reviews 2012, 9:pCD007146.

