## Supplementary for "Control, Fludrocortisone or Midodrine for the treatment of Orthostatic Hypotension (CONFORM-OH): Results from an internal pilot randomised controlled trial"

Online supplement

Contents

1. Trial data
   - Clinical outcome data
     i. Table S1: ‘Culprit’ medication at baseline
     ii. Table S2: Changes from baseline in primary and secondary outcome measures
     iii. Figure S1. Changes from baseline, individual outcome data plots
     iv. Table S3: Summary of outcome data at each time point
     v. Table S4. Correlation between baseline and follow-up OHQ scores
     vi. Table S5: Falls and syncope
     vii. Table S6. Falls-related injuries
   - Safety data
     i. Table S7: Adverse events
     ii. Table S8: Adverse events and severity per treatment arm
     iii. Table S9: Adverse reactions
     iv. Table S10: Serious adverse events
   - Health Economic Data
     i. Table S11: Frequency of GP Appointments
     ii. Table S12: Frequency of Nurse Appointments
     iii. Table S13: Frequency of Telephone GP Appointments
     iv. Table S14: Frequency of Telephone Nurse Appointments
     v. Table S15: Frequency of Doctor Visits at Home
     vi. Table S16: Frequency Walk in Centre appointments
     vii. Table S17: Frequency of ambulance services
     viii. Table S18: Frequency of Accident and Emergency services
     ix. Table S19: Frequency of other appointments in hospital
     x. Table S20: Frequency of private appointments
     xi. Table S21: Frequency of personal social service
     xii. Table S22: Frequency of Personal Guarantee Credit
     xiii. Table S23: Frequency of Attendance Allowance
     xiv. Table S24: Frequency of Personal Independence Payment (Daily Living)
     xv. Table S25: Frequency of Personal independent payment (Mobility)
     xvi. Table S26: Frequency of Employment and Support Allowance
     xvii. Table S27: Frequency of Universal Credit
     xviii. Table S28: Frequency of Carer’s Allowance
     xix. Table S29: Frequency Social Worker Service
     xx. Table S30: Frequency of Community Base Health Services
     xxi. Table S31: Completion rates for the EQ-5D-5L (both Self and Proxy completed)
     xxii. Table S32: Method of EQ-5D-5L data collection by study group
     xxiii. Table S33: EQ-5D-5L Mobility Score
     xxiv. Table S34: EQ-5D-5L Self Care
     xxv. Table S35: EQ-5D-5L Usual Activity (e.g. work, study, housework, family or leisure activities)
     xxvi. Table S36: EQ-5D-5L Pain/Discomfort
     xxvii. Table S37: EQ-5D-5L Anxiety and Depression
     xxviii. Table S38: Health Score Self EQ-5D VAS
     xxix. Table S39: EQ-5D-5L Utility
     xxx. Table S40: EQ-5D-5L Mobility proxy Score
     xxxi. Table S41: EQ-5D-5L Self Care proxy Score
     xxxii. Table S42: EQ-5D-5L Usual Activities proxy Score (e.g. work, study, housework, family or leisure activities)
     xxxiii. Table S43: Pain and Discomfort Proxy Score
     xxxiv. Table S44: EQ-5D-5L Anxiety and Depression Proxy Score
     xxxv. Table S45: EQ-5D-5L Proxy Utility Score
     xxxvi. Table S46: EQ-VAS Proxy Score
2. Documents
1. Statistical analysis plan
2. Health economic analysis plan

Table S1: ‘Culprit’ medication at baseline

|  | Conservative management | Conservative management plus fludrocortisone | Conservative management plus midodrine | Total |
| --- | --- | --- | --- | --- |
|  | (N = 3) | (N = 4) | (N = 6) | (N = 13) |
| Levodopa | 1 (33%) | 0 (0%) | 3 (50%) | 4 (31%) |
| SSRI antidepressants | 0 (0%) | 1 (25%) | 2 (33%) | 3 (23%) |
| Tricyclic antidepressant | 0 (0%) | 1 (25%) | 1 (17%) | 2 (15%) |
| Dopamine agonist | 0 (0%) | 0 (0%) | 2 (33%) | 2 (15%) |
| COMT inhibitors | 0 (0%) | 0 (0%) | 2 (33%) | 2 (15%) |
| MAOI | 0 (0%) | 0 (0%) | 1 (17%) | 1 (8%) |
| Anti-psychotics | 0 (0%) | 1 (25%) | 0 (0%) | 1 (8%) |
| Other vasodilator | 0 (0%) | 0 (0%) | 1 (17%) | 1 (8%) |
| Alpha-blocker | 0 (0%) | 0 (0%) | 1 (17%) | 1 (8%) |
| Beta-blocker | 1 (33%) | 0 (0%) | 0 (0%) | 1 (8%) |
| Calcium channel blocker | 0 (0%) | 0 (0%) | 0 (0%) | 0 (0%) |
| Diuretic | 0 (0%) | 0 (0%) | 0 (0%) | 0 (0%) |
| ACE-inhibitor | 0 (0%) | 0 (0%) | 0 (0%) | 0 (0%) |
| Angiotensin II Receptor Blocker | 0 (0%) | 0 (0%) | 0 (0%) | 0 (0%) |
| Nitrates (regular, not prn) | 0 (0%) | 0 (0%) | 0 (0%) | 0 (0%) |
| Other antihypertensive | 0 (0%) | 0 (0%) | 0 (0%) | 0 (0%) |

SSRI: Selective serotonin reuptake inhibitor; COMT: catechol-O-methyltransferase; MAOI: Monoamine oxidase inhibitor; ACE: Angiotensin-converting-enzyme

Table S2: Changes from baseline in primary and secondary outcome measures

|  | **3-month follow-up** | | | **6-month follow-up** | | | **12-month follow-up** | | |
| --- | --- | --- | --- | --- | --- | --- | --- | --- | --- |
|  | Conservative management | Conservative management plus fludrocortisone | Conservative management plus midodrine | Conservative management | Conservative management plus fludrocortisone | Conservative management plus midodrine | Conservative management | Conservative management plus fludrocortisone | Conservative management plus midodrine |
|  | N= 1 | N = 4 | N = 6 | N= 0 | N = 4 | N = 6 | N= 0 | N = 4 | N = 5 |
| **OHQ** |  |  |  |  |  |  |  |  |  |
| Mean (SD) | -6.9 (.) | -4.2 (2.2) | -2.4 (3.6) | NA | -4.5 (2.0) | -1.7 (2.0) | NA | -4.9 (2.6) | -1.3 (2.7) |
| Median (Range) | -6.9 (-6.9, -6.9) | -3.9 (-7.0, -1.9) | -2.1 (-8.5, 1.3) | NA | -5.0 (-6.0, -1.8) | -2.0 (-4.4, 0.9) | NA | -5.6 (-7.2, -1.3) | -0.4 (-4.8, 2.2) |
| **NEADL** |  |  |  |  |  |  |  |  |  |
| Mean (SD) | -6.0 (.) | 2.3 (9.0) | -0.3 (4.1) | NA | 3.3 (8.8) | -1.0 (3.2) | NA | 3.3 (8.7) | -0.8 (3.7) |
| Median (Range) | -6.0 (-6.0, -6.0) | 0.0 (-6.0, 15.0) | 0.5 (-8.0, 3.0) | NA | 0.5 (-4.0, 16.0) | -0.5 (-7.0, 2.0) | NA | 0.0 (-3.0, 16.0) | -1.0 (-4.0, 5.0) |
| **Nadir standing blood pressure** |  |  |  |  |  |  |  |  |  |
| Systolic |  |  |  |  |  |  |  |  |  |
| Mean (SD) | -21.0 (.) | 23.3 (30.5) | 9.2 (29.1) | NA | 23.5 (27.0) | 26.3 (21.8) | NA | 18.8 (32.4) | 18.4 (27.2) |
| Median (Range) | -21 (-21, -21) | 21 (-11, 62) | 10 (-33, 52) | NA | 26 (-5, 48) | 30 (-1, 60) | NA | 20 (-14, 49) | 17 (-8, 63) |
| Diastolic |  |  |  |  |  |  |  |  |  |
| Mean (SD) | -20.0 (.) | 10.5 (18.7) | 0.8 (16.3) | NA | 12.5 (18.5) | 8.7 (14.2) | NA | 11.3 (18.6) | -0.2 (10.2) |
| Median (Range) | -20 (-20, -20) | 9 (-9, 34) | 4 (-25, 18) | NA | 10 (-5, 35) | 5 (-7, 26) | NA | 7 (-4, 36) | -7 (-8, 11) |
| **Postural blood pressure drop** |  |  |  |  |  |  |  |  |  |
| Systolic |  |  |  |  |  |  |  |  |  |
| Mean (SD) | -12.0 (.) | -17.0 (17.8) | -14.5 (23.3) | NA | -4.5 (19.8) | -21.2 (31.6) | NA | -7.8 (18.1) | -20.0 (19.6) |
| Median (Range) | -12 (-12, -12) | -22 (-31, 7) | -15 (-46, 10) | NA | 3 (-33, 9) | -13 (-61, 7) | NA | -4 (-32, 8) | -26 (-45, 1) |
| Diastolic |  |  |  |  |  |  |  |  |  |
| Mean (SD) | 8.0 (.) | 1.8 (20.2) | -3.3 (19.9) | NA | 2.8 (15.3) | -4.7 (21.8) | NA | -2.0 (13.8) | -1.2 (12.1) |
| Median (Range) | 8 (8, 8) | 5 (-25, 23) | -4 (-28, 21) | NA | 3 (-14, 19) | -7 (-32, 31) | NA | 3 (-22, 9) | -7 (-10, 19) |

Figure S1. Changes from baseline, individual outcome data plots

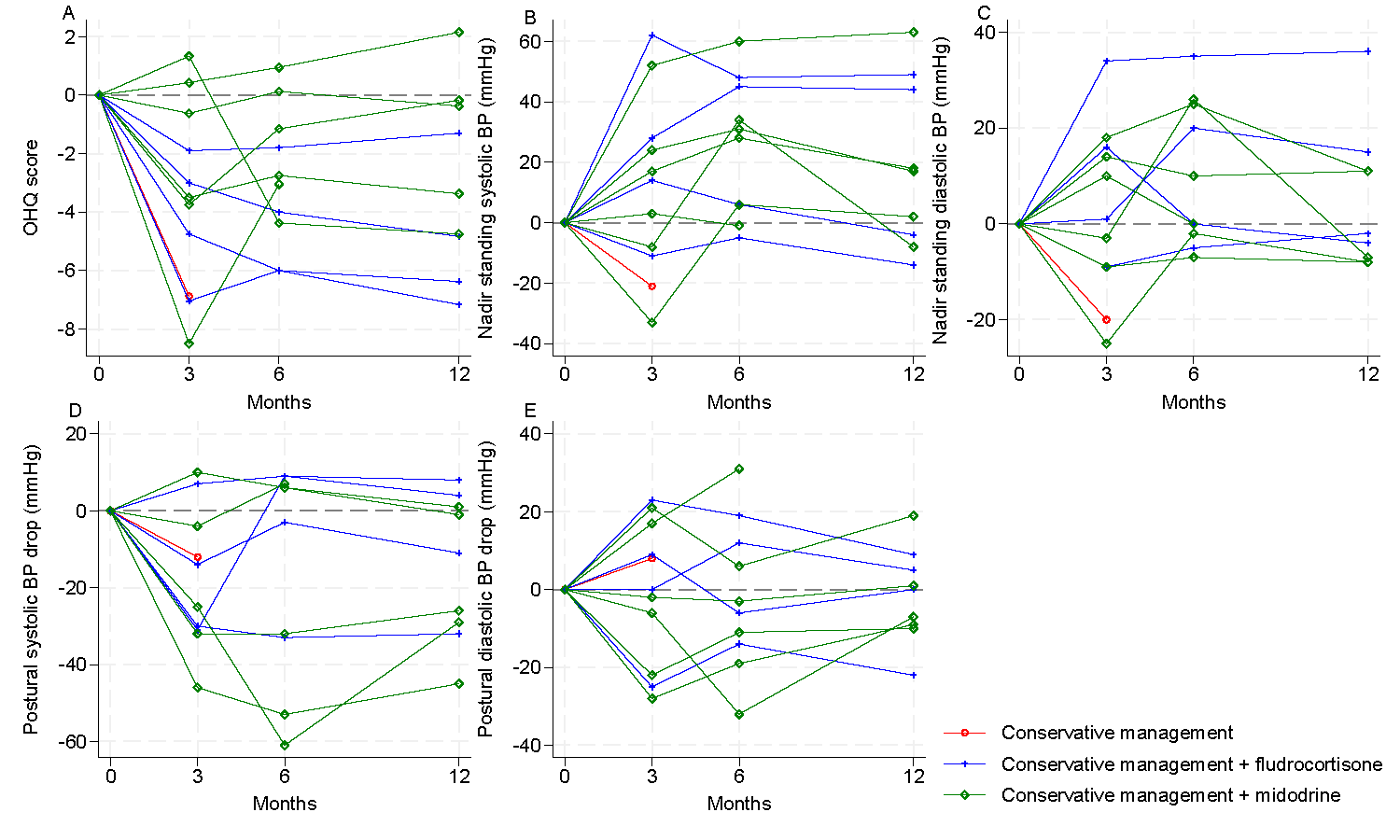

Table S3: Summary of outcome data at each time point

|  | **3-month follow-up** | | | **6-month follow-up** | | | **12-month follow-up** | | |
| --- | --- | --- | --- | --- | --- | --- | --- | --- | --- |
|  | Conservative management | Conservative management plus fludrocortisone | Conservative management plus midodrine | Conservative management | Conservative management plus fludrocortisone | Conservative management plus midodrine | Conservative management | Conservative management plus fludrocortisone | Conservative management plus midodrine |
|  | N= 1 | N = 4 | N = 6 | N= 0 | N = 4 | N = 6 | N= 0 | N = 4 | N = 5 |
| **OHQ** |  |  |  |  |  |  |  |  |  |
| Mean (SD) | 0.6 (.) | 2.6 (1.7) | 3.6 (3.0) | NA | 2.3 (0.9) | 4.3 (1.9) | NA | 1.9 (1.6) | 4.2 (2.4) |
| Median (Range) | 0.6 (0.6, 0.6) | 3.2 (0.1, 4.0) | 4.2 (0.0, 7.3) | NA | 2.5 (1.2, 3.3) | 4.8 (1.6, 6.3) | NA | 1.9 (0.0, 3.8) | 3.6 (1.2, 7.5) |
| **NEADL** |  |  |  |  |  |  |  |  |  |
| Mean (SD) | 2.0 (.) | 18.3 (2.2) | 13.8 (7.3) | NA | 19.3 (1.5) | 13.2 (6.6) | NA | 19.3 (1.0) | 12.8 (7.6) |
| Median (Range) | 2.0 (2.0, 2.0) | 18.0 (16.0, 21.0) | 15.0 (5.0, 21.0) | NA | 19.0 (18.0, 21.0) | 14.5 (5.0, 20.0) | NA | 19.5 (18.0, 20.0) | 9.0 (6.0, 22.0) |
| **Nadir standing blood pressure** |  |  |  |  |  |  |  |  |  |
| Systolic |  |  |  |  |  |  |  |  |  |
| Mean (SD) | 59.0 (.) | 112.0 (22.7) | 114.2 (31.4) | NA | 112.3 (23.4) | 131.3 (32.2) | NA | 107.5 (28.3) | 124.4 (33.4) |
| Median (Range) | 59 (59, 59) | 115 (83, 135) | 106 (77, 171) | NA | 110 (89, 141) | 118 (99, 179) | NA | 105 (80, 140) | 112 (97, 182) |
| Diastolic |  |  |  |  |  |  |  |  |  |
| Mean (SD) | 26.0 (.) | 72.0 (11.0) | 66.2 (14.8) | NA | 74.0 (18.5) | 74.0 (17.7) | NA | 72.8 (17.3) | 66.4 (10.8) |
| Median (Range) | 26 (26, 26) | 76 (56, 80) | 71 (46, 85) | NA | 71 (58, 97) | 75 (53, 102) | NA | 73 (54, 92) | 66 (52, 82) |
| **Postural blood pressure drop** |  |  |  |  |  |  |  |  |  |
| Systolic |  |  |  |  |  |  |  |  |  |
| Mean (SD) | 39.0 (.) | 22.8 (28.7) | 21.8 (22.2) | NA | 35.3 (25.6) | 15.2 (31.8) | NA | 32.0 (22.3) | 18.2 (22.8) |
| Median (Range) | 39 (39, 39) | 23 (-8, 54) | 22 (-7,57) | NA | 37 (3, 65) | 25 (-31, 53) | NA | 34 (4, 57) | 12 (-6, 46) |
| Diastolic |  |  |  |  |  |  |  |  |  |
| Mean (SD) | 39.0 (.) | 10.8 (13.5) | 5.8 (15.9) | NA | 11.8 (18.2) | 4.5 (18.8) | NA | 7.0 (12.2) | 11.0 (13.2) |
| Median (Range) | 39 (39, 39) | 11 (-4, 25) | 5 (-14, 28) | NA | 15 (-12, 30) | 8 (-28, 25) | NA | 6 (-6, 22) | 13 (-3, 26) |

Table S4. Correlation between baseline and follow-up OHQ scores

| OHQ score | Pearson’s correlation coefficient | | | |
| --- | --- | --- | --- | --- |
|  | Control | Fludrocortisone | Midodrine | Overall |
| Month 3 | N/A | 0.06 | -0.13 | -0.19 |
| Month 6 | N/A | -0.27 | 0.36 | 0.03 |
| Month 12 | N/A | -0.35 | -0.15 | -0.42 |

Table S5: Falls and syncope

|  | Conservative management | Conservative management plus fludrocortisone | Conservative management plus midodrine |
| --- | --- | --- | --- |
| **Falls** | **(N = 0)** | **(N = 4)** | **(N = 6)** |
| Number of events |  |  |  |
| 0 | N/A | 3 (75%) | 1 (17%) |
| 1 | N/A | 0 (0%) | 0 (0%) |
| 2 | N/A | 1 (25%) | 2 (33%) |
| ≥3 | N/A | 0 (0%) | 3 (50%) |
| Event rate |  |  |  |
| Total number of events | N/A | 2 | 46 |
| Total observation time (years) | N/A | 4.2 | 5.4 |
| Incidence rate (95% CI) | N/A | 0.5 (0.1, 1.7) | 8.6 (6.3, 11.4) |
| Fallers (≥ 1 fall) | N/A | 1 (25%) | 5 (83%) |
| Time to first fall |  |  |  |
| < 3months | N/A | 1 (100%) | 4 (80%) |
| 3-6 months | N/A | 0 (0%) | 1 (20%) |
| **Syncopal events** | **(N = 0)** | **(N = 4)** | **(N = 6)** |
| Number of events |  |  |  |
| 0 | N/A | 4 (100%) | 5 (83%) |
| ≥1 | N/A | 0 (0%) | 1 (17%) |
| Total number of events | N/A | 0 | 9 |

Table S6. Falls-related injuries

| Subject ID | Randomised group | Treatment group at time of AE onset* | Weeks from randomisation to AE onset | AE | Serious | Severity | Causality | Outcome |
| --- | --- | --- | --- | --- | --- | --- | --- | --- |
| 106 | Conservative management |  | 29 | Multiple fractures | Yes | Severe | Unrelated | Recovered with sequelae |
| 49 | Conservative management plus fludrocortisone |  | 12 | Ligament sprain | No | Mild | Unrelated | Recovered |
|  |  |  | 12 | Head injury | No | Mild | Unrelated | Recovered |
| 98 | Conservative management plus midodrine | Conservative management plus fludrocortisone | 47 | Back injury | No | Mild | Unrelated | Recovered |

**if different from randomised group*

Table S7: Adverse events

|  | Conservative management | Conservative management plus fludrocortisone | Conservative management plus midodrine |
| --- | --- | --- | --- |
| **Adverse events (AEs)*** | **(N = 3)** | **(N = 5)** | **(N = 6)** |
| No. of AEs reported per participant |  |  |  |
| 0 | 1 (33%) | 1 (20%) | 2 (33%) |
| 1 | 1 (33%) | 1 (20%) | 2 (33%) |
| 2-3 | 1 (33%) | 2 (40%) | 2 (33%) |
| >3 | 0 (0%) | 1 (20%) | 0 (0%) |
| Median (Range) | 1 (0, 3) | 2 (0, 11) | 1 (0, 2) |
| Worst grade AE reported per participant |  |  |  |
| No AE | 1 (33%) | 1 (20%) | 2 (33%) |
| Mild | 0 (0%) | 2 (40%) | 4 (67%) |
| Moderate | 0 (0%) | 0 (0%) | 0 (0%) |
| Severe | 2 (67%) | 2 (40%) | 0 (0%) |
| **Adverse reactions (ARs)*** | **(N = 3)** | **(N = 5)** | **(N = 6)** |
| No. of ARs reported per participant |  |  |  |
| 0 | 3 (100%) | 5 (100%) | 3 (50%) |
| 1 | 0 (0%) | 0 (0%) | 1 (17%) |
| 2 | 0 (0%) | 0 (0%) | 2 (33%) |
| Worst grade AR reported per participant |  |  |  |
| No AR | 3 (100%) | 5 (100%) | 3 (50%) |
| Mild | 0 (0%) | 0 (0%) | 3 (50%) |
| **Serious adverse events (SAEs)*** | **(N = 3)** | **(N = 5)** | **(N = 6)** |
| Number of SAEs reported per participant |  |  |  |
| 0 | 1 (33%) | 3 (40%) | 6 (100%) |
| 1 | 1 (33%) | 1 (20%) | 0 (0%) |
| ≥2 | 1 (33%) | 1 (20%) | 0 (0%) |
| Range | 0, 3 | 0, 4 | 0, 0 |

*Data reported out of number exposed to each treatment, allowing for crossover between interventions

Table S8: Adverse events and severity per treatment arm

|  | | Number (%) of participants affected | | |
| --- | --- | --- | --- | --- |
|  |  | Conservative management | Conservative management plus fludrocortisone | Conservative management plus midodrine |
| Participants exposed | | 3 | 5 | 6 |
| Abscess | Total | 0 (0%) | 1 (20%) | 0 (0%) |
|  | Mild | 0 (0%) | 1 (20%) | 0 (0%) |
|  | Moderate | 0 (0%) | 0 (0%) | 0 (0%) |
|  | Severe | 0 (0%) | 0 (0%) | 0 (0%) |
| Atrioventricular block | Total | 1 (33%) | 0 (0%) | 0 (0%) |
|  | Mild | 0 (0%) | 0 (0%) | 0 (0%) |
|  | Moderate | 0 (0%) | 0 (0%) | 0 (0%) |
|  | Severe | 1 (33%) | 0 (0%) | 0 (0%) |
| Back injury | Total | 0 (0%) | 1 (20%) | 0 (0%) |
|  | Mild | 0 (0%) | 1 (20%) | 0 (0%) |
|  | Moderate | 0 (0%) | 0 (0%) | 0 (0%) |
|  | Severe | 0 (0%) | 0 (0%) | 0 (0%) |
| Bradycardia | Total | 0 (0%) | 0 (0%) | 1 (17%) |
|  | Mild | 0 (0%) | 0 (0%) | 1 (17%) |
|  | Moderate | 0 (0%) | 0 (0%) | 0 (0%) |
|  | Severe | 0 (0%) | 0 (0%) | 0 (0%) |
| COVID-19 | Total | 1 (33%) | 2 (40%) | 0 (0%) |
|  | Mild | 0 (0%) | 2 (40%) | 0 (0%) |
|  | Moderate | 0 (0%) | 0 (0%) | 0 (0%) |
|  | Severe | 1 (33%) | 0 (0%) | 0 (0%) |
| Cardiac arrest | Total | 1 (33%) | 0 (0%) | 0 (0%) |
|  | Mild | 0 (0%) | 0 (0%) | 0 (0%) |
|  | Moderate | 0 (0%) | 0 (0%) | 0 (0%) |
|  | Severe | 1 (33%) | 0 (0%) | 0 (0%) |
| Cellulitis | Total | 0 (0%) | 1 (20%) | 0 (0%) |
|  | Mild | 0 (0%) | 1 (20%) | 0 (0%) |
|  | Moderate | 0 (0%) | 0 (0%) | 0 (0%) |
|  | Severe | 0 (0%) | 0 (0%) | 0 (0%) |
| Dyspnoea | Total | 0 (0%) | 1 (20%) | 0 (0%) |
|  | Mild | 0 (0%) | 0 (0%) | 0 (0%) |
|  | Moderate | 0 (0%) | 0 (0%) | 0 (0%) |
|  | Severe | 0 (0%) | 1 (20%) | 0 (0%) |
| HIV infection | Total | 0 (0%) | 1 (20%) | 0 (0%) |
|  | Mild | 0 (0%) | 0 (0%) | 0 (0%) |
|  | Moderate | 0 (0%) | 0 (0%) | 0 (0%) |
|  | Severe | 0 (0%) | 1 (20%) | 0 (0%) |
| Head injury | Total | 0 (0%) | 1 (20%) | 0 (0%) |
|  | Mild | 0 (0%) | 1 (20%) | 0 (0%) |
|  | Moderate | 0 (0%) | 0 (0%) | 0 (0%) |
|  | Severe | 0 (0%) | 0 (0%) | 0 (0%) |
| Headache | Total | 0 (0%) | 0 (0%) | 1 (17%) |
|  | Mild | 0 (0%) | 0 (0%) | 1 (17%) |
|  | Moderate | 0 (0%) | 0 (0%) | 0 (0%) |
|  | Severe | 0 (0%) | 0 (0%) | 0 (0%) |
| Lethargy | Total | 0 (0%) | 0 (0%) | 1 (17%) |
|  | Mild | 0 (0%) | 0 (0%) | 1 (17%) |
|  | Moderate | 0 (0%) | 0 (0%) | 0 (0%) |
|  | Severe | 0 (0%) | 0 (0%) | 0 (0%) |
| Ligament sprain | Total | 0 (0%) | 1 (20%) | 0 (0%) |
|  | Mild | 0 (0%) | 1 (20%) | 0 (0%) |
|  | Moderate | 0 (0%) | 0 (0%) | 0 (0%) |
|  | Severe | 0 (0%) | 0 (0%) | 0 (0%) |
| Lower respiratory tract infection | Total | 0 (0%) | 1 (20%) | 1 (17%) |
|  | Mild | 0 (0%) | 1 (20%) | 1 (17%) |
|  | Moderate | 0 (0%) | 0 (0%) | 0 (0%) |
|  | Severe | 0 (0%) | 0 (0%) | 0 (0%) |
| Multiple fractures | Total | 1 (33%) | 0 (0%) | 0 (0%) |
|  | Mild | 0 (0%) | 0 (0%) | 0 (0%) |
|  | Moderate | 0 (0%) | 0 (0%) | 0 (0%) |
|  | Severe | 1 (33%) | 0 (0%) | 0 (0%) |
| Muscle Spasms | Total | 0 (0%) | 0 (0%) | 1 (17%) |
|  | Mild | 0 (0%) | 0 (0%) | 1 (17%) |
|  | Moderate | 0 (0%) | 0 (0%) | 0 (0%) |
|  | Severe | 0 (0%) | 0 (0%) | 0 (0%) |
| Pneumocystis jirovecii pneumonia | Total | 0 (0%) | 1 (20%) | 0 (0%) |
|  | Mild | 0 (0%) | 1 (20%) | 0 (0%) |
|  | Moderate | 0 (0%) | 0 (0%) | 0 (0%) |
|  | Severe | 0 (0%) | 0 (0%) | 0 (0%) |
| Prostate cancer | Total | 0 (0%) | 1 (20%) | 0 (0%) |
|  | Mild | 0 (0%) | 1 (20%) | 0 (0%) |
|  | Moderate | 0 (0%) | 0 (0%) | 0 (0%) |
|  | Severe | 0 (0%) | 0 (0%) | 0 (0%) |
| Staphylococcal infection | Total | 0 (0%) | 1 (20%) | 0 (0%) |
|  | Mild | 0 (0%) | 1 (20%) | 0 (0%) |
|  | Moderate | 0 (0%) | 0 (0%) | 0 (0%) |
|  | Severe | 0 (0%) | 0 (0%) | 0 (0%) |
| Urinary infection | Total | 0 (0%) | 1 (20%) | 0 (0%) |
|  | Mild | 0 (0%) | 1 (20%) | 0 (0%) |
|  | Moderate | 0 (0%) | 0 (0%) | 0 (0%) |
|  | Severe | 0 (0%) | 0 (0%) | 0 (0%) |
| Urinary tract infection | Total | 0 (0%) | 1 (20%) | 0 (0%) |
|  | Mild | 0 (0%) | 1 (20%) | 0 (0%) |
|  | Moderate | 0 (0%) | 0 (0%) | 0 (0%) |
|  | Severe | 0 (0%) | 0 (0%) | 0 (0%) |
| Vomiting | Total | 0 (0%) | 0 (0%) | 1 (17%) |
|  | Mild | 0 (0%) | 0 (0%) | 1 (17%) |
|  | Moderate | 0 (0%) | 0 (0%) | 0 (0%) |
|  | Severe | 0 (0%) | 0 (0%) | 0 (0%) |

Table S9: Adverse reactions

| Subject ID | Treatment group | Days from randomisation to AE onset | AE | Severity | Causality |
| --- | --- | --- | --- | --- | --- |
| 98 | Conservative management plus midodrine | 19 | Vomiting | Mild | Possibly related |
|  |  | 19 | Headache | Mild | Possibly related |
| 110 | Conservative management plus midodrine | 5 | Lethargy | Mild | Possibly related |
|  |  | 5 | Bradycardia | Mild | Possibly related |
| 31 | Conservative management plus midodrine | 6 | Muscle Spasms | Mild | Possibly related |

Table S10: Serious adverse events

| Subject ID | Treatment group | AE | Injury as a result of a fall | Severity | Causality | Serious criteria | Outcome |
| --- | --- | --- | --- | --- | --- | --- | --- |
| 25 | Conservative management plus fludrocortisone | Dyspnoea | No | Severe | Unlikely to be related | Hospitalisation/prolongation of hospital stay | Recovered |
| 106 | Conservative management | Atrioventricular block | No | Severe | Unrelated | Hospitalisation/prolongation of hospital stay | Condition improved |
|  |  | COVID-19 | No | Severe | Unrelated | Hospitalisation/prolongation of hospital stay | Recovered |
|  |  | Multiple fractures | Yes | Severe | Unrelated | Hospitalisation/prolongation of hospital stay | Recovered with sequelae |
| 35 | Conservative management | Cardiac arrest | No | Severe | Unrelated | Resulted in death | Participant died |
| 49 | Conservative management plus fludrocortisone | HIV infection | No | Severe | Unrelated | Hospitalisation/prolongation of hospital stay | Condition improved |
|  |  | Pneumocystis jirovecii pneumonia | No | Mild | Unrelated | Hospitalisation/prolongation of hospital stay | Recovered |
|  |  | COVID-19 | No | Mild | Unrelated | Hospitalisation/prolongation of hospital stay | Recovered |
|  |  | Urinary infection | No | Mild | Unrelated | Hospitalisation/prolongation of hospital stay | Recovered |

Table S11: Frequency of GP Appointments

|  | Conservative management | Conservative management plus fludrocortisone | Conservative management plus midodrine | Overall |
| --- | --- | --- | --- | --- |
| **Baseline** | **Exp = 3** | **Exp = 4** | **Exp = 6** | **Exp = 13** |
| 0 | 3 | 2 | 5 | 10 |
| 1 | 0 | 0 | 1 | 1 |
| 4 | 0 | 2 | 0 | 2 |
| Mean (SD) | 0 (0) | 2.00 (2.31) | 0.17 (0.41) | 0.69 (1.49) |
| Median (Range) | 0 (0) | 2 (0 -4) | 0 (0-1) | 0 (0-4) |
| **Month 3** | **Exp = 1** | **Exp = 4** | **Exp = 6** | **Exp = 11** |
| 0 | 1 | 3 | 4 | 8 |
| 1 | 0 | 1 | 2 | 3 |
| Mean (SD) | 0 (0) | 0.25 (0.5) | 0.33 (0.52) | 0.270.47) |
| Median (Range) | 0 (0) | 0 (0-1) | 0 (0-1) | 0 (0-1) |
| **Month 6** | **Exp = 0** | **Exp = 4** | **Exp = 6** | **Exp = 10** |
| 0 |  | 3 | 5 | 8 |
| 1 |  | 0 | 1 | 1 |
| 3 |  | 1 | 0 | 1 |
| Mean (SD) |  | 0.75 (1.50) \| | 0.17 (0.41) | 0.40 (0.97) |
| Median (Range) |  | 0 (0-1) | 0 (0-1) | 0(0-6) |
| **Month 12** | **Exp = 0** | **Exp = 4** | **Exp = 5** | **Exp = 9** |
| 1 |  | 2 | 2 | 4 |
| 2 |  | 1 | 2 | 3 |
| 3 |  | 0 | 1 | 1 |
| 12 |  | 1 | 0 | 1 |
| Mean (SD) |  | 3.25 (5.85) | 1 (1.22) | (2-3.87) |
| Median (Range) |  | 0.5 (0-12) | 1 (0-3) | 1 (0-12) |

Table S12: Frequency of Nurse Appointments

|  | Conservative management | Conservative management plus fludrocortisone | Conservative management plus midodrine | Overall |
| --- | --- | --- | --- | --- |
| **Baseline** | **Exp = 3** | **Exp = 4** | **Exp = 6** | **Exp = 13** |
| 0 | 2 | 3 | 2 | 7 |
| 1 | 1 | 1 | 2 | 4 |
| 2 | 0 | 0 | 1 | 1 |
| 3 | 0 | 0 | 1 | 1 |
| Mean (SD) | 0.33 (0.58) | 0.25 (0.50) | 1.17 (1.17) | 0.69 \| (0.95) |
| Median (Range) | 0 (0-1) | 0 (0-1) | 1 (0-3) | 0 (0-3) |
| **Month 3** | **Exp = 1** | **Exp = 4** | **Exp = 6** | **Exp = 11** |
| 0 | 1 | 3 | 4 | 8 |
| 1 | 0 | 1 | 1 | 2 |
| 2 | 0 | 0 | 1 | 1 |
| Mean (SD) | 0 (0) | 0.25 (0.5) | 0.50 (0.84) | 0.36 (0.67) |
| Median (Range) | 0(0) | 0 (0-1) | 0 (0-2) | 0 (0-2) |
| **Month 6** | **Exp = 0** | **Exp = 4** | **Exp = 6** | **Exp = 10** |
| 0 |  | 2 | 5 | 7 |
| 1 |  | 2 | 1 | 3 |
| Mean (SD) |  | 0.50 (0.58) | 0.17 (0.41) \| | 0.3 (0.48) |
| Median (Range) |  | 0.5 (0-1) | 0 (0-1) | 0 (0-1) |
| **Month 12** | **Exp = 0** | **Exp = 4** | **Exp = 5** | **Exp = 9** |
|  |  | 2 | 3 | 5 |
|  |  | 2 | 1 | 3 |
|  |  | 0 | 1 | 1 |
| Mean (SD) |  | 0.50 (0.58) | 0.80 (1.30) | 0.67 (1.00) |
| Median (Range) |  | 0.5 (0-1) | 0 (0-3) | 0 (0-3) |

Table S13: Frequency of Telephone GP Appointments

|  | Conservative management | Conservative management plus fludrocortisone | Conservative management plus midodrine | Overall |
| --- | --- | --- | --- | --- |
| **Baseline** | **Exp = 3** | **Exp = 4** | **Exp = 6** | **Exp = 13** |
| 0 | 2 | 2 | 5 | 8 |
| 1 | 1 | 2 | 1 | 4 |
| 2 | 1 | 0 | 0 | 1 |
| Mean (SD) | 1 (1) | 0.5 (0.58) | 0.17 (0.41) | 0.46 (0.66) |
| Median (Range) | 1 (0-2) | 0.5 (0-1) | 0 (0-1) | 0 (0-2) |
| **Month 3** | **Exp = 1** | **Exp = 4** | **Exp = 6** | **Exp = 11** |
| 0 | 1 | 3 | 4 | 8 |
| 1 | 0 | 1 | 1 | 2 |
| 2 | 0 | 0 | 1 | 1 |
| Mean (SD) | 0 (0) | 1.00 (1.15) | 0.33 (0.52) | 0.55 (0.82) |
| Median (Range) | 0 (0) | 1 (0-2) | 0 (0-1) | 0 (0-2) |
| **Month 6** | **Exp = 0** | **Exp = 4** | **Exp = 6** | **Exp = 10** |
| 0 |  | 3 | 5 | 8 |
| 1 |  | 0 | 1 | 1 |
| 3 |  | 1 | 0 | 1 |
| Mean (SD) |  | 0.75 (1.5) | 0.17 (0.41) | 0.4 (0.97) |
| Median (Range) |  | 0 (0-3) | 0 (0-1) | 0 (0-3) |
| **Month 12** | **Exp = 0** | **Exp = 4** | **Exp = 5** | **Exp = 9** |
| 0 |  | 4 | 4 | 6 |
| 1 |  | 0 | 1 | 3 |
| Mean (SD) |  | 0 (0) | 0.2 (0.45) | 0.11(0.33) |
| Median (Range) |  | 0 (0) | 0 (0-1) | 0 (0.-1) |

Table S14: Frequency of Telephone Nurse Appointments

|  | Conservative management | Conservative management plus fludrocortisone | Conservative management plus midodrine | Overall |
| --- | --- | --- | --- | --- |
| **Baseline** | **Exp = 3** | **Exp = 4** | **Exp = 6** | **Exp = 13** |
| 0 | 3 | 4 | 5 | 12 |
| 1 | 0 | 0 | 1 | 1 |
| Mean (SD) | 0 (0) | 0 (0) | 0.17 (0.41) | 0.08 (0.28) |
| Median (Range) | 0 (0) | 0 (0) | 0 (0-1) | 0 (0-1) |
| **Month 3** | **Exp = 1** | **Exp = 4** | **Exp = 6** | **Exp = 11** |
| 0 | 0 | 4 | 5 | 9 |
| 1 | 1 | 0 | 1 | 2 |
| Mean (SD) | 1 (0) | 0 (0) | 0.17 (0.41) | 0.18 (0.40) |
| Median (Range) | 1 (1 | 0 (0) | 0 (0-1) | 0 (0-1) |
| **Month 6** | **Exp = 0** | **Exp = 4** | **Exp = 6** | **Exp = 10** |
|  |  | 4 | 5 | 9 |
|  |  | 0 | 1 | 1 |
| Mean (SD) |  | 0 (0) | 0.17 (0.41) | 0.10 (0.32) |
| Median (Range) |  | 0 (0-1) | 0 (0-1) | 0 (0-1) |
| **Month 12** | **Exp = 0** | **Exp = 4** | **Exp = 5** | **Exp = 9** |
| 0 |  | 4 | 3 | 7 |
| 1 |  | 0 | 1 | 1 |
| 2 |  | 0 | 1 | `1 |
| Mean (SD) |  | 0 (0) | 0.60 (0.89) | 0.33 (0.71) |
| Median (Range) |  | 0(0) | 0 (0-2) | 0 (0-2) |

Table S15: Frequency of doctor visits at home

|  | Conservative management | Conservative management plus fludrocortisone | Conservative management plus midodrine | Overall |
| --- | --- | --- | --- | --- |
| **Baseline** | **Exp = 3** | **Exp = 4** | **Exp = 6** | **Exp = 13** |
| 0 | 2 | 4 | 5 | 11 |
| 1 | 0 | 0 | 1 | 1 |
| 2 | 1 | 0 | 0 | 1 |
| Mean (SD) | 0.67 (1.15) | 0 (N/A) | 0.17 (0.41) | 0.23 (0.60) |
| Median (Range) | 0 (0-2) | 0 (N/A) | 0 (0-1) | 0 (0-2) |
| **Month 3** | **Exp = 1** | **Exp = 4** | **Exp = 6** | **Exp = 11** |
| 0 | 1 | 2 | 6 | 9 |
| 1 | 0 | 2 | 0 | 2 |
| Mean (SD) | 0 (0) | 0.5(0.58) | 0 (0) | 0.18 (0.40) |
| Median (Range) | 0 (0) | 0.5 (0-1) | 0 (0-1) | 0 (0-1) |
| **Month 6** | **Exp = 0** | **Exp = 4** | **Exp = 6** | **Exp = 10** |
| 0 |  | 3 | 6 | 9 |
| 1 |  | 1 | 0 | 1 |
| Mean (SD) |  | 0.25 (0.5) | 0 (0) | (0.1-0.32) |
| Median (Range) |  | 0 (0-1) | 0 (0) | 0 (0-1) |
| **Month 12** | **Exp = 0** | **Exp = 4** | **Exp = 5** | **Exp = 9** |
| 0 |  | 4 | 4 | 8 |
| 2 |  | 0 | 1 | 1 |
| Mean (SD) |  | 0 (0) | 0.40 (0.89) | 0.22 (0.67) |
| Median (Range) |  | 0 (0) | 0 (0 -2) | 0 (0-2) |

Table S16: Frequency Walk in Centre appointments

|  | Conservative management | Conservative management plus fludrocortisone | Conservative management plus midodrine | Overall |
| --- | --- | --- | --- | --- |
| **Baseline** | **Exp = 3** | **Exp = 4** | **Exp = 6** | **Exp = 13** |
| 0 | 3 | 4 | 6 | 13 |
| Mean (SD) | 0 (0) | 0 (0) | 0 (0) | 0 (0) |
| Median (Range) | 0 (0) | 0 (0) | 0 (0) | 0 (0) |
| **Month 3** | **Exp = 1** | **Exp = 4** | **Exp = 6** | **Exp = 11** |
| 0 | 0 | 0 | 2 | 5 |
| Mean (SD) | 0 (0) | 0 (0) | 0 (0) | 0 (0) |
| Median (Range) | 0 (0) | 0 (0) | 0 (0) | 0 (0) |
| **Month 6** | **Exp = 0** | **Exp = 4** | **Exp = 6** | **Exp = 10** |
| 0 |  | 3 | 5 | 8 |
| 1 |  | 1 | 1 | 2 |
| Mean (SD) |  | 0.25 (0.50) | 0.17 (0.41) | 0.20 (0.42) |
| Median (Range) |  | 0 (0-1) | 0 (0-1) | 0 (0-1) |
| **Month 12** | **Exp = 0** | **Exp = 4** | **Exp = 5** | **Exp = 9** |
| 0 |  | 4 | 4 | 8 |
| 1 |  | 0 | 1 | 1 |
| Mean (SD) |  | 0 (0) | 0.40 (0.89) | 0.11 \| (0.33) |
| Median (Range) |  | 0 (0) | 0 (0-2) | 0 (0-1) |

Table S17: Frequency of ambulance services

|  | Conservative management | Conservative management plus fludrocortisone | Conservative management plus midodrine | Overall |
| --- | --- | --- | --- | --- |
| **Baseline** | **Exp = 3** | **Exp = 4** | **Exp = 6** | **Exp = 13** |
| 0 | 2 | 3 | 4 | 9 |
| 1 | 0 | 1 | 1 | 2 |
| 2 | 1 | 0 | 1 | 2 |
| Mean (SD) | 0.67 (1.15) | 0.25 (0.50) | 0.50 (0.84) | 0.46 (0.78) |
| Median (Range) | 0 (0-2) | 0 (0-1) | 0(0-2) | 0 (0-2) |
| **Month 3** | **Exp = 1** | **Exp = 4** | **Exp = 6** | **Exp = 11** |
| 0 | 0 | 2 | 5 | 7 |
| 1 | 1 | 2 | 1 | 4 |
| Mean (SD) | 1(0) | 0.50 (0.58) | 0.17 (0.41) | 0.36 (0.50) |
| Median (Range) | 1 (0) | 0.5 (0-1) | 0 (0-1) | 0 (0-1) |
| **Month 6** | **Exp = 0** | **Exp = 4** | **Exp = 6** | **Exp = 10** |
| 0 |  | 4 | 5 | 9 |
| 1 |  | 0 | 1 | 1 |
| Mean (SD) |  | 0(0) | 0.17 (0.41) | 0.1 (0.32) |
| Median (Range) |  | 0 (0) | 0 (0-1) | 0 (0-1) |
| **Month 12** | **Exp = 0** | **Exp = 4** | **Exp = 5** | **Exp = 9** |
| 0 |  | 3 | 5 | 8 |
| 1 |  | 1 | 0 | 1 |
| Mean (SD) |  | 0.25 (0.50) | 0 (0) | 0.11 (0.33) |
| Median (Range) |  | 0 (0-1) | 0 (0) | 0 (0-1) |

Table S18: Frequency of Accident and Emergency services

|  | Conservative management | Conservative management plus fludrocortisone | Conservative management plus midodrine | Overall |
| --- | --- | --- | --- | --- |
| **Baseline** | **Exp = 3** | **Exp = 4** | **Exp = 6** | **Exp = 13** |
| 0 | 2 | 2 | 4 | 8 |
| 1 | 1 | 1 | 1 | 3 |
| 2 | 0 | 1 | 1 | 2 |
| Mean (SD) | 0.33 (0.75) | 0.75 (0.96) | 0.5 (0.84) | 0.54 (0.78) |
| Median (Range) | 0 (0-1) | 0.5 (0-2) | 0 (0-2) | 0 (0-2) |
| **Month 3** | **Exp = 1** | **Exp = 4** | **Exp = 6** | **Exp = 11** |
| 0 | 0 | 2 | 5 | 7 |
| 1 | 1 | 2 | 1 | 4 |
| Mean (SD) | 1 | 0.50 (0.58) | 0.17 (0.41) | 0.36 (0.5) |
| Median (Range) | 1 | 0.5 (0-1) | 0 (0-1) | 0 (0-1) |
| **Month 6** | **Exp = 0** | **Exp = 4** | **Exp = 6** | **Exp = 10** |
| 0 |  | 4 | 5 | 9 |
| 1 |  | 0 | 1 | 1 |
| Mean (SD) |  | 0 (0) | 0.17 (0.41) | 0.1 (0.32) |
| Median (Range) |  | 0 (0) | 0 (0-1) | 0 (0-1) |
| **Month 12** | **Exp = 0** | **Exp = 4** | **Exp = 5** | **Exp = 9** |
| 0 |  | 3 | 4 | 7 |
| 1 |  | 1 | 0 | 1 |
| 2 |  | 0 | 1 | 1 |
| Mean (SD) |  | 0.25 (0.50) | 0.4 (0.89) | 0.33 (0.71) |
| Median (Range) |  | 0 (0-1) | 0 (0-2) | 0 (0-2) |

Table S19: Frequency of other appointments in hospital

|  | Conservative management | Conservative management plus fludrocortisone | Conservative management plus midodrine | Overall |
| --- | --- | --- | --- | --- |
| **Baseline** | **Exp = 3** | **Exp = 4** | **Exp = 6** | **Exp = 13** |
| Yes | 2 | 0 | 2 | 4 |
| No | 1 | 4 | 4 | 9 |
| **Month 3** | **Exp = 1** | **Exp = 4** | **Exp = 6** | **Exp = 11** |
| Yes | 0 | 0 | 3 | 3 |
| No | 1 | 4 | 3 | 8 |
| **Month 6** | **Exp = 0** | **Exp = 4** | **Exp = 6** | **Exp = 10** |
| Yes |  | 1 | 3 | 4 |
| No |  | 3 | 3 | 6 |
| **Month 12** | **Exp = 0** | **Exp = 4** | **Exp = 5** | **Exp = 9** |
| Yes |  | 2 | 1 | 3 |
| No |  | 2 | 4 | 6 |

Table S20: Frequency of private appointments

|  | Conservative management | Conservative management plus fludrocortisone | Conservative management plus midodrine | Overall |
| --- | --- | --- | --- | --- |
| **Baseline** | **Exp = 3** | **Exp = 4** | **Exp = 6** | **Exp = 13** |
| No | 3 | 4 | 6 | 13 |
| **Month 3** | **Exp = 1** | **Exp = 4** | **Exp = 6** | **Exp = 11** |
| No | 1 | 4 | 6 | 11 |
| **Month 6** | **Exp = 0** | **Exp = 4** | **Exp = 6** | **Exp = 10** |
| No |  | 4 | 6 | 10 |
| **Month 12** | **Exp = 0** | **Exp = 4** | **Exp = 5** | **Exp = 9** |
| No |  | 4 | 5 | 9 |

Table S21: Frequency of personal social service

|  | Conservative management | Conservative management plus fludrocortisone | Conservative management plus midodrine | Overall |
| --- | --- | --- | --- | --- |
| **Baseline** | **Exp = 3** | **Exp = 4** | **Exp = 6** | **Exp = 13** |
| Yes | 1 | 0 | 2 | 3 |
| No | 2 | 4 | 4 | 10 |
| **Month 3** | **Exp = 1** | **Exp = 4** | **Exp = 6** | **Exp = 11** |
| Yes | 0 | 0 | 1 | 1 |
| No | 1 | 4 | 5 | 10 |
| **Month 6** | **Exp = 0** | **Exp = 4** | **Exp = 6** | **Exp = 10** |
| Yes |  | 0 | 1 | 1 |
| No |  | 4 | 5 | 9 |
| **Month 12** | **Exp = 0** | **Exp = 4** | **Exp = 5** | **Exp = 9** |
| Yes |  | 1 | 0 | 1 |
| No |  | 3 | 5 | 8 |

Table S22: Frequency of Personal Guarantee Credit

|  | Conservative management | Conservative management plus fludrocortisone | Conservative management plus midodrine | Overall |
| --- | --- | --- | --- | --- |
| **Baseline** | **Exp = 3** | **Exp = 4** | **Exp = 6** | **Exp = 13** |
| Yes | 0 | 0 | 1 | 1 |
| No | 3 | 4 | 5 | 12 |
| **Month 3** | **Exp = 1** | **Exp = 4** | **Exp = 6** | **Exp = 11** |
| Yes | 0 | 0 | 1 | 1 |
| No | 1 | 4 | 5 | 10 |
| **Month 6** | **Exp = 0** | **Exp = 4** | **Exp = 6** | **Exp = 10** |
| Yes |  | 0 | 2 | 2 |
| No |  | 4 | 4 | 8 |
| **Month 12** | **Exp = 0** | **Exp = 4** | **Exp = 5** | **Exp = 9** |
| Yes |  | 0 | 1 | 1 |
| No |  | 4 | 4 | 8 |

Table S23: Frequency of Attendance Allowance

|  | Conservative management | Conservative management plus fludrocortisone | Conservative management plus midodrine | Overall |
| --- | --- | --- | --- | --- |
| **Baseline** | **Exp = 3** | **Exp = 4** | **Exp = 6** | **Exp = 13** |
| Yes | 2 | 0 | 1 | 3 |
| No | 1 | 4 | 5 | 10 |
| **Month 3** | **Exp = 1** | **Exp = 4** | **Exp = 6** | **Exp = 11** |
| Yes | 0 | 1 | 2 | 3 |
| No | 1 | 3 | 4 | 8 |
| **Month 6** | **Exp = 0** | **Exp = 4** | **Exp = 6** | **Exp = 10** |
| Yes |  | 1 | 1 | 2 |
| No |  | 3 | 4 | 7 |
| Missing |  | 0 | 1 | 1 |
| **Month 12** | **Exp = 0** | **Exp = 4** | **Exp = 5** | **Exp = 9** |
| Yes |  | 1 | 1 | 2 |
| No |  | 3 | 4 | 7 |

Table S24: Frequency of Personal Independence Payment (Daily Living)

|  | Conservative management | Conservative management plus fludrocortisone | Conservative management plus midodrine | Overall |
| --- | --- | --- | --- | --- |
| **Baseline** | **Exp = 3** | **Exp = 4** | **Exp = 6** | **Exp = 13** |
| Yes | 0 | 1 | 4 | 5 |
| No | 3 | 3 | 2 | 8 |
| **Month 3** | **Exp = 1** | **Exp = 4** | **Exp = 6** | **Exp = 11** |
| Yes | 0 | 1 | 2 | 3 |
| No | 1 | 3 | 4 | 8 |
| **Month 6** | **Exp = 0** | **Exp = 4** | **Exp = 6** | **Exp = 10** |
| Yes |  | 1 | 2 | 3 |
| No |  | 3 | 3 | 6 |
| Missing |  | 0 | 1 | 1 |
| **Month 12** | **Exp = 0** | **Exp = 4** | **Exp = 5** | **Exp = 9** |
| Yes |  | 1 | 2 | 3 |
| No |  | 3 | 3 | 6 |

Table S25: Frequency of Personal independent payment (Mobility)

|  | Conservative management | Conservative management plus fludrocortisone | Conservative management plus midodrine | Overall |
| --- | --- | --- | --- | --- |
| **Baseline** | **Exp = 3** | **Exp = 4** | **Exp = 6** | **Exp = 13** |
| Yes | 0 | 1 | 4 | 5 |
| No | 3 | 3 | 2 | 8 |
| **Month 3** | **Exp = 1** | **Exp = 4** | **Exp = 6** | **Exp = 11** |
| Yes | 0 | 1 | 2 | 3 |
| No | 1 | 3 | 4 | 8 |
| **Month 6** | **Exp = 0** | **Exp = 4** | **Exp = 6** | **Exp = 10** |
| Yes |  | 1 | 2 | 3 |
| No |  | 3 | 3 | 6 |
| Missing |  | 0 | 1 | 1 |
| **Month 12** | **Exp = 0** | **Exp = 4** | **Exp = 5** | **Exp = 9** |
| Yes |  | 1 | 2 | 3 |
| No |  | 3 | 3 | 6 |

Table S26: Frequency of Employment and Support Allowance

|  | Conservative management | Conservative management plus fludrocortisone | Conservative management plus midodrine | Overall |
| --- | --- | --- | --- | --- |
| **Baseline** | **Exp = 3** | **Exp = 4** | **Exp = 6** | **Exp = 13** |
| No | 3 | 4 | 6 | 13 |
| **Month 3** | **Exp = 1** | **Exp = 4** | **Exp = 6** | **Exp = 11** |
| No | 1 | 4 | 6 | 11 |
| **Month 6** | **Exp = 0** | **Exp = 4** | **Exp = 6** | **Exp = 10** |
| No |  | 4 | 5 | 9 |
| Missing |  | 0 | 1 | 1 |
| **Month 12** | **Exp = 0** | **Exp = 4** | **Exp = 5** | **Exp = 9** |
| No |  | 4 | 5 | 9 |

Table S27: Frequency of Universal Credit

|  | Conservative management | Conservative management plus fludrocortisone | Conservative management plus midodrine | Overall |
| --- | --- | --- | --- | --- |
| **Baseline** | **Exp = 3** | **Exp = 4** | **Exp = 6** | **Exp = 13** |
| Yes | 1 | 0 | 0 | 1 |
| No | 2 | 4 | 6 | 12 |
| **Month 3** | **Exp = 1** | **Exp = 4** | **Exp = 6** | **Exp = 11** |
| No | 1 | 4 | 6 | 11 |
| **Month 6** | **Exp = 0** | **Exp = 4** | **Exp = 6** | **Exp = 10** |
| Yes |  | 4 | 5 | 9 |
| Missing |  | 0 | 1 | 1 |
| **Month 12** | **Exp = 0** | **Exp = 4** | **Exp = 5** | **Exp = 9** |
| No |  | 4 | 5 | 9 |

Table S28: Frequency of Carer’s Allowance

|  | Conservative management | Conservative management plus fludrocortisone | Conservative management plus midodrine | Overall |
| --- | --- | --- | --- | --- |
| **Baseline** | **Exp = 3** | **Exp = 4** | **Exp = 6** | **Exp = 13** |
| No | 3 | 4 | 6 | 13 |
| **Month 3** | **Exp = 1** | **Exp = 4** | **Exp = 6** | **Exp = 11** |
| No | 1 | 4 | 6 | 11 |
| **Month 6** | **Exp = 0** | **Exp = 4** | **Exp = 6** | **Exp = 10** |
| Yes |  | 4 | 5 | 9 |
| Missing |  | 0 | 1 | 1 |
| **Month 12** | **Exp = 0** | **Exp = 4** | **Exp = 5** | **Exp = 9** |

Table S29: Frequency Social Worker Service

|  | Conservative management | Conservative management plus fludrocortisone | Conservative management plus midodrine | Overall |
| --- | --- | --- | --- | --- |
| **Baseline** | **Exp = 3** | **Exp = 4** | **Exp = 6** | **Exp = 13** |
| Yes | 1 | 1 | 1 | 3 |
| No | 2 | 3 | 5 | 10 |
| **Month 3** | **Exp = 1** | **Exp = 4** | **Exp = 6** | **Exp = 11** |
| No | 1 | 4 | 6 | 11 |
| **Month 6** | **Exp = 0** | **Exp = 4** | **Exp = 6** | **Exp = 10** |
| No |  | 4 | 6 | 10 |
| **Month 12** | **Exp = 0** | **Exp = 4** | **Exp = 5** | **Exp = 9** |
| Yes |  | 0 | 1 | 1 |
| No |  | 4 | 4 | 8 |

Table S30: Frequency of Community Base Health Services

|  | Conservative management | Conservative management plus fludrocortisone | Conservative management plus midodrine | Overall |
| --- | --- | --- | --- | --- |
| **Baseline** | **Exp = 3** | **Exp = 4** | **Exp = 6** | **Exp = 13** |
| Yes | 2 | 0 | 0 | 2 |
| No | 1 | 4 | 6 | 11 |
| **Month 3** | **Exp = 1** | **Exp = 4** | **Exp = 6** | **Exp = 11** |
| Yes | 0 | 1 | 0 | 1 |
| No | 1 | 3 | 6 | 10 |
| **Month 6** | **Exp = 0** | **Exp = 4** | **Exp = 6** | **Exp = 10** |
| Yes |  | 0 | 1 | 1 |
| No |  | 4 | 5 | 9 |
| **Month 12** | **Exp = 0** | **Exp = 4** | **Exp = 5** | **Exp = 9** |
| Yes |  | 0 | 2 | 2 |
| No |  | 4 | 3 | 7 |

Table S31: Completion rates for the EQ-5D-5L (both Self and Proxy completed)

|  | Conservative management | Conservative management plus fludrocortisone | Conservative management plus midodrine | Overall |
| --- | --- | --- | --- | --- |
| **Baseline** | **Exp =3** | **Exp = 4** | **Exp =6** | **Exp = 13** |
| Self-Completed | 3 | 4 | 5 | 12 |
| Proxy Completed | 0 | 0 | 1 | 1 |
| **Month 3** | **Exp = 1** | **Exp = 4** | **Exp = 6** | **Exp = 11** |
| Self-Completed | 1 | 4 | 6 | 11 |
| Proxy Completed | 0 | 0 | 0 | 0 |
| **Month 6** | **Exp = 0** | **Exp = 4** | **Exp = 6** | **Exp = 10** |
| Self-Completed |  | 4 | 4 | 8 |
| Proxy Completed |  | 0 | 2 | 2 |
| **Month 12** | **Exp = 0** | **Exp = 4** | **Exp = 5** | **Exp = 9** |
| Self-Completed |  | 3 | 5 | 8 |
| Proxy Completed |  | 1 | 0 | 1 |

Table S32: Method of EQ-5D-5L data collection by study group

|  | Conservative management | Conservative management plus fludrocortisone | Conservative management plus midodrine | Overall |
| --- | --- | --- | --- | --- |
| **Baseline** | **Exp =3** | **Exp = 4** | **Exp = 6** | **Exp = 13** |
| Face-to-face | 3 | 4 | 6 | 13 |
| Remote | 0 | 0 | 0 | 0 |
| **Month 3** | **Exp = 1** | **Exp = 4** | **Exp = 6** | **Exp = 11** |
| Face-to-face | 1 | 4 | 6 | 11 |
| Remote | 0 | 0 | 0 | 0 |
| **Month 6** | **Exp = 0** | **Exp = 4** | **Exp = 6** | **Exp = 10** |
| Face-to-face |  | 4 | 5 | 9 |
| Remote |  | 0 | 1 | 1 |
| **Month 12** | **Exp = 0** | **Exp = 4** | **Exp = 5** | **Exp = 9** |
| Face-to-face |  | 3 | 5 | 8 |
| Remote |  | 1 | 0 | 1 |

Table S33: EQ-5D-5L Mobility Score

|  | Conservative management | Conservative management plus fludrocortisone | Conservative management plus midodrine | Overall |
| --- | --- | --- | --- | --- |
| **Baseline** | **Exp = 3** | **Exp = 4** | **Exp = 5** | **Exp = 12** |
| 1. No problems in walking about | 1 | 2 | 1 | 4 |
| 2. Slight problems in walking about | 1 | 1 | 3 | 5 |
| 3. Moderate problems in walking about | 1 | 1 | 0 | 2 |
| 4. Severe problems in walking about | 0 | 0 | 1 | 1 |
| 5. Unable to walk about |  |  |  |  |
| Mean (SD) | 2.00 (1) | 1.75 (0.96) | 2.20 (1.10) | 1.75 (0.96) |
| Median (Range) | 0.5 (0-2) | 1.5 (1-3) | 2.00 (1.-4) | 1.50 (1-4) |
| **Month 3** | **Exp = 1** | **Exp = 4** | **Exp = 5** | **Exp = 10** |
| 1. No problems in walking about | 0 | 2 | 0 | 2 |
| 2. Slight problems in walking about | 1 | 1 | 1 | 3 |
| 3. Moderate problems in walking about | 0 | 1 | 2 | 3 |
| 4. Severe problems in walking about | 0 | 0 | 3 | 3 |
| 5. Unable to walk about |  |  |  |  |
| Mean (SD) | 2 (0) | 1,75 (0.96) | 3.33 (0.82) | 2.64 (1.12) |
| Median (Range) | 2 (0) | 1,5 (1-3) | 3.5 (2-4( | 3 (1-4) |
| **Month 6** | **Exp = 0** | **Exp = 4** | **Exp = 4** | **Exp = 8** |
| 1. No problems in walking about |  | 4 | 1 | 5 |
| 2. Slight problems in walking about |  | 0 | 2 | 2 |
| 3. Moderate problems in walking about |  |  |  |  |
| 4. Severe problems in walking about |  | 0 | 1 | 1 |
| 5. Unable to walk about |  |  |  |  |
| Mean (SD) |  | 1 (0) | 2,25 (126) | 1.63 (1.06) |
| Median (Range) |  | 1 (0) | 2 (1-4) | 1 (1-4) |
| **Month 12** | **Exp = 0** | **Exp = 3** | **Exp = 5** | **Exp = 8** |
| 1. No problems in walking about |  | 3 | 1 | 4 |
| 2. Slight problems in walking about |  |  | 1 | 1 |
| 3. Moderate problems in walking about |  |  | 1 | 1 |
| 4. Severe problems in walking about |  |  | 2 | 2 |
| 5. Unable to walk about |  |  |  |  |
| Mean (SD) |  | 1 (0) | 2.8 (1.3) | 2.13 (1.41) |
| Median (Range) |  | 1 (N/A) | 3 (1-4) | 1.5 (1-5) |

Table S34: EQ-5D-5L Self Care

|  | Conservative management | Conservative management plus fludrocortisone | Conservative management plus midodrine | Overall |
| --- | --- | --- | --- | --- |
| **Baseline** | **Exp = 3** | **Exp =** | **Exp =** | **Exp =** |
| 1. No problems with washing and dressing | 1 | 4 | 2 | 7 |
| 2. Slight problems in washing and dressing | 1 | 0 | 2 | 3 |
| 3. Moderate problems in washing and dressing | 1 | 0 | 0 | 1 |
| 4. Severe problems in washing and dressing | 0 | 0 | 1 | 1 |
| 5. Extreme problems in washing and dressing |  |  |  |  |
| Mean (SD) | 2 (1) | 1(0) | 2 (1-1.22) | 1.67 (0.98) |
| Median (Range) | 2 (1-3) | 1 (1) | 2(1-4) | 1 (1-4) |
| **Month 3** | **Exp = 1** | **Exp = 4** | **Exp = 5** | **Exp = 10** |
| 1. No problems with washing and dressing | 0 | 4 | 2 | 6 |
| 2. Slight problems in washing and dressing | 0 | 0 | 2 | 2 |
| 3. Moderate problems in washing and dressing | 1 | 0 | 2 | 3 |
| 4. Severe problems in washing and dressing |  |  |  |  |
| 5. Extreme problems in washing and dressing |  |  |  |  |
| Mean (SD) | 3 (0) | 1 (0) | 2 (0.89) | 1.73 (0.9) |
| Median (Range) | 3 (0) | 1 (0) | 2 (1-3) | 1 (1-3) |
| **Month 6** | **Exp = 0** | **Exp = 4** | **Exp = 4** | **Exp = 8** |
| 1. No problems with washing and dressing |  | 4 | 1 | 5 |
| 2. Slight problems in washing and dressing |  | 0 | 2 | 2 |
| 3. Moderate problems in washing and dressing |  | 0 | 1 | 1 |
| 4. Severe problems in washing and dressing |  |  |  |  |
| 5. Extreme problems in washing and dressing |  |  |  |  |
| Mean (SD) |  | 1 (0) | 2 (0.82) | 1.5 (0.76) |
| Median (Range) |  | 1 (0) | 2 (1-3) | 1 (1-3) |
| **Month 12** | **Exp = 0** | **Exp = 3** | **Exp = 5** | **Exp = 8** |
| 1. No problems with washing and dressing |  | 3 | 1 | 4 |
| 2. Slight problems in washing and dressing |  | 0 | 2 | 2 |
| 3. Moderate problems in washing and dressing |  | 0 | 1 | 1 |
| 4. Severe problems in washing and dressing |  |  |  |  |
| 5. Extreme problems in washing and dressing |  | 0 | 1 | 1 |
| Mean (SD) |  | 1 (N/A) | 2.60 (1.52) | 2.13 (1.36) |
| Median (Range) |  | 1(N/A) | 2 (1-5) | 2 (1-5) |

Table S35: EQ-5D-5L Usual Activity (e.g. work, study, housework, family or leisure activities)

|  | Conservative management | Conservative management plus fludrocortisone | Conservative management plus midodrine | Overall |
| --- | --- | --- | --- | --- |
| **Baseline** | **Exp = 3** | **Exp = 4** | **Exp = 6** | **Exp = 13** |
| 1. No problems doing usual activities | 1 | 3 | 3 | 7 |
| 2. Slight problems doing usual activities | 0 | 0 | 1 | 1 |
| 3. Moderate problems doing usual activities | 0 | 1 | 0 | 1 |
| 4. Severe problems doing usual activities | 1 | 0 | 0 | 1 |
| 5. Unable to do usual activities | 1 | 0 | 1 | 2 |
| Mean (SD) |  | 1.5 (1) | 2 (1.73) | 2.17 (1.64) |
| Median (Range) |  | 1 (1-3) | 1 (1-5) | 1 (1-5) |
| **Month 3** | **Exp = 1** | **Exp = 4** | **Exp = 5** | **Exp = 10** |
| 1. No problems doing usual activities | 0 | 3 | 1 | 4 |
| 2. Slight problems doing usual activities | 1 | 1 | 1 | 3 |
| 3. Moderate problems doing usual activities | 0 | 0 | 1 | 1 |
| 4. Severe problems doing usual activities | 0 | 0 | 2 | 2 |
| 5. Unable to do usual activities | 0 | 0 | 1 | 1 |
| Mean (SD) | 3 (0) | 1.25 (0.5) | 3.17 (1.47) | 2.36 (1.43) |
| Median (Range) | 3 (0) | 1 (1-2) | 3.5 (1-5) | 2 (1-5) |
| **Month 6** | **Exp = 0** | **Exp = 4** | **Exp = 4** | **Exp = 8** |
| 1. No problems doing usual activities |  | 4 | 2 | 6 |
| 2. Slight problems doing usual activities |  | 0 | 1 | 1 |
| 3. Moderate problems doing usual activities |  | 0 | 1 | 1 |
| 4. Severe problems doing usual activities |  |  |  |  |
| 5. Unable to do usual activities |  |  |  |  |
| Mean (SD) |  | 1 (0) | 1.75 (0.96) | 1.38 (0.74) |
| Median (Range) |  | 1 (0) | 1.5 (1-3) | 1 (1-3 ) |
| **Month 12** | **Exp = 0** | **Exp = 3** | **Exp = 5** | **Exp = 8** |
| 1. No problems doing usual activities |  | 2 | 1 | 3 |
| 2. Slight problems doing usual activities |  | 1 | 2 | 3 |
| 3. Moderate problems doing usual activities |  | 0 | 1 | 1 |
| 4. Severe problems doing usual activities |  |  |  |  |
| 5. Unable to do usual activities |  | 0 | 1 | 1 |
| Mean (SD) |  | 1.3 (0.58) | 2.6 (1.52) | 2.13 (1.36) |
| Median (Range) |  | 1(1-2) | 2 (1-5) | 2 (1-5) |

Table S36: EQ-5D-5L Pain/ Discomfort

|  | Conservative management | Conservative management plus fludrocortisone | Conservative management plus midodrine | Overall |
| --- | --- | --- | --- | --- |
| **Baseline** | **Exp = 3** | **Exp = 4** | **Exp = 6** | **Exp = 13** |
| 1. No pain or discomfort | 0 | 1 | 2 | 3 |
| 2. Slight pain or discomfort | 1 | 1 | 0 | 2 |
| 3. Moderate pain or discomfort | 2 | 2 | 2 | 6 |
| 4. Severe pain or discomfort |  |  |  |  |
| 5. Extreme pain or discomfort | 0 | 0 | 1 | 1 |
| Mean (SD) | 2.67 (0.58) | 2.25 (0.96) | 2.6 (1.67) | 2.5 (1.17) |
| Median (Range) | 3 (2-3) | 2.5 ( 1-3) | 3 (1-5) | 3 (1-5) |
| **Month 3** | **Exp = 1** | **Exp = 4** | **Exp = 5** | **Exp = 10** |
| 1. No pain or discomfort | 1 | 3 | 2 | 5 |
| 2. Slight pain or discomfort | 0 | 1 | 1 | 2 |
| 3. Moderate pain or discomfort | 0 | 0 | 3 | 3 |
| 4. Severe pain or discomfort |  |  |  |  |
| 5. Extreme pain or discomfort |  |  |  |  |
| Mean (SD) | 1 (0) | 1.25 (0.5) | 2.17 (0.98) | 2.36 (1.21) |
| Median (Range) | 1 (0) | 1(1-2) | 2.5 (1-3) | 1 (1-3) |
| **Month 6** | **Exp = 0** | **Exp = 4** | **Exp = 4** | **Exp = 8** |
| 1. No pain or discomfort |  | 2 | 1 | 3 |
| 2. Slight pain or discomfort |  | 0 | 2 | 2 |
| 3. Moderate pain or discomfort |  | 1 | 1 | 2 |
| 4. Severe pain or discomfort |  |  |  |  |
| 5. Extreme pain or discomfort |  | 1 | 0 | 1 |
| Mean (SD) |  | 2.5 (1.91) | 2 (0.82) | 2.24 |
| Median (Range) |  | 2 (1-5) | 2 (1-3) | 2 (1-5) |
| **Month 12** | **Exp = 0** | **Exp = 3** | **Exp = 8** | **Exp = 8** |
| 1. No pain or discomfort |  | 2 | 3 | 5 |
| 2. Slight pain or discomfort |  | 1 | 1 | 2 |
| 3. Moderate pain or discomfort |  |  | 1 | 1 |
| 4. Severe pain or discomfort |  |  |  |  |
| 5. Extreme pain or discomfort |  |  |  |  |
| Mean (SD) |  | 1.33 (0.58) | 1.60 (0.89) | 1.50 (0.76) |
| Median (Range) |  | 1 (1-2) | 1 (1-3) | 1 (1-3) |

Table S37: EQ-5D-5L Anxiety and Depression

|  | Conservative management | Conservative management plus fludrocortisone | Conservative management plus midodrine | Overall |
| --- | --- | --- | --- | --- |
| **Baseline** | **Exp = 3** | **Exp = 4** | **Exp = 6** | **Exp = 13** |
| 1. Not anxious or depressed | 1 | 0 | 2 | 3 |
| 2. Slightly anxious or depressed | 2 | 2 | 1 | 5 |
| 3. Moderately anxious or depressed | 0 | 1 | 1 | 2 |
| 4. Severely anxious or depressed | 0 | 0 | 1 | 1 |
| 5. Extremely anxious or depressed | 0 | 1 | 0 | 1 |
| Mean (SD) | 1.67 (0.58) | 3 (1.41) | 2 (1.3) | 2.33 (1.33) |
| Median (Range) | 2 (1-2) | 2.5 (2-5) | 2 (1-4) | 2 (1-5) |
| **Month 3** | **Exp = 1** | **Exp = 4** | **Exp = 5** | **Exp = 10** |
| 1. Not anxious or depressed | 0 | 2 | 1 | 3 |
| 2. Slightly anxious or depressed | 1 | 0 | 2 | 3 |
| 3. Moderately anxious or depressed | 0 | 2 | 2 | 4 |
| 4. Severely anxious or depressed |  |  |  |  |
| 5. Extremely anxious or depressed | 0 | 0 | 1 | 1 |
| Mean (SD) | 2 (0) | 2 (1.15) | 2.67 (1.37) | 2.36 (1.21) |
| Median (Range) | 2 (0)2.36 | 2 (1-3) | 2.5 (1-5) |  |
| **Month 6** | **Exp = 0** | **Exp = 4** | **Exp = 4** | **Exp = 8** |
| 1. Not anxious or depressed |  | 1 | 1 | 2 |
| 2. Slightly anxious or depressed |  | 1 | 1 | 2 |
| 3. Moderately anxious or depressed |  | 1 | 1 | 2 |
| 4. Severely anxious or depressed |  | 1 | 0 | 1 |
| 5. Extremely anxious or depressed |  | 0 | 1 | 1 |
| Mean (SD) |  | 2.5 (1.29) | 2.75 (1.71) | 2.63 (1.41) |
| Median (Range) |  | 2.5 (1-4 ) | 2.5 (1.5) | 2.5 (1-5) |
| **Month 12** | **Exp = 0** | **Exp = 3** | **Exp = 5** | **Exp = 9** |
| 1. Not anxious or depressed |  | 1 | 2 | 3 |
| 2. Slightly anxious or depressed |  |  |  |  |
| 3. Moderately anxious or depressed |  | 1 | 3 | 4 |
| 4. Severely anxious or depressed |  | 1 | 0 | 1 |
| 5. Extremely anxious or depressed |  |  |  |  |
| Mean (SD) |  | 2.67 (1.53) | 2.2 (1.10) | 2.38 (1.19) |
| Median (Range) |  | 2 (1-4) | 3 (1-3) | 3 (1-4) |

Table S38: Health Score Self EQ-5D VAS

|  | Conservative management | Conservative management plus fludrocortisone | Conservative management plus midodrine | Overall |
| --- | --- | --- | --- | --- |
| **Baseline** | **Exp = 3** | **Exp = 4** | **Exp = 6** | **Exp = 13** |
| 50 | 1 | 1 | 2 | 4 |
| 60 | 1 | 2 | 0 | 3 |
| 70 | 0 | 0 | 3 | 3 |
| 80 | 1 | 0 | 0 | 1 |
| 85 | **0** | 1 | 0 | 1 |
| Mean (SD) | 63.33 (15.28) | (63.75 (14.93) | 63.33 (15.28) | 62.92 (12.15) |
| Median (Range) | 60 (50 -80) | 60 (50-85 | 60 (50-80) | 60 (50-85) |
| **Month 3** | **Exp = 1** | **Exp = 4** | **Exp = 5** | **Exp = 10** |
| 40 | 0 | 0 | 2 | 2to |
| 50 | 0 | 1 | 1 | 2 |
| 65 | 0 | 1 | 1 | 2 |
| 70 | 0 | 0 | 1 | 1 |
| 80 | 0 | 1 | 0 | 1 |
| 90 | 1 | 1 | 1 | 3 |
| Mean (SD) | 90 (0) | 71.25 (17.5) | 59.17 (19.60) | 66.36 (19.1) |
| Median (Range) | 90 (0) | 72.50 (50-90) | 57.50 (40 -90 ) | 65 (40-90) |
| **Month 6** | **Exp = 0** | **Exp = 4** | **Exp = 6** | **Exp = 10** |
| 65 |  | 1 | 0 | 1 |
| 70 |  | 3 | 1 | 4 |
| 75 |  | 0 | 1 | 1 |
| 80 |  | 0 | 2 | 2 |
| Mean (SD) |  | 68.75 (2.5) | 76.25 (4.79) | 72.50 (5.35) |
| Median (Range) |  | 70 (65-70) | 77.50 (70-80) | 70 (65-80) |
| **Month 12** | **Exp = 0** | **Exp = 3** | **Exp = 5** | **Exp = 8** |
| 10 |  | 0 | 1 | 1 |
| 50 |  | 0 | 2 | 2 |
| 60 |  | 1 | 1 | 2 |
| 75 |  | 2 | 0 | 2 |
| 85 |  | 0 | 1 | 1 |
| Mean (SD) |  | 70 (8.66) | 50 (25.25) | 57.5 (22.5) |
| Median (Range) |  | 75 (60-75) | 50 (10-80) | 60 (10-80) |

Table S39: EQ-5D-5L Utility

|  | Conservative management | Conservative management plus fludrocortisone | Conservative management plus midodrine | Overall |
| --- | --- | --- | --- | --- |
| **Baseline** | **Exp = 3** | **Exp = 4** | **Exp = 6** | **Exp = 13** |
| Mean (SD) |  | 0.6 (0.24) | 0.53 (0.42) | 0.58 (0.3) |
| Median (Range) |  | 0.67 (0.26 -0.8) | 0.59 (-0.15-0 92) | 0.62 (-0.15-0.92) |
| **Month 3** | **Exp = 1** | **Exp = 4** | **Exp = 5** | **Exp = 10** |
| Mean (SD) | 0.68 (N/A) | 0.84 (0.15) | 0.49 (0.24) | 0.63 (0.25) |
| Median (Range) | 0.68 (N/Q | 0.84 (0.69-0.99) | 0.54 (0.03 -0.71) | 0.68 (0.03-0.99) |
| **Month 6** | **Exp = 0** | **Exp = 4** | **Exp = 4** | **Exp = 8** |
| Mean (SD) |  | 0.76 (0.21) | 0.54 (0.26) | 0.65 (0.25) |
| Median (Range) |  | 0.85 (0.45-0.89) | 0.6 (0.18-0.8) | 0.71 (0.18-0.89) |
| **Month 12** | **Exp = 0** | **Exp = 4** | **Exp = 5** | **Exp = 9** |
| Mean (SD) |  | 0..72 (0.26) | 0.67 (0.78) | 0.62 (0.27) |
| Median (Range) |  | 0.68 (0.47) | 0.70 (0.15) | 0.64 (0.91) |

Table S40: EQ-5D-5L Mobility proxy Score

|  | Conservative management | Conservative management plus fludrocortisone | Conservative management plus midodrine | Overall |
| --- | --- | --- | --- | --- |
| **Baseline** | **Exp = 0** | **Exp = 0** | **Exp = 1** | **Exp = 1** |
| 1. No problems in walking about |  |  |  |  |
| 2. Slight problems in walking about |  |  |  |  |
| 3. Moderate problems in walking about |  |  |  |  |
| 4. Severe problems in walking about |  |  | 1 | 1 |
| 5. Unable to walk about |  |  |  |  |
| Mean (SD) |  |  | 1 (N/A) | 1 (N/A) |
| Median (Range) |  |  | 1 (N/A) | 1 (N/A) |
| **Month 3** | **Exp = 0** | **Exp = 0** | **Exp = 0** | **Exp = 0** |
| 1. No problems in walking about |  |  |  |  |
| 2. Slight problems in walking about |  |  |  |  |
| 3. Moderate problems in walking about |  |  |  |  |
| 4. Severe problems in walking about |  |  |  |  |
| 5. Unable to walk about |  |  |  |  |
| Mean (SD) |  |  |  |  |
| Median (Range) |  |  |  |  |
| **Month 6** | **Exp = 0** | **Exp = 0** | **Exp = 2** | **Exp = 2** |
| 1. No problems in walking about |  |  |  |  |
| 2. Slight problems in walking about |  |  |  |  |
| 3. Moderate problems in walking about |  |  | 1 | 1 |
| 4. Severe problems in walking about |  |  | 1 | 1 |
| 5. Unable to walk about |  |  |  |  |
| Mean (SD) |  |  | 3.5 (0.71) | 3.5 (0.71) |
| Median (Range) |  |  | 3.5 (3-4) | 3.5 (3-4) |
| **Month 12** | **Exp=0** | **Exp = 1** | **Exp = 0** | **Exp = 1** |
| 1. No problems in walking about |  | 1 |  | 1 |
| 2. Slight problems in walking about |  |  |  |  |
| 3. Moderate problems in walking about |  |  |  |  |
| 4. Severe problems in walking about |  |  |  |  |
| 5. Unable to walk about |  |  |  |  |
| Mean (SD) |  | 1 (N/A) |  | 1 (N/A) |
| Median (Range) |  | 1 (N/A) |  | 1 (N/A) |

Table S41: EQ-5D-5L Self Care proxy Score

|  | Conservative management | Conservative management plus fludrocortisone | Conservative management plus midodrine | Overall |
| --- | --- | --- | --- | --- |
| **Baseline** | **Exp = 0** | **Exp = 0** | **Exp = 0** | **Exp = 1** |
| 1. No problems with washing and dressing |  |  |  |  |
| 2. Slight problems in washing and dressing |  |  | **1** | **1** |
| 3. Moderate problems in washing and dressing |  |  |  |  |
| 4. Severe problems in washing and dressing |  |  |  |  |
| 5. Extreme problems in washing and dressing |  |  |  |  |
| Mean (SD) |  |  | 1 (N/A) | 1 (N/A) |
| Median (Range) |  |  | 1 (N/A) | 1 (N/A) |
| **Month 3** | **Exp = 0** | **Exp = 0** | **Exp = 0** | **Exp = 0** |
| 1. No problems with washing and dressing |  |  |  |  |
| 2. Slight problems in washing and dressing |  |  |  |  |
| 3. Moderate problems in washing and dressing |  |  |  |  |
| 4. Severe problems in washing and dressing |  |  |  |  |
| 5. Extreme problems in washing and dressing |  |  |  |  |
| Mean (SD) |  |  |  |  |
| Median (Range) |  |  |  |  |
| **Month 6** | **Exp = 0** | **Exp = 0** | **Exp = 2** | **Exp = 2** |
| 1. No problems with washing and dressing |  |  |  |  |
| 2. Slight problems in washing and dressing |  |  |  |  |
| 3. Moderate problems in washing and dressing |  |  | 1 | 1 |
| 4. Severe problems in washing and dressing |  |  | 1 | 1 |
| 5. Extreme problems in washing and dressing |  |  |  |  |
| Mean (SD) |  |  | 3.5 (0.71) | 3.5 (0.71) |
| Median (Range) |  |  | 3.5 (3-4) | 3.5 (3-4) |
| **Month 12** | **Exp = 0** | **Exp = 1** | **Exp = 0** | **Exp = 1** |
| 1. No problems with washing and dressing |  | 1 |  | 1 |
| 2. Slight problems in washing and dressing |  |  |  |  |
| 3. Moderate problems in washing and dressing |  |  |  |  |
| 4. Severe problems in washing and dressing |  |  |  |  |
| 5. Extreme problems in washing and dressing |  |  |  |  |
| Mean (SD) |  | 1 (N/A) |  | 1 (N/A) |
| Median (Range) |  | 1 (N/A) |  | 1 (N/A) |

Table S42: EQ-5D-5L Usual Activities proxy Score (e.g. work, study, housework, family or leisure activities)

|  | Conservative management | Conservative management plus fludrocortisone | Conservative management plus midodrine | Overall |
| --- | --- | --- | --- | --- |
| **Baseline** | **Exp = 0** | **Exp = 0** | **Exp = 1** | **Exp = 1** |
| 1. No problems doing usual activities |  |  |  |  |
| 2. Slight problems doing usual activities |  |  | 1 | 1 |
| 3. Moderate problems doing usual activities |  |  |  |  |
| 4. Severe problems doing usual activities |  |  |  |  |
| 5. Unable to do usual activities |  |  |  |  |
| Mean (SD) |  |  | 1 (N/A) | 1 (N/A) |
| Median (Range) |  |  | 1 (N/A) | 1 (N/A) |
| **Month 3** | **Exp = 0** | **Exp = 0** | **Exp = 0** | **Exp = 0** |
| 1. No problems doing usual activities |  |  |  |  |
| 2. Slight problems doing usual activities |  |  |  |  |
| 3. Moderate problems doing usual activities |  |  |  |  |
| 4. Severe problems doing usual activities |  |  |  |  |
| 5. Unable to do usual activities |  |  |  |  |
| Mean (SD) |  |  |  |  |
| Median (Range) |  |  |  |  |
| **Month 6** | **Exp = 0** | **Exp = 0** | **Exp = 2** | **Exp = 2** |
| 1. No problems doing usual activities |  |  |  |  |
| 2. Slight problems doing usual activities |  |  |  |  |
| 3. Moderate problems doing usual activities |  |  | 1 | 1 |
| 4. Severe problems doing usual activities |  |  |  |  |
| 5. Unable to do usual activities |  |  | 1 | 1 |
| Mean (SD) |  |  | 4 (1.41) | 4 (1.41) |
| Median (Range) |  |  | 4 (3-5) | 4 (3-5) |
| **Month 12** | **Exp = 0** | **Exp = 1** | **Exp = 0** | **Exp = 1** |
| 1. No problems doing usual activities | 1 |  |  | 1 |
| 2. Slight problems doing usual activities |  |  |  |  |
| 3. Moderate problems doing usual activities |  |  |  |  |
| 4. Severe problems doing usual activities |  |  |  |  |
| 5. Unable to do usual activities |  |  |  |  |
| Mean (SD) |  | 1 (N/A) |  | 1 (N/A) |
| Median (Range) |  | 1 (N/A) |  | 1 (N/A) |

Table S43: Pain and Discomfort Proxy Score

|  | Conservative management | Conservative management plus fludrocortisone | Conservative management plus midodrine | Overall |
| --- | --- | --- | --- | --- |
| **Baseline** | **Exp = 0** | **Exp = 0** | **Exp = 1** | **Exp = 1** |
| 1. No pain or discomfort |  |  |  |  |
| 2. Slight pain or discomfort |  |  |  |  |
| 3. 4. Moderate pain or discomfort |  |  | 1 | 1 |
| 5. Severe pain or discomfort |  |  |  |  |
| 6. Extreme pain or discomfort |  |  |  |  |
| Mean (SD) |  |  | 1 (N/A) | 1 (N/A) |
| Median (Range) |  |  | 1 (N/A) | 1 (N/A) |
| **Month 3** | **Exp = 0** | **Exp = 0** | **Exp = 0** | **Exp = 0** |
| 1. No pain or discomfort |  |  |  |  |
| 2. Slight pain or discomfort |  |  |  |  |
| 3. Moderate pain or discomfort |  |  |  |  |
| 4. Severe pain or discomfort |  |  |  |  |
| 5. Extreme pain or discomfort |  |  |  |  |
| Mean (SD) |  |  |  |  |
| Median (Range) |  |  |  |  |
| **Month 6** | **Exp = 0** | **Exp = 0** | **Exp = 2** | **Exp = 2** |
| 1. No pain or discomfort |  |  |  |  |
| 2. Slight pain or discomfort |  |  |  |  |
| 3. Moderate pain or discomfort |  |  | 1 | 1 |
| 4. Severe pain or discomfort |  |  |  |  |
| 5. Extreme pain or discomfort |  |  | 1 | 1 |
| Mean (SD) |  |  | 4 (1.41) | 4 (1.41) |
| Median (Range) |  |  | 4 (3-5) | 4 (3-5) |
| **Month 12** |  | **Exp = 1** | **Exp = 0** | **Exp = 1** |
| 1. No pain or discomfort |  |  |  |  |
| 2. Slight pain or discomfort |  |  |  |  |
| 3. Moderate pain or discomfort |  | 1 |  | 1 |
| 4. Severe pain or discomfort |  |  |  |  |
| 5. Extreme pain or discomfort |  |  |  |  |
| Mean (SD) |  | 3 (N/A) |  | 3 (N/A) |
| Median (Range) |  | 3 (N/A) |  | 3 (N/A) |

Table S44: EQ-5D-5L Anxiety and Depression Proxy Score

|  | Conservative management | Conservative management plus fludrocortisone | Conservative management plus midodrine | Overall |
| --- | --- | --- | --- | --- |
| **Baseline** | **Exp = 0** | **Exp = 0** | **Exp = 1** | **Exp = 1** |
| 1. Not anxious or depressed |  |  |  |  |
| 2. Slightly anxious or depressed |  |  |  |  |
| 3. Moderately anxious or depressed |  |  | 1 | 1 |
| 4. Severely anxious or depressed |  |  |  |  |
| 5. Extremely anxious or depressed |  |  |  |  |
| Mean (SD) |  |  | 1 (N/A) | 1 (N/A) |
| Median (Range) |  |  | 1 (N/A | 1 (N/A) |
| **Month 3** | **Exp = 0** | **Exp = 0** | **Exp = 0** | **Exp = 0** |
| 1. Not anxious or depressed |  |  |  |  |
| 2. Slightly anxious or depressed |  |  |  |  |
| 3. Moderately anxious or depressed |  |  |  |  |
| 4. Severely anxious or depressed |  |  |  |  |
| 5. Extremely anxious or depressed |  |  |  |  |
| Mean (SD) |  |  |  |  |
| Median (Range) |  |  |  |  |
| **Month 6** | **Exp = 0** | **Exp = 0** | **Exp = 2** | **Exp = 2** |
| 1. Not anxious or depressed |  |  |  |  |
| 2. Slightly anxious or depressed |  |  |  |  |
| 3. Moderately anxious or depressed |  |  | 1 | 1 |
| 4. Severely anxious or depressed |  |  | 1 | 1 |
| 5. Extremely anxious or depressed |  |  |  |  |
| Mean (SD) |  |  | 3.5 (0.71) | 3.5 (0.71) |
| Median (Range) |  |  | 3.5 (3-4) | 3.5 (3-4) |
| **Month 12** | **Exp=0** | **Exp = 1** | **Exp = 0** | **Exp = 1** |
| 1. Not anxious or depressed |  | 1 |  | 1 |
| 2. Slightly anxious or depressed |  |  |  |  |
| 3. Moderately anxious or depressed |  |  |  |  |
| 4. Severely anxious or depressed |  |  |  |  |
| 5. Extremely anxious or depressed |  |  |  |  |
| Mean (SD) |  | 1 (N/A) |  | 1 (N/A) |
| Median (Range) |  | 1 (N/A) |  | 1 (N/A) |

Table S45: EQ-5D-5L Proxy Utility Score

|  | Conservative management | Conservative management plus fludrocortisone | Conservative management plus midodrine | Overall |
| --- | --- | --- | --- | --- |
| **Baseline** | **Exp =0** | **Exp = 0** | **Exp = 1** | **Exp = 1** |
| Mean (SD) |  |  | 0.49 (N/A) | 0.49 (N/A) |
| Median (Range) |  |  | 0.49 (N/A) | 0.49 (N/A) |
| **Month 3** | **Exp = 0** | **Exp = 0** | **Exp = 0** | **Exp = 0** |
| Mean (SD) |  |  |  |  |
| Median (Range) |  |  |  |  |
| **Month 6** | **Exp = 0** | **Exp = 0** | **Exp = 2** | **Exp = 2** |
| Mean (SD) |  |  | 0.1 (0.45) | 0.1 (0.45) |
| Median (Range) |  |  | 0.1 (0.2-0.42) | 0.1 (0.2-0.42) |
| **Month 12** | **Exp = 0** | **Exp =1** | **Exp = 0** | **Exp = 1** |
| Mean (SD) |  | 0.71 (N/A) |  | 0.71 (N/A) |
| Median (Range) |  | 0.71 (N/A) |  | 0.71 (N/A) |

Table S46: EQ-VAS Proxy Score

|  | Conservative management | Conservative management plus fludrocortisone | Conservative management plus midodrine | Overall |
| --- | --- | --- | --- | --- |
| **Baseline** | **Exp = 0** | **Exp = 0** | **Exp = 1** | **Exp = 1** |
| 45 |  |  | **1** | **1** |
| Mean (SD) |  |  | 45 (N/A) | 45 (N/A) |
| Median (Range) |  |  | 45 (N/A) | 45 (N/A) |
| **Month 3** | **Exp = 0** | **Exp = 0** | **Exp = 0** | **Exp = 0** |
| Mean (SD) |  |  |  |  |
| Median (Range) |  |  |  |  |
| **Month 6** | **Exp = 0** | **Exp = 0** | **Exp = 2** | **Exp = 2** |
| 30 |  |  | 1 | 1 |
| 50 |  |  | 1 | 1 |
| Mean (SD) |  |  | 40 (14.14) | 40 (14.14) |
| Median (Range) |  |  | 40 (30-50) | 40 (30-50) |
| **Month 12** | **Exp = 1** | **Exp = 1** | **Exp = 2** | **Exp = 1** |
| 90 |  | 1 |  | 1 |
| Mean (SD) |  | 90 (N/A) |  | 90 (NA) |
| Median (Range) |  | 90 (N/A) |  | 90 (NA) |

Document 1. Statistical Analysis Plan

**Revision history**

| **Version** | **Date** | **Changes made** | **Justification for change** | **Timing of change** |
| --- | --- | --- | --- | --- |
| 1.0 | 15/06/2023 | First version | Not Applicable | Not Applicable |

**Abbreviations**

| ADL | Activities of daily living |
| --- | --- |
| AE | Adverse Event |
| BP | Blood pressure |
| CDMS | Clinical Data Management System |
| CI | Confidence interval |
| DMC | Data Monitoring Committee |
| FWER | Family wise error rate |
| HR | Hazard Ratio |
| IMP | Investigational medicinal product |
| IQR | Interquartile range |
| IR | Incidence Rate |
| IRR | Incidence Rate Ratio |
| ITT | Intention to treat |
| LEDD | Levodopa equivalent daily dose |
| MAR | Missing at random |
| MCID | Minimally clinically important difference |
| MD | Mean difference |
| MedDRA | Medical dictionary for regulatory activities |
| MI | Multiple imputation |
| MNAR | Missing not at random |
| NEADL | Nottingham Extended Activities of Daily Living |
| OH | Orthostatic Hypotension |
| OHDAS | Orthostatic Hypotension Daily Activity Scale |
| OHQ | Orthostatic Hypotension Questionnaire |
| OHSAS | Orthostatic Hypotension Symptom Assessment Scale |
| OT | On-treatment |
| PPI | Patient Public Involvement |
| REML | Restricted maximum likelihood |
| SAE | Serious Adverse Event |
| SAP | Statistical analysis plan |
| SAS | Safety Analysis Set |
| SD | Standard deviation |
| TSC | Trial Steering Committee |
| UPDRS | Unified Parkinson’s Disease Rating Scale |

### 1. **Introduction**

#### 2. Background and rationale

Orthostatic hypotension (OH) is a common and disabling condition characterised by a significant reduction in blood pressure (BP) on standing upright, typically causing dizziness and falls.

There is little good quality evidence to support the management of OH. First line treatment is usually lifestyle advice and non-drug therapies. Where these are not effective there are two pharmacological options, fludrocortisone and midodrine; however, there is a lack of robust evidence on the clinical and cost-effectiveness of these treatment strategies and long-term efficacy and safety is unclear.

The CONFORM-OH trial, commissioned by NIHR HTA, was designed to evaluate the clinical and cost-effectiveness of fludrocortisone and midodrine for the management of symptomatic OH in comparison with conservative management (lifestyle advice and non-drug therapies).

The trial included a 10-month internal pilot phase to assess recruitment and retention with defined progression criteria. At a meeting of the Trial Steering Committee (TSC) on 30^th^ August 2022, nine months into the internal pilot phase, it was agreed the that the trial should close to recruitment on the basis of very low recruitment rates.

This statistical analysis plan describes the trial design, outcome measures and plans to descriptively summarise available data. Details of analyses which would have been performed, had the trial continued to full enrolment, are described in an Appendix.

#### 3. Objectives

This trial was designed to evaluate the clinical and cost-effectiveness of three different treatment strategies for the management of symptomatic OH:

I. Control: Conservative management (lifestyle advice and non-drug therapies)
II. Conservative management plus fludrocortisone
III. Conservative management plus midodrine
     3.1.1 **Primary objective**

To determine whether the treatment strategies of conservative management plus fludrocortisone, and conservative management plus midodrine, improve symptoms of OH compared to conservative management alone, as measured by change in the Orthostatic Hypotension Questionnaire (OHQ) score at six months.

3.1.2 **Secondary objectives**

To determine how the treatment strategies of conservative management plus fludrocortisone, and conservative management plus midodrine, affect the following outcomes compared to conservative management alone over a 12-month period:

1. Activities of daily living (ADLs) measured by the Nottingham Extended ADL (NEADL) scale
2. Falls and syncope (number of falls, number of fallers/non-fallers, fall rate per person year, time to first fall, fall-related injuries, number of syncopal events)
3. Standing blood pressure and postural blood pressure drop
4. Side effects and the safety data associated with each treatment strategy
5. *Health-related quality of life measured by the EQ-5D-5L*
6. *Quality adjusted life years (QALYs) estimated from responses to the EQ-5D-5L and data derived from the literature*
7. *Costs to the NHS, personal social services and patients*
8. *Cost-effectiveness of each treatment strategy modelled from a patient and NHS and personal social services perspective measured in terms of the incremental costs per QALY gained*

*Objectives 5-8 are outside the scope of this analysis plan and will be covered in the Health Economics Analysis Plan.*

3.1.3 **Exploratory objectives**

An exploratory objective was to compare the clinical effectiveness (measured by change in the OHQ score at six months) of fludrocortisone with midodrine.

### 4. **study methods**

#### 5. Trial design

CONFORM-OH is a pragmatic, multi-arm, multi-stage, parallel group, prospective, randomised, open label, superiority trial. The trial was designed to assess the effectiveness of two drug therapies to improve the symptoms of OH compared to conservative management.

Adult patients presenting with symptomatic OH refractory to lifestyle modification were allocated (1:1:1) to receive either conservative management alone, conservative management plus fludrocortisone or conservative management plus midodrine for a period of 12 months.

The primary outcome is assessed after a period of 6 months. This timing was chosen based on a number of factors, including patient consultation, allowing adequate time for dose titration, and balancing adequate exposure with likely adherence and retention rates. Longer term follow-up will continue to 12 months.

An interim analysis, based on available three- and six-month primary outcome data, was planned to take place after the 200^th^ patient had been recruited. If an intervention arm showed no benefit compared to the control arm (see Appendix for further details) it would be recommended to be dropped from the study, with control (conservative management) and the alternative intervention arm continuing to the planned recruitment target of 366 participants in a 1:1 ratio. If both intervention arms showed lack of benefit, the study would be recommended to stop. Recruitment would continue to all three arms while the interim analysis was being conducted.

The trial also included a 10-month internal pilot phase to assess recruitment and retention rates.

#### 6. Study setting and patient population

Adult patients with symptomatic OH refractory to a minimum of 4 weeks of lifestyle modification were recruited. Patients were to be recruited from approximately 20 NHS trusts across the UK, typically from secondary care settings such as falls clinics, day hospitals, geriatric medicine clinics and movement disorder clinics.

For a full list of inclusion and exclusion criteria refer to section 4 of the study protocol.

#### 7. Randomisation and blinding

Participants were allocated in a 1:1:1 ratio to receive conservative management alone, conservative management plus fludrocortisone or conservative management plus midodrine. A minimisation algorithm with a random element was used to assign treatment allocation. Minimisation factors were age (≥80 vs <80 years), aetiology (neurogenic vs non-neurogenic OH) and recruiting site/centre (to account for possible difference in usual care practice).

The minimisation system was provided by SealedEnvelope^TM^ as a 24-hour, central, secure, web-based system accessed by delegated members of the research team at each site to perform randomisation.

The minimisation algorithm incorporates a random element such that there is an 80% chance the participant is allocated to the arm which minimises imbalance, with the remaining arms chosen with 10% probability each. In the case of ties, one of the tied arms is chosen at random to be the ‘preferred’ arm and assigned an 80% probability of being chosen with the remaining arms assigned a 10% probability each.

This is an open-label trial and there will be no blinding of participants, clinicians or research staff. It was planned that the trial statistician would not have access to outcome data by treatment group until the end of the trial. The interim analysis and closed reports to the Data Monitoring Committee (DMC), i.e. containing data presented by randomised treatment group, would have been performed/prepared by a statistician (a member of the Biostatistics Research Group) not otherwise involved in the study and reviewed by the Lead Statistician. However, following the decision to close the trial early it was agreed the trial statistician no longer needed to remain blinded. The perceived risk of having an unblinded statistician in an otherwise open-label trial was deemed to be minimal given that data would only be summarised descriptively and no interim or final inferential analyses would be performed.

#### 8. Definition of outcome measures

#### 8.1.1 Primary endpoint

The Orthostatic Hypotension Questionnaire (OHQ) [1] is completed at baseline, three, six and 12 month follow-up. The primary endpoint is the overall composite OHQ score at six months. The OHQ is validated for use in both clinical and research settings [2] and will be scored according to the validated scoring method [1].

The OHQ is composed of two sections: the first section consists of six questions, which rate the severity of six different symptoms on a scale of 0 to 10 (Orthostatic Hypotension Symptom Assessment Scale – OHSAS). The second section is composed of four questions, which rate the impact of symptoms on standing and walking (Orthostatic Hypotension Daily Activity Scale – OHDAS). The questions on the OHDAS are scored on a scale of 0 to 10 but also include an option of ‘cannot be done for other reasons’.

The OHSAS score is calculated by averaging the responses to the six questions on the OHSAS. Similarly, the OHDAS score is calculated by averaging responses to the four questions on the OHDAS. Items which are scored as zero or answered ‘cannot be done for other reasons’ at baseline are not included in the scoring. Post-baseline scores are calculated using only those items which were included in the baseline score. If any item on the OHDAS which was included in the baseline score is missing or answered ‘cannot do for other reasons’ at a post-baseline assessment a value will be assigned using last observation carried forward.

An overall composite OHQ score will be calculated by averaging the OHSAS score and the OHDAS score. This score takes a value between 0-10, with higher scores indicating worse symptoms / interference.

At each follow-up time point the change from baseline will also be calculated as the follow-up score minus the baseline score.

#### 8.1.2 Secondary endpoints

Activities of daily living (ADLs) measured by the Nottingham Extended ADL scale

The Nottingham Extended ADL (NEADL) scale is a 22 item questionnaire designed to assess the level of independence in carrying out social and domestic activities [3]. There are four sections/subscales which are mobility (six items), kitchen (five items), domestic (five items) and leisure (six items). Each item is scored as “on my own”; “own my own with difficulty”; “with help”; and “not at all/no”.

When no more than 2 items are missing within subscales we will use simple imputation methods by replacing the missing item with the median response from the respondent specific completed questions within the subscale. This is can provide a valid approach for psychometrically validated questionnaires where responses to items within subscales are correlated [4]. A similar approach has also been used in other randomised controlled trials involving the NEADL scale [5].

One point is awarded per item if the participant selects either “on my own” or “on my own with difficulty.” A score of zero is awarded if the participants selects “with help” or “not at all/no”. All items are then summed to give an overall NEADL score which ranges from 0 – 22. Higher scores indicate greater independence. The score will not be calculated if any items are missing, after using the imputation method described above. The NEADL questionnaire is measured at baseline, three, six and 12 month follow-up. At each follow-up time point the change from baseline will also be calculated as the follow-up score minus the baseline score.

Falls and syncope (number of falls, number of fallers/non-fallers, fall rate per person year, time to first fall, fall-related injuries, number of syncopal events)

Falls and faints (syncopal events) are self-reported by the participant and collected in monthly falls diaries which should be returned at three, six and 12 month follow up time points.

The number of falls for each participant will be summed over the 12 month period. The fall rate per person year will be calculated in each arm as the total number of falls from all participants in that arm divided by the total observation time for all participants in that arm. For each participant their observation time will be measured as the time (in years) from randomisation to the date of the last completed fall diary.

Participants will be classed as fallers if they report at least one fall over the 12 month period and a non-faller if they did not report a fall and returned at least one fall diary; i.e. participants who do not return at least one fall diary will be excluded from the analysis. We will also categorise participants as a single faller if they fall once, a recurrent faller if they fall twice or more, and a non-faller if they do not report any falls.

The number of faints (syncopal events) for each participant will be summed over the 12 month period. The syncopal event rate will be calculated as described above. In addition, we will report the total number, and rate of, a combined outcome of fall and faints.

Time to first fall will be measured as the time from randomisation to the first reported fall.

Fall related injuries are reported as free-text fields. There will be a medical review of any coding of free-text fields required.

Standing blood pressure and postural blood pressure drop

The lowest (nadir) standing blood pressure (systolic and diastolic) is measured at baseline, three, six and 12 month follow-up.

Blood pressure will also be measured in the supine position (lying down) at baseline, three, six and 12 month follow-up.

Postural blood pressure drop will be defined as (supine – lowest standing blood pressure) for systolic and diastolic measurements at each time point.

For each measurement the change from baseline will also be calculated as the follow-up value minus the baseline value.

Hospital admissions

Hospital admissions during the 12 month trial period will be collected from medical records. The number of admissions per participant will be calculated.

Reasons for admission will be tabulated. ‘Other’ reasons will be coded/grouped where possible with a medical review of any coding.

The rate of hospital admissions per person year will be calculated in each arm as the total number of admissions from all participants in that arm divided by the total observation time for all participants in that arm. For each participant their observation time will be measured as the time (in years) from randomisation to their last completed follow-up visit.

Side effects and the safety data associated with each treatment strategy

Adverse Events (AEs) will be collected over the 12-month trial period. At each follow-up visit participants will be asked about any side effects or adverse events they have experienced. This will be by open-ended questioning rather than a review of specific side effects or symptoms. Adverse events will be coded using the MedDRA dictionary (version 24) and summarised at the preferred term level. Severity (mild / moderate / severe), seriousness (Yes / No) and relationship to study treatment (Unrelated / Unlikely to be related / Possibly related /Probably related / Definitely related) will also be collected.

An adverse event of the same type (i.e. same preferred term) will be counted as a separate occurrence if the start and end date of sequential events are separated by > 1 day.

Further detail on how adverse events will be summarised and reported is provided in section 6.

#### 8.1.3 Exploratory endpoints

There are no planned exploratory endpoints, however, had the trial continued to full enrolment, an exploratory analysis would have compared the primary endpoint between the two intervention arms (conservative management plus fludrocortisone and conservative management plus midodrine).

#### 9. Study assessments

Participants are assessed at three, six and 12 months from trial entry. As this is a pragmatic trial, assessments can take place within +/- 4, 6 and 8 weeks for the three, six and 12 month visits respectively. A simplified schedule of assessment is given in **Table 1**.

**Table 1: Simplified schedule of assessments**

| Form | Visit | | | |
| --- | --- | --- | --- | --- |
|  | Baseline | Month 3  (+/- 4 weeks) | Month 6  (+/- 6 weeks) | Month 12  (+/- 8 weeks) |
| Demographics | X |  |  |  |
| Medical History | X |  |  |  |
| UPDRS* | X |  |  |  |
| Blood pressure | X | X | X | X |
| Culprit medication review | X | X | X | X |
| OHQ | X | X | X | X |
| NEADL | X | X | X | X |
| EQ-5D-5L | X | X | X | X |
| Falls diary return |  | X | X | X |
| Hospital admission review |  | X | X | X |

**Only for participants with Parkinson’s disease, Dementia with Lewy bodies or Multi System Atrophy*

#### 10. Sample size and power

The trial was designed to test two null hypotheses:

I. The mean difference in six-month OHQ score between conservative management plus fludrocortisone and conservative management alone is = 0
II. The mean difference in six-month OHQ score between conservative management plus midodrine and conservative management alone is = 0

Using standard sample size formula for a two sample t-test, a three-arm trial without an interim analysis would require 103 participants per arm to detect a difference of 1.0 point on the OHQ with 90% power, a two-sided type I error of 5% (equivalently a one-sided type I error of 2.5%), and assuming a standard deviation of 2.2.

A difference of 1.0 point on the OHQ is used as this represents an established minimally clinically important difference (MCID) [1] and was confirmed following discussion with patient and public involvement (PPI) representatives. A standard deviation of 2.2 was assumed based on local audit data from a cohort of 100 neurogenic and non-neurogenic patients.

Assuming attrition of 15% (based on comparable clinical trials in older people of similar duration [6,7] the recruitment target was 366 (122 per arm).

If at the interim analysis an intervention arm showed no benefit compared to the control arm (i.e. if the estimated mean OHQ score at 6 months was worse than control) it would be recommended to be dropped from the study. The multi-arm design would control the one-sided family-wise error rate (FWER), the total chance of falsely recommending an ineffective treatment, at 4.5%. With the possibility of early lack-of-benefit stopping, the FWER would be lower than this. The pair-wise error rate (PWER) would be controlled at 2.5% for each comparison.

Taking into account the interim analysis, which would be conducted once 200 participants had been recruited, the power of the design to recommend each treatment is displayed in the table below (as determined from one million simulation replicates per scenario).

We assumed participants would be recruited at a rate of 0.8 per site per month over a 30-month period with a staggered opening of sites (six sites open for the first six months with two new sites opening per month from month 7 onwards until a total of 20 sites are open). Under this assumed pattern of recruitment we would expect around 150 and 100 participants to have reached the three and six month follow-up time points respectively. The below table assumes only participants with six month data collected are included in the interim analysis. In practice including participants with three-month data but not yet six-month data would allow modest additional efficiency.

| Scenario | Probability recommend fludrocortisone | Probability recommend midodrine | Probability recommend both | Probability recommend at least one |
| --- | --- | --- | --- | --- |
| Mean effect of both = 0 (null scenario) | 2.4% | 2.4% | 0.4% | 4.4% |
| Fludrocortisone has MCID, midodrine effect = 0 | 91.8% | 2.5% | 2.5% | 91.8% |
| Midodrine has MCID, fludrocortisone effect = 0 | 2.4% | 91.7% | 2.4% | 91.7% |
| Both treatments have MCID effect | 89.9% | 89.7% | 82.8% | 96.8% |

*R code used to perform the power calculations can be found in S:\School Statistics\NCTU\CONFORM-OH\2. Study design\2.1 Statistical considerations\Sample size calculation*

### 11. **Statistical considerations**

#### 12. Timing of final analysis

The final analysis will take place once the last 12-month follow-up visit is complete. Once all data queries have been resolved (as far as possible) the database will be locked and the final analysis will commence.

#### 13. Interim analyses, stopping guidelines and data monitoring

Had the trial continued past the internal pilot phase an interim analysis would have been performed based on available three and six month primary outcome data following recruitment of the 200^th^ participant.

Analysis methods which would have been followed for the interim analysis can be found in the Appendix.

The DMC will meet periodically to review progress of the trial and the safety of trial participants. Further detail on the roles and responsibilities of the DMC can be found in the current version of the DMC charter. The DMC will have access to unblind outcome data. With the exception of the interim analysis time point (which will no longer be reached), outcome data will be summarised descriptively and no formal analyses will be conducted.

#### 14. Confidence intervals and p-values

Had the trial continued to full enrolment all statistical tests for the final analysis would have been 2-sided and performed using a 5% significance level. For the interim analysis, an experimental arm would have been recommended to be dropped if the estimated mean between-group difference for an experimental arm is ≥0 (corresponding to the arm having worse outcome than control). This is equivalent to a one-sided p-value of >0.5.

Given that the trial has ended early no statistical testing will be performed. Any confidence intervals will be reported at the 95% level.

#### 15. Analysis sets

The following analysis sets will be defined:

| Analysis Set 1 | - Participants will be analysed according to the treatment group they were randomised to receive, i.e. following the intention-to-treat (ITT) principle - All available outcome data will be included in the analysis |
| --- | --- |
| Analysis Set 2 | - Participants will be analysed according to the treatment group they were randomised to receive, i.e. following the intention-to-treat (ITT) principle - Outcome data following cross-over to an alternative treatment group, or discontinuation of allocated trial treatment, will be set to missing |
| Safety Analysis Set (SAS) | - For each treatment group, the Safety Analysis Set will comprise all participants exposed to that treatment strategy - Safety data (adverse events) will be summarised according to the treatment strategy received at the time of onset of each adverse event. - Participants who cross-over between treatment groups will be included in the Safety Analysis Set for each treatment group |

Primary and secondary clinical outcome measures will be reported according to randomised treatment group; safety data will be reported according to the treatment strategies received. Analysis Set 1 will be the main analysis set used to report the primary and secondary clinical outcome measures. Analysis Set 2 may also be used to provide supplementary information on the primary outcome measure. All safety data will be reported using the Safety Analysis Set.

### 16. **STUDY POPULATION**

#### 17. Participant flow through trial

Participant flow through the trial will be presented using a CONSORT diagram, see Example Figure 1. Information will be provided on numbers and reasons for: screened patients not being eligible; eligible patients not being randomised; participants found to be ineligible after randomisation; participants deviating from allocated treatment; participants not evaluable for the primary endpoints and participant withdrawal from follow-up.

**Example Figure 1: CONSORT flow diagram**

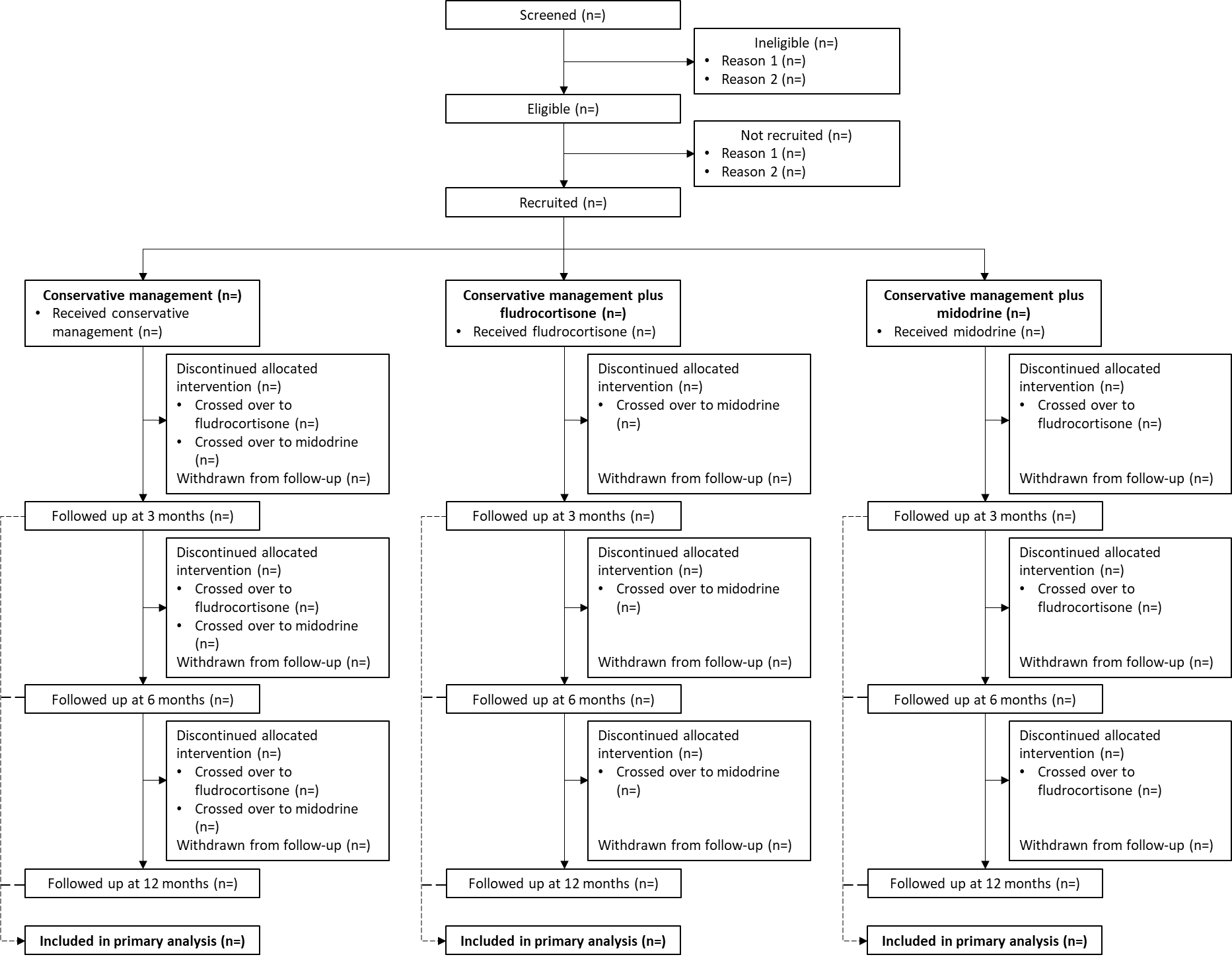

#### 17.1.1 Screening, eligibility and recruitment

The representativeness of the study sample will be assessed using the following data (shown in the CONSORT flow diagram):

- The number of patients identified at screening
- The number of patients screened and not meeting eligibility criteria (with reasons)
- The number of eligible patients identified at screening
- The number of eligible patients not taking part in the study (with reasons where available)
- The number of eligible patients randomised into the study

Data will be reported overall and by site. Observed and projected recruitment rates over time will be plotted.

#### 17.1.2 Protocol deviations

Protocol deviations will be captured on a Deviation Tracking Log which will be held centrally by the Newcastle Clinical Trials Unit (NCTU).

Cross-over between treatment groups may occur during the course of this study and is permitted within the protocol; however, clinical teams will be asked to allow sufficient time, and ideally until the primary outcome is assessed at six months, before considering changing allocated treatment. Supplementary reporting of the primary outcome measure may be conducted using only data collected prior to cross-over or treatment discontinuation (see section 3.4). The number of participants changing treatment allocation or prematurely discontinuing their allocated trial treatment will be reported (see section 4.3).

Given the pragmatic nature of this trial we have not pre-specified any major protocol deviations which would affect the validity of the study or have a direct bearing on the primary outcome analysis or analysis populations.

A listing of all reported protocol deviations will be provided to the trial statistician at the end of the study. The number and type of protocol deviations will be summarised by treatment group.

#### 17.1.3 Follow-up

Availability of outcome data at three, six and 12 month visits will be tabulated as frequency and percentage in each randomised group. The reasons for outcome assessments not being completed will be tabulated where available (e.g. due to withdrawal, death, loss to follow-up, participant too unwell etc.). The timing of outcome assessments (measured as days from randomisation) will be summarised by the median and range (minimum and maximum). The frequency and percentage of outcome assessments performed within protocol specified visit windows will be tabulated. Missing items within the OHQ and NEADL questionnaires will also be summarised by time point and treatment group.

Participants may withdraw consent to provide any further follow-up data. Unless the participant requests otherwise, routinely collected outcome data such as blood pressure measurements will be obtained where available from medical records. The number of participants withdrawing from follow-up (and whether they will allow continued collection of routine data) will be tabulated as frequency and percentage in each randomised group, with reasons where available. The timing of withdrawal (i.e. prior to month three, month six or month 12) will also be summarised, see example Table 1.

The number, and timing of, withdrawals will also be presented in a CONSORT diagram (Example Figure 1), with numbers of participants withdrawing between each stage (trial entry, month three, month six, month 12) shown.

**Example Table 1: Withdrawals from trial by treatment group**

|  | Conservative management | Conservative management plus fludrocortisone | Conservative management plus midodrine | Overall |
| --- | --- | --- | --- | --- |
|  | N= | N= | N= | N= |
| **By 3 months** |  |  |  |  |
| Reason 1 |  |  |  |  |
| Reason 2 |  |  |  |  |
| Reason 3 |  |  |  |  |
| Allowing routine data collection |  |  |  |  |
| **By 6 months** |  |  |  |  |
| Reason 1 |  |  |  |  |
| Reason 2 |  |  |  |  |
| Reason 3 |  |  |  |  |
| Allowing routine data collection |  |  |  |  |
| **By 12 months** |  |  |  |  |
| Reason 1 |  |  |  |  |
| Reason 2 |  |  |  |  |
| Reason 3 |  |  |  |  |
| Allowing routine data collection |  |  |  |  |

Data are n (%)

#### 18. Baseline characteristics

Baseline characteristics will be summarised descriptively, both overall and by randomised treatment group. Categorical variables will be summarised by frequency and percentage. Continuous data will be summarised by the mean and standard deviation and/or median and range, as appropriate. No significance testing will be carried out due to the randomised nature of the study.

Parts I, II and IV of the Unified Parkinson’s Disease Rating Scale (UPDRS) will be completed at baseline for participants with Parkinson’s disease, dementia with Lewy bodies or Multi system atrophy. Part I and Part II each comprise 13 questions rated on a 5-point scale from 0 (normal) to 4 (severe). The Part I and Part II subscale scores are calculated by summing the responses to each of the 13 questions to obtain a score ranging from 0 to 52, with higher scores corresponding to worse outcomes. When no more than 20% of items are missing within subscales we will use simple imputation methods by replacing the missing item with the median response from the respondent specific completed questions within the subscale. If more than 20% of items are missing the subscale score will be set as missing. Part IV comprises six questions, rated on a 5-point scale from 0 (normal) to 4 (severe). The Part IV subscale score is calculated by summing the responses to each of the 6 questions to obtain a score ranging from 0 to 24, with higher scores corresponding to worse outcomes. When no more than 20% of items are missing we will use simple imputation methods by replacing the missing item with the median response from the respondent specific completed questions within Part IV. If more than 20% of items are missing the subscale score will be set as missing.

Details of characteristics to be reported are given in Example Table 2 below.

**Example Table 2: Baseline characteristics**

|  | Conservative management | Conservative management plus fludrocortisone | Conservative management plus midodrine | Overall |
| --- | --- | --- | --- | --- |
|  | N= | N= | N= | N= |
| **Demographics** | | | | |
| Age (years)^1^ |  |  |  |  |
| < 80 years^2^ |  |  |  |  |
| ≥80 years^2^ |  |  |  |  |
| Sex |  |  |  |  |
| Male^2^ |  |  |  |  |
| Female^2^ |  |  |  |  |
| Ethnicity |  |  |  |  |
| Any white background^2^ |  |  |  |  |
| Mixed^2^ |  |  |  |  |
| Asian^2^ |  |  |  |  |
| African^2^ |  |  |  |  |
| Chinese^2^ |  |  |  |  |
| Other^2^ |  |  |  |  |
| **Orthostatic Hypotension Questionnaire (OHQ)** | | | | |
| Overall OHQ score^1^ |  |  |  |  |
| Mean; SD |  |  |  |  |
| Median (IQR); Range |  |  |  |  |
| **Blood pressure (mmHg)** | | | | |
| Supine blood pressure |  |  |  |  |
| Systolic^1^ |  |  |  |  |
| Diastolic^1^ |  |  |  |  |
| Lowest standing blood pressure |  |  |  |  |
| Systolic^1^ |  |  |  |  |
| Diastolic^1^ |  |  |  |  |
| Postural blood pressure drop |  |  |  |  |
| Systolic^1^ |  |  |  |  |
| Diastolic^1^ |  |  |  |  |
| **Medical history** | | | | |
| Disease aetiology |  |  |  |  |
| Neurogenic^2^ |  |  |  |  |
| Non-neurogenic^2^ |  |  |  |  |
| Diabetes^2^ |  |  |  |  |
| Type 1^2^ |  |  |  |  |
| Type 2^2^ |  |  |  |  |
| Pure autonomic failure^2^ |  |  |  |  |
| Parkinson's disease^2^ |  |  |  |  |
| *Disease duration (months)* ^1^ |  |  |  |  |
| Multi system atrophy^2^ |  |  |  |  |
| *Disease duration (months)* ^1^ |  |  |  |  |
| Dementia with Lewy bodies^2^ |  |  |  |  |
| *Disease duration (months)* ^1^ |  |  |  |  |
| **UPDRS^3^** |  |  |  |  |
| Part I |  |  |  |  |
| Part II |  |  |  |  |
| Part IV |  |  |  |  |

Unless otherwise stated data are; ^1^ mean (SD) and/or median (range); ^2^ n (%)

^3^Only completed for participants with Parkinson’s disease, dementia with Lewy bodies or Multi system atrophy

#### 19. Treatment compliance

Participants will be randomised to receive conservative management, conservative management plus fludrocortisone or conservative management plus midodrine.

Conservative management is standard first-line care and forms the control arm of this study. Conservative management consists of non-pharmacologic therapy and will be implemented according to each site’s usual clinical practice. The conservative, non-pharmacological measures advised at baseline will be tabulated by randomised treatment group.

Fludrocortisone and midodrine will be prescribed according to local clinical practice.

Given the pragmatic nature of this study adherence to prescribed medication and non-drug therapies will not be monitored. Participants may cross-over between treatment arms and this will be recorded. Participants who cross-over between treatment groups, or discontinue trial treatment should remain in the trial and be offered all follow-up visits.

The frequency and percentage of participants prematurely discontinuing their allocated trial treatment will be reported along with a reason, where available, and the timing of treatment discontinuation. Whether the participant crossed over to an alternative treatment group will be tabulated, see Example Table 3. Multiple cross-overs between treatment groups will be reported if this occurs.

For each pharmacological intervention (fludrocortisone and midodrine) the number of participants prescribed each treatment (regardless of allocated treatment group) will be reported along with the duration they received the treatment within the 12 month trial period and whether the treatment was prematurely discontinued, with reasons where available. See Example Table 4.

We will also report the frequency and percentage of participants taking ‘culprit medications’ at baseline and each follow-up visit and any changes to these since the previous visit (started, stopped, dose increased, dose decreased, or no change). This will be reported by randomised treatment group for each type of medication collected in the study database. In addition, for participants with Parkinson’s disease, we will calculate the Levodopa equivalent daily dose (LEDD) at baseline and each follow-up visit according to the method proposed by Tomlinson et al [8]. This will be reported in each randomised treatment group as the median and range. See Example Table 5.

**Example Table 3: Treatment adherence**

|  | Conservative management | Conservative management plus fludrocortisone | Conservative management plus midodrine |
| --- | --- | --- | --- |
|  | N= | N= | N= |
| Discontinued allocated treatment |  |  |  |
| Yes |  |  |  |
| No |  |  |  |
| *If yes, due to* |  |  |  |
| Lack of efficacy |  |  |  |
| Side effects |  |  |  |
| Other reason |  |  |  |
| Time from randomisation to discontinuation of allocated treatment |  |  |  |
| <3 months |  |  |  |
| 3-6 months |  |  |  |
| >6 months |  |  |  |
| Crossed over to another treatment group |  |  |  |
| Yes |  |  |  |
| No |  |  |  |
| *If yes, switched to* |  |  |  |
| Conservative management | NA |  |  |
| Conservative management plus fludrocortisone |  | NA |  |
| Conservative management plus midodrine |  |  | NA |

Data are n(%)

**Example Table 4: Trial pharmacological treatments received**

|  | Fludrocortisone | Midodrine |
| --- | --- | --- |
| Prescribed treatment |  |  |
| Duration of treatment (months) |  |  |
| Median; Range |  |  |
| Prematurely discontinued treatment |  |  |
| Yes |  |  |
| No |  |  |
| *If yes, due to* |  |  |
| Lack of efficacy |  |  |
| Side effects |  |  |
| Other reason |  |  |

Data are n(%) unless otherwise stated

**Example Table 5: Culprit medications**

|  | Conservative management | Conservative management plus fludrocortisone | Conservative management plus midodrine |
| --- | --- | --- | --- |
|  | N= | N= | N= |
| ***Culprit medication 1**** | | | |
| Baseline^1^ |  |  |  |
| Month 3^1^ |  |  |  |
| *Started since baseline* |  |  |  |
| *Stopped since baseline* |  |  |  |
| *Dose increased* |  |  |  |
| *Dose decreased* |  |  |  |
| *No change* |  |  |  |
| Month 6^1^ |  |  |  |
| *Started since Month 3* |  |  |  |
| *Stopped since Month 3* |  |  |  |
| *Dose increased* |  |  |  |
| *Dose decreased* |  |  |  |
| *No change* |  |  |  |
| Month 12^1^ |  |  |  |
| *Started since Month 6* |  |  |  |
| *Stopped since Month 6* |  |  |  |
| *Dose increased* |  |  |  |
| *Dose decreased* |  |  |  |
| *No change* |  |  |  |
| **LEDD** | | | |
| Baseline |  |  |  |
| Number with data |  |  |  |
| Median; Range |  |  |  |
| Month 3 |  |  |  |
| Number with data |  |  |  |
| Median; Range |  |  |  |
| Month 6 |  |  |  |
| Number with data |  |  |  |
| Median; Range |  |  |  |
| Month 12 |  |  |  |
| Number with data |  |  |  |
| Median; Range |  |  |  |

**Will be repeated for each culprit medication.*

*^1^Will be reported as Number taking at visit / number assessed at visit (%)*

### 20. **analysIs methods**

#### 21. Primary outcome measure

#### 21.1.1 Main analysis methods

Analysis Set 1 will be used for analysis (see Section 3.4). All data will be reported according to randomised treatment allocation and all available outcome data will be included.

The OHQ score and change from baseline at each follow-up visit will be summarised descriptively by randomised treatment group. The number with data, mean, standard deviation, median and range will be reported, see Example Table 6a. The OHSAS and OHDAS may also be summarised descriptively.

**Example Table 6a: Summary of OHQ scores and change from baseline**

| OHQ score | Value at follow-up visit | | | | Change from baseline | | |
| --- | --- | --- | --- | --- | --- | --- | --- |
|  | Control | Fludrocortisone | Midodrine | Control | | Fludrocortisone | Midodrine |
| Month 3 |  |  |  |  | |  |  |
| N |  |  |  |  | |  |  |
| Mean (SD) |  |  |  |  | |  |  |
| Median (Range) |  |  |  |  | |  |  |
| Month 6 |  |  |  |  | |  |  |
| N |  |  |  |  | |  |  |
| Mean (SD) |  |  |  |  | |  |  |
| Median (Range) |  |  |  |  | |  |  |
| Month 12 |  |  |  |  | |  |  |
| N |  |  |  |  | |  |  |
| Mean (SD) |  |  |  |  | |  |  |
| Median (Range) |  |  |  |  | |  |  |

Subject to sufficient numbers with data (at least three participants per group with outcome data), we will also estimate the mean difference in the change in OHQ score from baseline to each follow-up time point between each intervention and control group, see Example Table 6b. This will be reported with a standard deviation and 95% CI. The purpose of this is to inform potential future meta-analyses. No inferences will be drawn from this data.

**Example Table 6b: Change in OHQ score - mean difference between groups**

| OHQ score | Mean difference (MD) | |
| --- | --- | --- |
|  | Fludrocortisone - control | Midodrine - control |
|  | MD (SD; 95% CI) | MD (SD; 95% CI) |
| Month 3 |  |  |
| Month 6 |  |  |
| Month 12 |  |  |

To inform future sample size calculations will we report the correlation (Pearson’s correlation coefficient) between baseline and follow-up OHQ scores. This will be reported within each randomised treatment group and overall.

**Example Table 6c: Correlation between baseline and follow-up OHQ scores**

| OHQ score | Pearson’s correlation coefficient | | | |
| --- | --- | --- | --- | --- |
|  | Control | Fludrocortisone | Midodrine | Overall |
| Month 3 |  |  |  |  |
| Month 6 |  |  |  |  |
| Month 12 |  |  |  |  |

#### 21.1.2 Supplementary analyses

Primary outcome data may also be reported using Analysis Set 2 (see Section 3.4). Data will be reported according to allocated treatment group but data collected after discontinuation of allocated treatment or initiation of non-allocated fludrocortisone or midodrine will be set as missing. This dataset will be reported descriptively as described in section 5.1.1. Data will only be reported in this way if there is at least three participants per group with outcome data at each follow-up visit.

#### 22. Analysis of secondary outcomes

Activities of daily living (ADLs) measured by the Nottingham Extended ADL scale

Analysis Set 1 will be used for analysis. All available data will be reported according to randomised treatment allocation.

The NEADL score and change from baseline at each follow-up visit will be summarised descriptively by randomised treatment group. The number with data, mean, standard deviation, median and range will be reported, see Example Table 7.

**Example Table 7: NEADL**

| NEADL score | Value at follow-up visit | | | Change from baseline | | |
| --- | --- | --- | --- | --- | --- | --- |
|  | Control | Fludrocortisone | Midodrine | Control | Fludrocortisone | Midodrine |
| Month 3 |  |  |  |  |  |  |
| N |  |  |  |  |  |  |
| Mean (SD) |  |  |  |  |  |  |
| Median (Range) |  |  |  |  |  |  |
| Month 6 |  |  |  |  |  |  |
| N |  |  |  |  |  |  |
| Mean (SD) |  |  |  |  |  |  |
| Median (Range) |  |  |  |  |  |  |
| Month 12 |  |  |  |  |  |  |
| N |  |  |  |  |  |  |
| Mean (SD) |  |  |  |  |  |  |
| Median (Range) |  |  |  |  |  |  |

Falls and syncope (number of falls, number of fallers/non-fallers, fall rate per person year, time to first fall, fall-related injuries, number of syncopal events)

Analysis Set 1 will be used for analysis. Data from all participants with at least one falls diary available will be reported according to their randomised treatment allocation.

The number of falls reported per participant will be tabulated as frequency and percentage and summarised as mean, SD, median and range. The total number of falls and total follow-up time from participants allocated to each randomised group will be reported. The simple, unadjusted incidence rate (IR) of falls per person year (i.e. total number of falls / total follow-up time) will be calculated in each randomised treatment group and presented with 95% CIs.

The same approach as above will be used for reporting of syncopal events, and a combined outcome of falls and syncopal events.

The number of fallers (i.e. the number of participants reporting at least one fall over the 12 month period) will be reported as frequency and percentage out of those returning at least one fall diary.

Time to first fall will be calculated and tabulated by whether this was within the first 3 months from randomisation, between 3 and 6 month follow-up or after 6 month follow-up.

Fall-related injuries will be tabulated by type, with the number of participants affected and the total number of occurrences reported by randomised treatment group. The number and proportion of participants reporting at least one fall-related injury will be tabulated by treatment group. The total number of fall-related injuries reported per participant will be tabulated as frequency and percentage and as mean, SD, median and range. The total number of fall-related injuries reported in participants allocated to each treatment group will be presented. For the purpose of calculating percentages the number of participants returning at least one fall diary will be used as the denominator.

**Example Table 8: Falls and syncopal events**

|  | Control | Fludrocortisone | Midodrine |
| --- | --- | --- | --- |
|  | N = | N = | N = |
| **Number of falls** |  |  |  |
| 0 |  |  |  |
| 1 |  |  |  |
| >2 |  |  |  |
| Mean (SD) |  |  |  |
| Median (Range) |  |  |  |
| **Fall rate** |  |  |  |
| Total number of falls |  |  |  |
| Total follow-up time (years) |  |  |  |
| Incidence rate (95% CI) |  |  |  |
| **Fallers / non-fallers** |  |  |  |
| ≥1 fall / N (%) |  |  |  |
| **Time to first fall** |  |  |  |
| < 3months |  |  |  |
| 3-6 months |  |  |  |
| >6 months |  |  |  |
| **Number of syncopal events** |  |  |  |
| 0 |  |  |  |
| 1 |  |  |  |
| >2 |  |  |  |
| Mean (SD) |  |  |  |
| Median (Range) |  |  |  |
| **Syncopal event rate** |  |  |  |
| Total number of syncopal events |  |  |  |
| Incidence rate (95% CI) |  |  |  |
| **Number of falls and syncopal events** |  |  |  |
| 0 |  |  |  |
| 1 |  |  |  |
| >2 |  |  |  |
| Mean (SD) |  |  |  |
| Median (Range) |  |  |  |
| **Falls and syncope event rate** |  |  |  |
| Total number of events |  |  |  |
| Incidence rate (95% CI) |  |  |  |

Standing blood pressure and postural blood pressure drop

The methods described below will be applied to the following variables; nadir standing systolic blood pressure (BP), nadir standing diastolic BP, systolic postural BP drop, diastolic postural BP drop.

Analysis Set 1 will be used for analysis. All data available will be reported according to randomised treatment allocation.

Each BP measurement and change from baseline will be summarised descriptively by randomised treatment group. The number with data, mean, standard deviation, median and range will be summarised at each visit.

**Example Table 9: Blood pressure**

| Blood pressure | Value at follow-up visit | | | Change from baseline | | |
| --- | --- | --- | --- | --- | --- | --- |
|  | Control | Fludrocortisone | Midodrine | Control | Fludrocortisone | Midodrine |
| **Nadir standing** |  |  |  |  |  |  |
| Month 3* |  |  |  |  |  |  |
| N |  |  |  |  |  |  |
| Systolic |  |  |  |  |  |  |
| Mean (SD) |  |  |  |  |  |  |
| Median (Range) |  |  |  |  |  |  |
| Diastolic |  |  |  |  |  |  |
| Mean (SD) |  |  |  |  |  |  |
| Median (Range) |  |  |  |  |  |  |
| **Postural drop** |  |  |  |  |  |  |
| Month 3* |  |  |  |  |  |  |
| N |  |  |  |  |  |  |
| Systolic |  |  |  |  |  |  |
| Mean (SD) |  |  |  |  |  |  |
| Median (Range) |  |  |  |  |  |  |
| Diastolic |  |  |  |  |  |  |
| Mean (SD) |  |  |  |  |  |  |
| Median (Range) |  |  |  |  |  |  |

**Data will be presented in the same way for Month 6 and Month 12*

Hospital admissions

Hospital admissions will be reported similarly to falls.

Analysis Set 1 will be used for analysis. Data from all participants with at least one review of hospital admissions available will be reported according to randomised treatment allocation.

The number of admissions reported per participant will be tabulated as frequency and percentage and summarised as mean, SD, median and range. Reasons for admission will also be tabulated as frequency and percentage. The total number of admissions and total follow-up time from participants allocated to each randomised group will be reported. The simple, unadjusted incidence rate of admissions per person year (i.e. total number of admissions / total follow-up time) will be calculated in each randomised treatment group and presented with 95% CIs.

#### 23. Additional / Exploratory analyses

No additional or exploratory analyses are planned. Details of exploratory analyses which would have been performed had the trial continued to full enrolment are described in the Appendix.

#### 24. Missing data

The availability of outcome data will be summarised as described in section 4.1.3. Given the small sample size we do not plan any imputation of missing data (except where part of questionnaire scoring). Methods which would have been employed had the trial continued are described in the Appendix.

### 25. **SAFETY**

#### 26. Adverse Events

Safety data will be reported in the Safety Analysis Set, with adverse events reported according to treatment received at the time of AE onset.

The following metrics will be used to provide an overall summary of adverse events reported while exposed to each treatment group, see Example Table 10:

- The number of participants exposed to each treatment group.
- The total exposure time (measured in years) to each treatment group, i.e. a sum across all participants of the amount of time they were exposed to each treatment group.
- Number and proportion of participants affected by at least one adverse event while exposed to each treatment group. Percentages will be calculated out of those exposed.
- Worst grade (mild, moderate, severe) adverse event reported while exposed to each treatment group. This will be tabulated with percentages calculated out of those exposed.
- Total number of adverse event occurrences reported by each participant while exposed to each treatment group. This will be tabulated (with percentages calculated out of those exposed) and/or reported as median and range, as appropriate
- Total number of unique adverse event occurrences (i.e. different preferred terms) reported by each participant while exposed to each treatment group. This will be tabulated (with percentages calculated out of those exposed) and/or reported as median and range, as appropriate
- Total number of adverse event occurrences reported across all participants while exposed to each treatment group. This will also be broken down by severity (mild, moderate, severe).
- The incidence rate (IR) of adverse event occurrence while exposed to each treatment group (i.e. total number of AE occurrences / total exposure time).

For each adverse event, at the preferred term level, we will also report the following metrics by treatment group, see Example Table 11:

- The number and proportion of participants experiencing the adverse event at any point while exposed to each treatment group. Percentages will be calculated out of those exposed.
- The worst reported severity (mild, moderate, severe) for the adverse event at any point while exposed to each treatment group. Percentages will be calculated out of those exposed.
- The total number of occurrences of the adverse event while exposed to each treatment group.
- The total number of occurrences of the adverse event, by severity (mild, moderate, severe), while exposed to each treatment group.

This data may also be presented graphically as dot plots of the proportion of participants affected by each adverse event. The number of participants affected and the total number of occurrences will be shown on the graph. See Example Figure 5. Had the trial continued to full enrolment the graph would have also shown a measure of relative risk with 95% CIs, with separate graphs produced for the comparison of fludrocortisone and control, midodrine and control, and fludrocortisone and midodrine. If there are many different adverse events reported we will focus on the graphical presentation of only those occurring in a specified proportion of participants (e.g. >3% participants in either group).

The same summaries will also be reported for adverse reactions (adverse events reported as possibly, probably or definitely related to trial treatment) and for non-serious events (as required for EudraCT reporting).

Adverse events which resulted in treatment discontinuation will also be tabulated. Percentages will be calculated out of those exposed to each treatment group.

**Example Table 10: Summary of adverse events**

|  | Control | Fludrocortisone | Midodrine |
| --- | --- | --- | --- |
| Number of participants exposed; N |  |  |  |
| Total exposure time (years); N |  |  |  |
| Number of AEs per participant; N (%) |  |  |  |
| 0 |  |  |  |
| 1-3 |  |  |  |
| 4-6 |  |  |  |
| >6 |  |  |  |
| Median (IQR); Range |  |  |  |
| Number of unique AEs per participant; N (%) |  |  |  |
| 0 |  |  |  |
| 1-3 |  |  |  |
| 4-6 |  |  |  |
| >6 |  |  |  |
| Median (IQR); Range |  |  |  |
| Worst grade reported across all AEs; N (%) |  |  |  |
| *Mild* |  |  |  |
| *Moderate* |  |  |  |
| *Severe* |  |  |  |
| Total number of AEs reported; N |  |  |  |
| *Mild* |  |  |  |
| *Moderate* |  |  |  |
| *Severe* |  |  |  |
| Incidence of AEs; IR (95% CI) |  |  |  |

**Example Table 11: Summary of adverse events by type**

| Adverse event term | | Number (%) of participants affected^1,2^ | | | Number of occurrences^1^ | | |
| --- | --- | --- | --- | --- | --- | --- | --- |
|  |  | Control | Fludrocortisone | Midodrine | Control | Fludrocortisone | Midodrine |
|  |  | N = | N = | N = | N = | N = | N = |
| Nausea | Overall |  |  |  |  |  |  |
|  | *Mild* |  |  |  |  |  |  |
|  | *Moderate* |  |  |  |  |  |  |
|  | *Severe* |  |  |  |  |  |  |
| Headache | Overall |  |  |  |  |  |  |
|  | *Mild* |  |  |  |  |  |  |
|  | *Moderate* |  |  |  |  |  |  |
|  | *Severe* |  |  |  |  |  |  |
| AE 1 | Overall |  |  |  |  |  |  |
|  | *Mild* |  |  |  |  |  |  |
|  | *Moderate* |  |  |  |  |  |  |
|  | *Severe* |  |  |  |  |  |  |
| AE 2 | Overall |  |  |  |  |  |  |
|  | *Mild* |  |  |  |  |  |  |
|  | *Moderate* |  |  |  |  |  |  |
|  | *Severe* |  |  |  |  |  |  |

*^1^While exposed to each treatment group*

*^2^% calculated out of total number exposed. If >1 occurrence per participant the worst reported severity is tabulated*

**Example Figure 5: Proportion of participants affected by each AE**

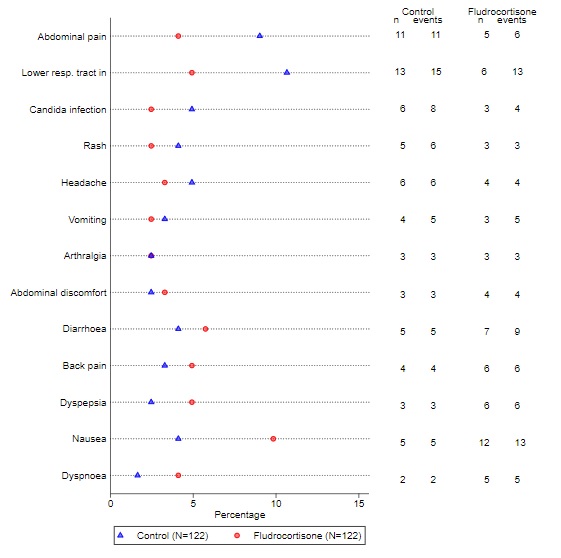

#### 27. Serious Adverse Events

A chronological listing of reportable serious adverse events (SAEs) will be presented, see Example Table 12.

The following metrics will also be reported:

- The number and proportion of participants reporting at least one SAE while exposed to each treatment group. Percentages will be calculated out of those exposed.
- The total number of occurrences of SAEs, across all participants, while exposed to each treatment group.
- For each SAE (using the preferred term), the total number of participants affected and the total number of occurrences while exposed to each treatment group.

**Example Table 12: Line listing of all SAEs**

| ID | SAE no. | Rand. group | IMP at onset | IMP start + end dates | Description | SAE onset date | Severity^1^ | Causality^2^ | Serious criteria^3^ | Outcome^4^ | Outcome date |
| --- | --- | --- | --- | --- | --- | --- | --- | --- | --- | --- | --- |

*^1^Mild; moderate; severe*

*^2^Related; unrelated; unable to determine*

*^3^Resulted in death; Life threatening; Inpatient hospitalisation / prolonged hospitalisation; Persistent or significant disability/incapacity; Congenital anomaly / birth defect; Important medical event*

*^4^Recovered; Condition improved; Condition deteriorated; Condition unchanged; Participant died; Recovered with sequelae; Condition stable and no change anticipated*

Non-reportable serious adverse events will also be summarised.

#### 28. Other safety measures

We do not plan analyses of any other safety measures (e.g. laboratory values, vital signs etc.).

### 29. **statistical software**

Data will be exported from the electronic Clinical Data Management System (CDMS) into a STATA format by the NCTU Data(base) Manager at time points agreed by the Trial Management Group. Statistical analyses will be carried out by the Trial Statistician at the Biostatistics Research Group predominately using Stata but R software may also be used. All programs and output will be stored in the School Statistics folder on the IHS server.

### **APPENDIX**

The planned analysis methods which would have been used had the trial continued to full enrolment are summarised below for both the interim and final analysis.

A. **Interim analysis**

A linear mixed-effects model would have been fitted to three and six month OHQ data, with fixed effects for baseline OHQ, age (<80 years or ≥80), and aetiology (neurogenic or non-neurogenic OH), and a treatment-by-time interaction term. We planned to include random effects for site and individual (nested within site), however if there were sites with few participants (<5) then site would have been excluded from the model.

Where $Y_{ij}$ denotes the OHQ score for participant $i$ at time $j$, the planned interim analysis model would be:

$$Y_{ij}=\beta_{0}+\beta_{1}{Trt1}_{i}+\beta_{2}{Trt2}_{i}+\beta_{3}{OHQ}_{i}^{0}+\beta_{4}{Age}_{i}+\beta_{5}{Aetiology}_{i}+\beta_{6}M6+\beta_{7}M6*{Trt1}_{i}+\beta_{8}M6*{Trt2}_{i}+b_{1,i}+{b_{2,k}+e}_{ij}$$

Where ${Trt1}_{i}$ and ${Trt2}_{i}$ are dummy variables for treatment allocation (${Trt1}_{i}$: 1 if allocated to conservative management plus fludrocortisone, 0 otherwise; ${Trt2}_{i}$: 1 if allocated to conservative management plus midodrine, 0 otherwise), ${OHQ}_{i}^{0}$ is the baseline OHQ score for participant $i$, $M6$ is a dummy variable for time (0 or 1) at month six. Note month three is represented by $M6$ = 0. $b_{1,i}$ and $b_{2,k}$ are random intercepts at the participant and site level respectively and are assumed to follow a normal distribution.

This model would have been fitted using restricted maximum likelihood (REML) and an unstructured covariance matrix. The following code in Stata would have been used:

mixed ohq i.arm##i.visit ohq_bl i.age i.aetiology || site: || id:, residuals(unstructured, t(visit)) reml

This model accounts for missing outcome data under a missing at random (MAR) assumption. Participants would be included in the model if they had at least one OHQ score available from the three- or six-month follow-up visit.

From this model we would obtain the estimated mean difference in six-month OHQ between fludrocortisone and control ($\beta_{1}$+$\beta_{7}$) and between midodrine and control ($\beta_{2}$+$\beta_{8}$). This would be reported with a corresponding 95% confidence interval and p-value. The Wald test would be used to test the null hypothesis of no treatment difference at six months for each experimental treatment against control (reference). The following Stata code would be used:

lincom 1.arm + 2.visit#1.arm

lincom 2.arm + 2.visit#2.arm

If an experimental treatment arm showed no benefit compared to the control arm, i.e. if the estimated mean difference for an experimental arm was 0 or more (corresponding to the arm having worse outcome than control), it would be recommended to be dropped from the trial. This is equivalent to a one-side p-value > 0.5. Otherwise the trial would be recommended to continue as planned.

The table below shows the chance of each arm being stopped for lack of benefit at the interim analysis.

| Scenario | Probability fludrocortisone stops at interim | Probability midodrine stops at interim |
| --- | --- | --- |
| Mean effect of both = 0 (null scenario) | 50.0% | 50.0% |
| Fludrocortisone has MCID, midodrine effect = 0 | 2.1% | 50.0% |
| Midodrine has MCID, fludrocortisone effect = 0 | 50.0% | 2.1% |
| Both treatments have MCID effect | 2.1% | 2.1% |

Any recommendation of the trial design to drop an experimental arm would be ratified by the DMC and the trial steering committee (TSC). A meeting of the DMC would have been held at the time of the interim analysis. The DMC would review the results of the interim analysis in context of other accumulating data such as adverse events, treatment crossovers and withdrawals. The DMC would make a recommendation, considering all aspects of the trial, on whether any experimental arm should be dropped at the interim analysis. The recommendation of the DMC would be passed onto the TSC who would then make a decision. Any decision to overrule the recommendation of the design would need to be clearly justified and documented. Unless the DMC specifically requested the release of some unblinded information, the TSC would not have access to any unblind outcome data.

If one experimental arm was recommended to be dropped at the interim analysis, and this was ratified by the oversight committees, the trial would continue to its planned enrolment of 366, but subsequent participants would be randomised in a 1:1 ratio between control and the remaining experimental arm. If both experimental arms were recommended for dropping at the interim analysis and this was ratified by the oversight committees, the trial would be terminated early. Recruitment would continue to all three arms while the interim analysis was being conducted.

It was planned that the interim analysis would have been conducted by a statistician (a member of the Biostatistics Research Group) not otherwise involved in the study and reviewed by the Lead Statistician.

B. **Final analysis**
   a. **Primary outcome**
      i. **Main analysis methods**

Analysis Set 1 would have been used for analysis (see Section 3.4). All participants would have been analysed according to their randomised treatment allocation and all available outcome data included in the analysis.

Using the estimand framework [9], we were primarily interested in the effect of being allocated to fludrocortisone or midodrine in addition to conservative management, regardless of treatment discontinuation or any other treatments received, i.e. a treatment policy estimand. The intended main primary outcome estimand is described in the table below:

| Estimand attribute | Description |
| --- | --- |
| Population | Patients with symptomatic OH refractory to lifestyle modification and meeting the CONFORM-OH eligibility criteria |
| Treatment | Allocated to six months of treatment with conservative management plus fludrocortisone or conservative management plus midodrine compared to conservative management alone |
| Outcome variable | Overall composite OHQ score at month six |
| Strategies used to handle intercurrent events | - Discontinuation of allocated treatment – treatment policy^1^ - Use of non-allocated treatment with fludrocortisone or midodrine – treatment policy^1^ |
| Population-level summary measure | Mean difference in OHQ score at six months (adjusted for baseline) between the intervention and control groups |

*^1^ A* ***treatment policy*** *strategy considers the occurrence of the intercurrent event as irrelevant, and participant data are analysed regardless*

The OHQ score would be summarised descriptively by randomised treatment group. The number with data, mean, standard deviation and the median, IQR and range would be summarised at each visit. Data would also be presented graphically by randomised treatment group as the mean value with 95% CIs at each visit, see Appendix Figure 1. The OHSAS and OHDAS would have also been summarised descriptively.

**Appendix Figure 1: OHQ score over the 12 month follow-up period (note this is not genuine data)**

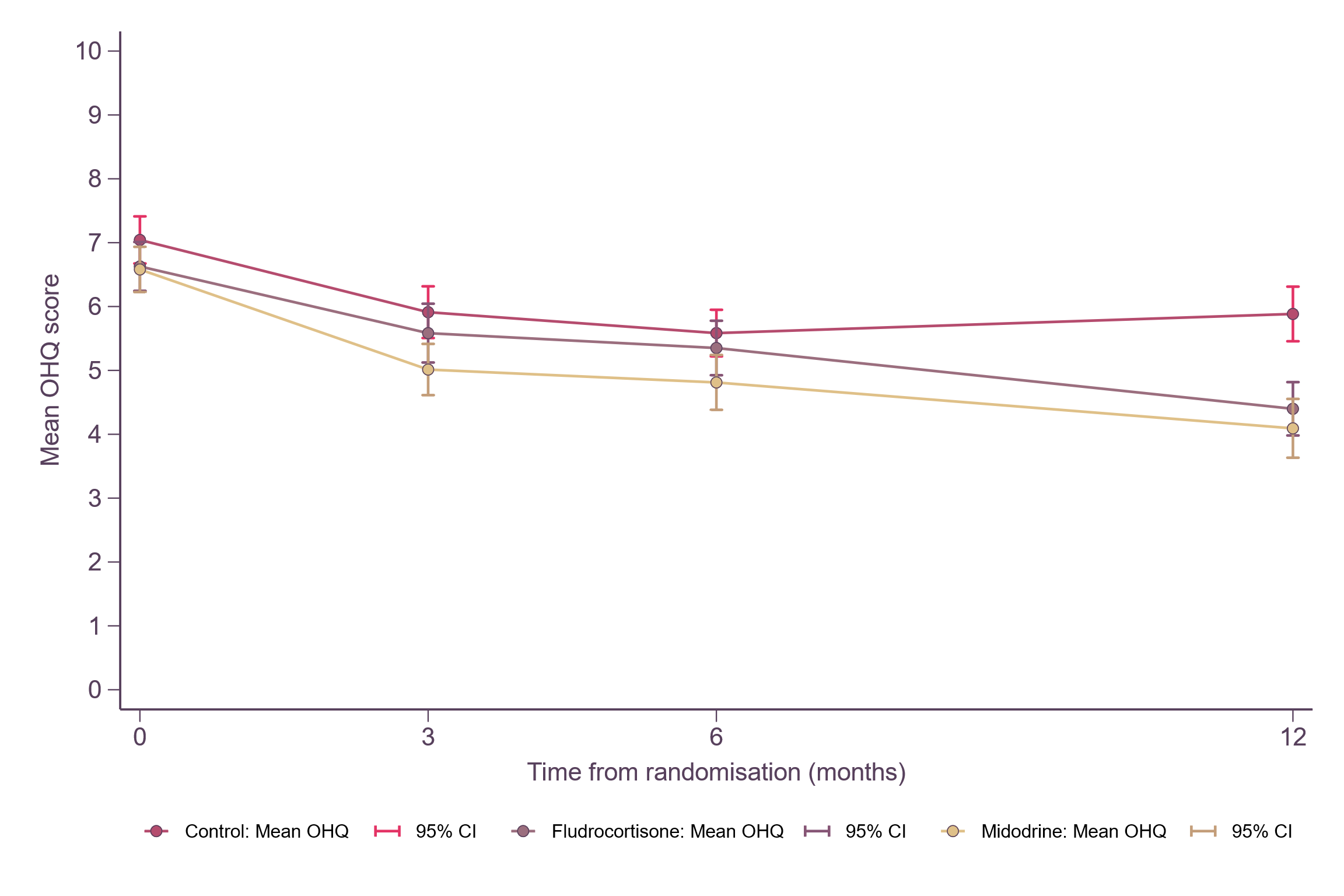

For the primary analysis of the overall OHQ score at six-months, a linear mixed-effects model would have been fitted to the three-and six-month OHQ data. Fixed effects would include baseline OHQ, baseline age (<80 years or >=80), aetiology (neurogenic or non-neurogenic) and a treatment-by-time interaction term. Random effects would include site and individual (nested within site).

Where $Y_{ijk}$ denotes the OHQ score for participant $i$ at time $j$ from site $k$, the primary analysis model would be:

$$Y_{ijk}=\beta_{0}+\beta_{1}{Trt1}_{i}+\beta_{2}{Trt2}_{i}+\beta_{3}{OHQ}_{i}^{0}+\beta_{4}{Age}_{i}+\beta_{5}{Aetiology}_{i}+\beta_{6}M6+\beta_{7}M6*{Trt1}_{i}+\beta_{8}M6*{Trt2}_{i}+b_{1,i}+b_{2,k}+e_{ijk}$$

Where ${Trt1}_{i}$ and ${Trt2}_{i}$ are dummy variables for treatment allocation (${Trt1}_{i}$: 1 if allocated to conservative management plus fludrocortisone, 0 otherwise; ${Trt2}_{i}$: 1 if allocated to conservative management plus midodrine, 0 otherwise), ${OHQ}_{i}^{0}$ is the baseline OHQ score for participant $i$, $M6$ is a dummy variable for time (0 or 1) at month six. Note month three is represented by $M6$ = 0. $b_{1,i}$ and $b_{1,k}$ are random intercepts at the participant and site level respectively and are assumed to follow normal distributions.

This model would be fitted using restricted maximum likelihood (REML) and an unstructured covariance matrix. The following code in Stata would have been used:

mixed ohq i.arm##i.visit ohq_bl i.age i.aetiology || site: || id:, residuals(unstructured, t(visit)) reml

Missing responses would be assumed to be missing at random (MAR). Participants would be included in the model if they had at least one OHQ score available from the three or six month follow-up visit.

From this model we would obtain the estimated mean difference in six-month OHQ between fludrocortisone and control ($\beta_{1}$+$\beta_{7}$) and between midodrine and control ($\beta_{2}$+$\beta_{8}$). This will be reported with a corresponding 95% confidence interval and p-value, see Appendix Table 1. The Wald test would be used to test the null hypothesis of no treatment difference at six months for each experimental treatment against control (reference). The following Stata code would be used:

lincom 1.arm + 2.visit#1.arm

lincom 2.arm + 2.visit#2.arm

We would have investigated the assumptions of the primary analysis model using the following methods:

- Plot of residuals versus fitted values
- Normal quantile plot of standardised residuals
- Normal quantile plot of the standardised predicted random effects

If the model failed to converge we would instead treat site as a fixed effect. Or, if there were sites with few participants (<5) then site would be excluded from the model. This may have been an issue particularly if recruitment to one (or all) arms is terminated at the interim analysis time point, resulting in a relatively small sample size (minimum 67 per arm).

**Appendix Table 1: Primary outcome, OHQ**

| OHQ score | Control | Fludrocortisone | Midodrine | Mean difference (MD) | |
| --- | --- | --- | --- | --- | --- |
|  |  |  |  | Fludrocortisone - control | Midodrine - control |
|  | N; Mean (SD) | N; Mean (SD) | N; Mean (SD) | MD (95% CI); p-value | MD (95% CI); p-value |
| Month 3 |  |  |  |  |  |
| Month 6* |  |  |  |  |  |
| Month 12 |  |  |  |  |  |

**Primary outcome*

ii. **Subgroup analyses**

We would have explored whether the treatment effect for the 6-month OHQ score was consistent across subgroups. To do this a linear mixed-effects regression model would be fitted to the 6-month OHQ data. Fixed effects would include treatment group, baseline OHQ, baseline age (<80 years or >=80) and aetiology (neurogenic or non-neurogenic), and an interaction between treatment and the subgroup variable of interest. Site would be included as a random effect.

For each subgroup, the estimated mean difference (and 95% CI) in six-month OHQ between fludrocortisone and control and between midodrine and control would be presented using Forest plots which would also show the p-value from a test of interaction. Subgroups we would have considered were:

- Neurogenic OH versus non-neurogenic OH
- Age ≥80 years versus age <80 years
- Male versus female
- Presence of diabetes versus no diabetes***

**Subject to sufficient numbers within each group*

As the trial had not been powered to detect subgroup effects these results would have been considered exploratory and hypothesis generating. Conclusions would not have been drawn on the basis of these results.

Should any model have failed to converge we would instead have included site as a fixed effect, or excluded it from the model, as described above (*i. Main analysis methods)*.

iii. **Sensitivity analyses**

To assess the robustness of the results (i.e. that inferences do not change) we would conduct the following sensitivity analyses, which would target the same treatment policy estimand:

1) The primary analysis is valid under the assumption that data are missing at random. If the OHQ score is missing at six months in more than >15% of participants (the assumed attrition rate), either overall or in any arm, we would perform sensitivity analyses using multiple imputation methods to explore the robustness of the results to the MAR assumption. This is discussed in more detail below (*d. Missing data*).
2) The main analysis would use all OHQ data reported at the three, six and 12-month visits. If >10% of questionnaires, either overall or within any arm, were completed outside of the protocol specified visit windows we would repeat the analysis excluding these data.
   iv. **Supplementary analyses**

A supplementary analysis would be performed targeting an ‘on-treatment’ or hypothetical estimand; the treatment effect if all participants had continued on their allocated treatment. The key aspects of this estimand are described in the table below:

| Estimand attribute | Description |
| --- | --- |
| Population | Patients with symptomatic OH refractory to lifestyle modification and meeting the CONFORM-OH eligibility criteria |
| Treatment | Six months of treatment with conservative management plus fludrocortisone or conservative management plus midodrine compared to conservative management alone |
| Outcome variable | Overall composite OHQ score at month six |
| Strategies used to handle intercurrent events | - Discontinuation of allocated treatment – hypothetical^1^ - Use of non-allocated treatment with fludrocortisone or midodrine – hypothetical^1^ |
| Population-level summary measure | Mean difference in OHQ score at six months (adjusted for baseline) between the intervention and control groups |

*^1^ Under a* ***hypothetical*** *strategy we are interested in the value the outcome variable would have taken had the intercurrent event not happened*

This analysis would be performed using Analysis Set 2 (see Section 3.4). Participants would be analysed according to their allocated treatment group but data collected after discontinuation of allocated treatment or initiation of non-allocated fludrocortisone or midodrine would be set as missing. This dataset would be analysed using the same linear mixed-model as described above (*i. Main analysis methods)*. The model assumes data are missing at random, i.e. that the missing data would be similar to that of other participants in the same treatment group with similar baseline characteristics.

While the missing at random assumption is plausible under hypothetical, on-treatment conditions, it is possible participants who discontinued allocated treatment or initiated non-allocated fludrocortisone or midodrine would have had worse outcomes had they continued on allocated treatment than similar participants who did not experience these intercurrent events. We would therefore also perform sensitivity analyses implementing controlled multiple imputation (MI) using a δ-based pattern-mixture approach. This is discussed in more detail below (*d. Missing data*).

b. **Secondary outcomes**

All secondary outcomes would have been analysed using treatment policy estimands.

OHQ score at Month 12

Analysis Set 1 would be used for analysis. All participants with OHQ data available at three, six or 12-month time points would be analysed according to their randomised treatment allocation.

The estimated mean difference in 12-month OHQ would be estimated by fitting a liner mixed-effects model, as described above (*i. Main analysis methods*) but to three-, six- and 12-month outcome data. Where $Y_{ijk}$ denotes the OHQ score for participant $i$ at time $j$ from site $k$, the analysis model would be:

$$Y_{ijk}=\beta_{0}+\beta_{1}{Trt1}_{i}+\beta_{2}{Trt2}_{i}+\beta_{3}{OHQ}_{i}^{0}+\beta_{4}Age+\beta_{5}Aetiology+\beta_{6}M6+\beta_{7}M6*{Trt1}_{i}+\beta_{8}M6*{Trt2}_{i}+\beta_{9}M12+\beta_{10}M12*{Trt1}_{i}+\beta_{11}M12*{Trt2}_{i}+b_{1,i}+b_{2,k}+e_{ijk}$$

Where $M6$ is a dummy variable for time (0 or 1) at month six,$M12$ is a dummy variable for time (0 or 1) at month 12. Note month three is represented by $M6$ = 0 and $M12$ = 0. All other parameters are as previously described.

Activities of daily living (ADLs) measured by the Nottingham Extended ADL scale

Analysis Set 1 would be used for analysis. All participants with data available would be analysed according to their randomised treatment allocation.

The NEADL score would be summarised descriptively by randomised treatment group. The number with data, mean, standard deviation and the median, IQR and range would be summarised at each visit. Data would also be presented graphically by randomised treatment group as the mean value with 95% CIs at each visit.

As for the OHQ data, a linear mixed-effects model would be fitted to the three, six and 12 month data. Fixed effects would include baseline NEADL score, baseline age (<80 years or >=80), aetiology (neurogenic or non-neurogenic) and a treatment assignment x time interaction term. Random effects would include site and individual (nested within site). This model would be fitted using REML and an unstructured covariance matrix.

Missing responses would be assumed to be missing at random (MAR). Participants would be included in the model if they had at least one NEADL score available from the three, six or 12 month follow-up visit.

From this model we would obtain the estimated mean difference in the three, six and 12-month follow-up NEADL score between fludrocortisone and control and between midodrine and control. This will be reported with a corresponding 95% confidence interval, see Appendix Table 2.

Assumptions of the model would be investigated as described for the analysis of the primary outcome.

**Appendix Table 2: NEADL**

| NEADL score | Control | Fludrocortisone | Midodrine | Mean difference (MD) | |
| --- | --- | --- | --- | --- | --- |
|  |  |  |  | Fludrocortisone - control | Midodrine - control |
|  | N; Mean (SD) | N; Mean (SD) | N; Mean (SD) | MD (95% CI);  p-value | MD (95% CI) ;  p-value |
| Month 3 |  |  |  |  |  |
| Month 6 |  |  |  |  |  |
| Month 12 |  |  |  |  |  |

Falls and syncope (number of falls, number of fallers/non-fallers, fall rate per person year, time to first fall, fall-related injuries, number of syncopal events)

Analysis Set 1 would be used for analysis. All participants with at least one falls diary available would be analysed according to their randomised treatment allocation.

The number of falls reported per participant would be tabulated as frequency and percentage and summarised as mean, SD, median, IQR and range. The number, or percentage, of falls may have also been presented graphically, see Appendix Figure 2. The total number of falls and total follow-up time from participants allocated to each randomised group would be reported. The simple, unadjusted incidence rate (IR) of falls per person year (i.e. total number of falls / total follow-up time) would be calculated in each randomised treatment group and presented with 95% CIs.

A mixed-effects negative binomial regression model would be fitted to the falls data. Site would be included as a random effect and treatment assignment, baseline age (<80 years or >=80) and aetiology (neurogenic or non-neurogenic) as fixed effects. The log of the exposure variable (follow-up time) would be included in the model with the coefficient constrained to be one. From this model we would obtain the estimated incidence rate ratio (IRR) and risk difference in the fall rate per person-year between fludrocortisone and control and midodrine and control. These estimates would be reported with corresponding 95% confidence intervals.

We may instead have used a Poisson or zero-inflated model if this was more suitable for the distribution of the data. We would explore this graphically by plotting the observed and predicted number of events using each model, using the Pearson chi-square goodness of fit test and using Akaike's and Bayesian information criterion (AIC and BIC).

The same approach as above would be used for the analysis and reporting of syncopal events, and a combined outcome of falls and syncopal events.

The number of fallers (i.e. the number of participants reporting at least one fall over the 12 month period) would be reported as frequency and percentage out of those returning at least one fall diary. A mixed-effects Poisson regression model would be fitted to this binary outcome measure. Site would be included as a random effect and treatment assignment, baseline age (<80 years or >=80) and aetiology (neurogenic or non-neurogenic) as fixed effects. The log of the exposure variable (follow-up time) would be included in the model with the coefficient constrained to be one. From this model we would obtain the incidence rate ratio for the treatment effect comparing fludrocortisone and control and midodrine and control. These estimates would be reported with corresponding 95% confidence intervals.

Time to first fall would be summarised graphically using Kaplan-Meier curves, see Appendix Figure 3. A Cox proportional hazards regression model with mixed effects (i.e. a Cox regression model with shared frailty) would be fitted to the time to first fall data. Treatment assignment, baseline age (<80 years or >=80) and aetiology (neurogenic or non-neurogenic) would be included as fixed effects and site as a shared frailty term. Shared frailty terms will be assumed to follow a gamma distribution. From this model we would obtain the hazard ratio (HR) for the treatment effect comparing fludrocortisone and control and midodrine and control. These estimates would be reported with corresponding 95% confidence intervals. The proportional hazards assumption would be investigated using Schoenfeld residuals and by testing the interaction of treatment assignment with log(survival time). If the proportional hazards assumption was violated an alternative approach, such as the restricted mean survival time, would be used.

Fall-related injuries would be tabulated by type, with the number of participants affected and the total number of occurrences reported by randomised treatment group. The number and proportion of participants reporting at least one fall-related injury would be tabulated by treatment group. The total number of fall-related injuries reported per participant would be tabulated as frequency and percentage and as mean, SD, median, IQR and range. The total number of fall-related injuries reported in participants allocated to each treatment group would be presented. For the purpose of calculating percentages the number of participants returning at least one fall diary would be used as the denominator.

**Appendix Table 3: Falls and syncopal events**

|  | Control | Fludrocortisone | Midodrine |
| --- | --- | --- | --- |
|  | N = | N = | N = |
| **Number of falls** |  |  |  |
| 0 |  |  |  |
| 1 |  |  |  |
| >2 |  |  |  |
| Mean (SD) |  |  |  |
| Median (IQR) |  |  |  |
| Range |  |  |  |
| **Fall rate** |  |  |  |
| Total number of falls |  |  |  |
| Total follow-up time (years) |  |  |  |
| Incidence rate (95% CI) |  |  |  |
| Absolute difference* (95% CI) | NA |  |  |
| Incidence Rate Ratio* (95% CI) | NA |  |  |
| **Fallers / non-fallers** |  |  |  |
| ≥1 fall / N (%) |  |  |  |
| Incidence Rate Ratio* (95% CI) | NA |  |  |
| **Number of syncopal events** |  |  |  |
| 0 |  |  |  |
| 1 |  |  |  |
| >2 |  |  |  |
| Mean (SD) |  |  |  |
| Median (IQR) |  |  |  |
| Range |  |  |  |
| **Syncopal event rate** |  |  |  |
| Total number of syncopal events |  |  |  |
| Incidence rate (95% CI) |  |  |  |
| Absolute difference* (95% CI) | NA |  |  |
| Incidence Rate Ratio* (95% CI) | NA |  |  |
| **Number of falls and syncopal events** |  |  |  |
| 0 |  |  |  |
| 1 |  |  |  |
| >2 |  |  |  |
| Mean (SD) |  |  |  |
| Median (IQR) |  |  |  |
| Range |  |  |  |
| **Falls and syncope event rate** |  |  |  |
| Total number of events |  |  |  |
| Incidence rate (95% CI) |  |  |  |
| Absolute difference* (95% CI) |  |  |  |
| Incidence Rate Ratio* (95% CI) |  |  |  |

**compared to control*

**Appendix Figure 2: Number of falls (note this is not genuine data)**

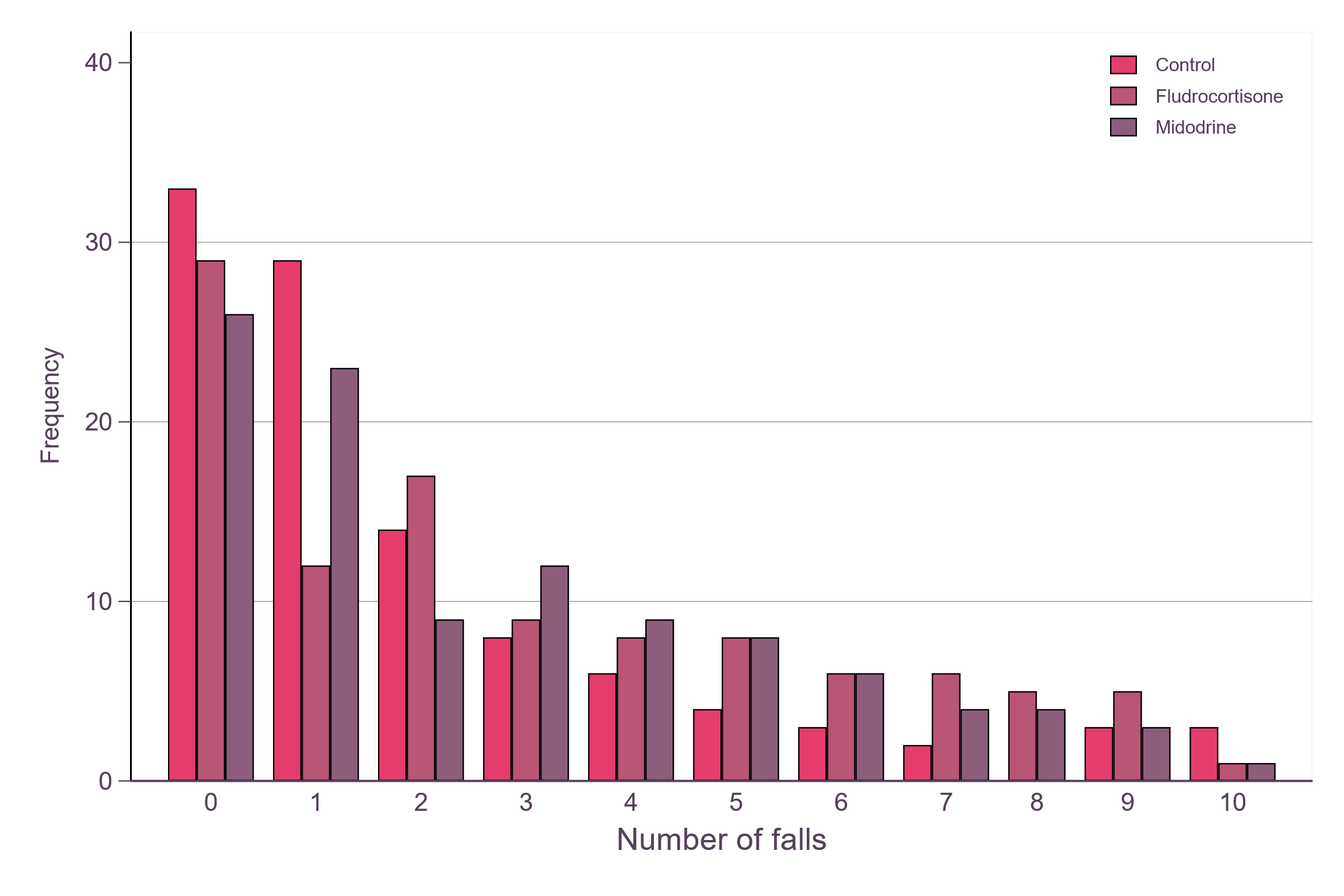

**Appendix Figure 3: Time to first fall (note this is not genuine data)**

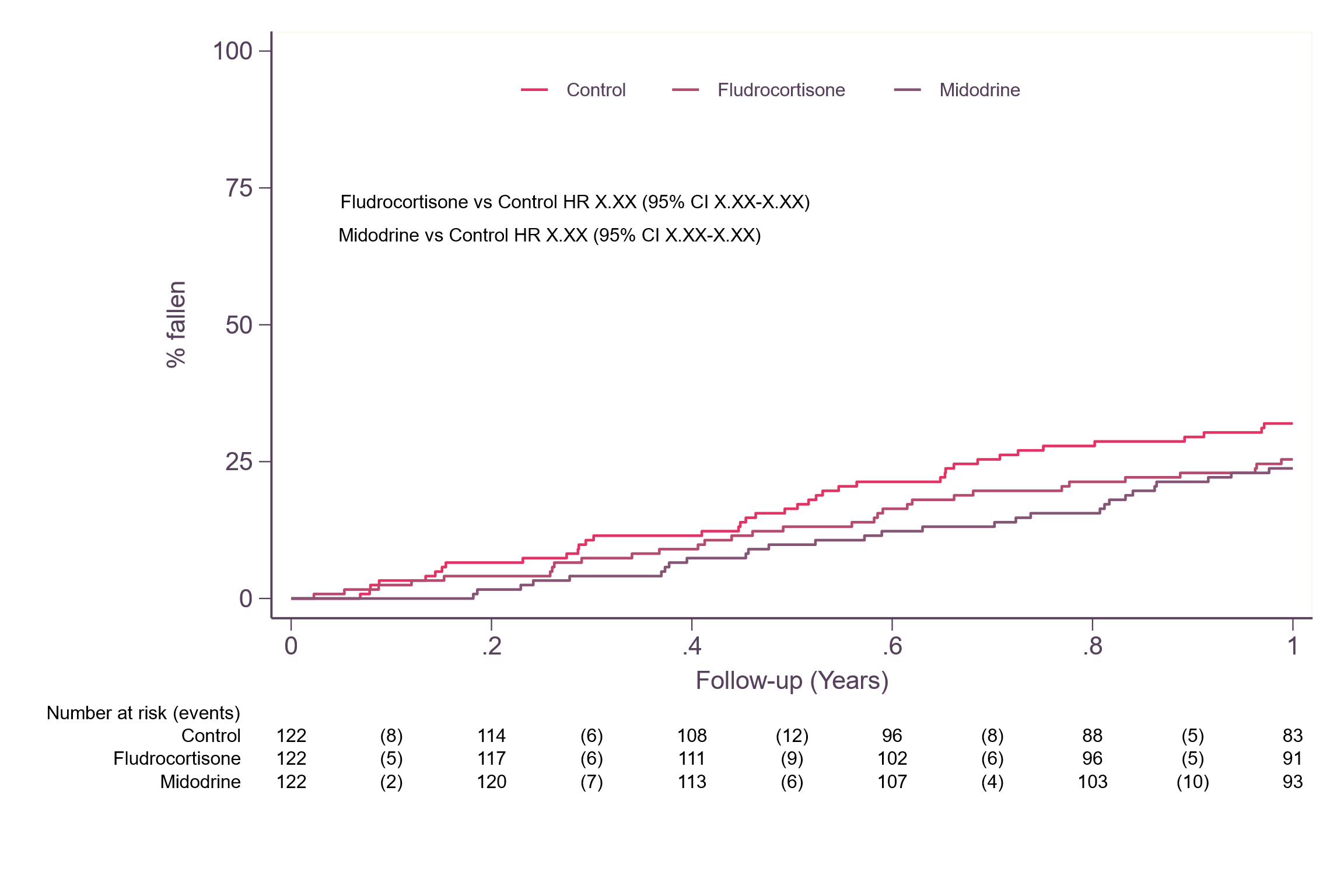

Standing blood pressure and postural blood pressure drop

The analysis methods described below would be applied to the following variables; nadir standing systolic blood pressure (BP), nadir standing diastolic BP, systolic postural BP drop, diastolic postural BP drop.

Analysis Set 1 would be used for analysis. All participants with data available would be analysed according to their randomised treatment allocation.

Each BP measurement would be summarised descriptively by randomised group. The number with data, mean, standard deviation and the median, IQR and range would be summarised at each visit. Data may have also been presented graphically by randomised treatment group as the mean value with 95% CIs at each visit.

As for the OHQ data, a linear mixed-effects model would have been fitted to the three, six and 12 month data. Fixed effects would include baseline BP measurement, baseline age (<80 years or >=80), aetiology (neurogenic or non-neurogenic) and a treatment assignment x time interaction term. Random effects would include site and individual (nested within site). This model would be fitted using REML and an unstructured covariance matrix.

Missing responses would be assumed to be missing at random (MAR). Participants would be included in the model if they had at least one BP measurement available from the three, six or 12 month follow-up visit.

From this model we would obtain the estimated mean difference in the three, six and 12-month follow-up BP measurements between fludrocortisone and control and between midodrine and control. This would be reported with a corresponding 95% confidence interval, see Appendix Table 4.

Assumptions of the model would be investigated as described for the analysis of the primary outcome.

**Appendix Table 4: Blood pressure**

| Blood pressure | Control | Fludrocortisone | Midodrine | Mean difference (MD) | |
| --- | --- | --- | --- | --- | --- |
|  |  |  |  | Fludrocortisone - control | Midodrine - control |
|  | N; Mean (SD) | N; Mean (SD) | N; Mean (SD) | MD (95% CI) ; p-value | MD (95% CI) ; p-value |
| **Nadir standing** |  |  |  |  |  |
| Month 3 |  |  |  |  |  |
| Systolic |  |  |  |  |  |
| Diastolic |  |  |  |  |  |
| Month 6 |  |  |  |  |  |
| Systolic |  |  |  |  |  |
| Diastolic |  |  |  |  |  |
| Month 12 |  |  |  |  |  |
| Systolic |  |  |  |  |  |
| Diastolic |  |  |  |  |  |
| **Postural drop** |  |  |  |  |  |
| Month 3 |  |  |  |  |  |
| Systolic |  |  |  |  |  |
| Diastolic |  |  |  |  |  |
| Month 6 |  |  |  |  |  |
| Systolic |  |  |  |  |  |
| Diastolic |  |  |  |  |  |
| Month 12 |  |  |  |  |  |
| Systolic |  |  |  |  |  |
| Diastolic |  |  |  |  |  |

Hospital admissions

Hospital admissions would be analysed similarly to falls.

Analysis Set 1 would be used for analysis. All participants with at least one review of hospital admissions available would be analysed according to their randomised treatment allocation.

The number of admissions reported per participant would be tabulated as frequency and percentage and summarised as mean, SD, median, IQR and range. Reasons for admission would also be tabulated as frequency and percentage. The number, or percentage, of admissions may also have been presented graphically. The total number of admissions and total follow-up time from participants allocated to each randomised group would be reported. The simple, unadjusted incidence rate of admissions per person year (i.e. total number of admissions / total follow-up time) would be calculated in each randomised treatment group and presented with 95% CIs.

A mixed-effects negative binomial regression model would be fitted to the admissions data. Site would be included as a random effect and treatment assignment, baseline age (<80 years or >=80) and aetiology (neurogenic or non-neurogenic) as fixed effects. The log of the exposure variable (follow-up time) would be included in the model with the coefficient constrained to be one.

From this model we would obtain the estimated incidence rate ratio and risk difference in the admission rate per person-year between fludrocortisone and control and midodrine and control. These estimates would be reported with corresponding 95% confidence intervals.

As for falls data, a Poisson or zero-inflated model may have been used if this was more suitable for the distribution of the data.

c. **Additional / Exploratory analyses**

A pre-specified exploratory analysis would compare the clinical effectiveness of fludrocortisone with midodrine for the primary outcome. This would use the same model as for the primary analysis but with fludrocortisone as the reference group.

d. **Missing data**
   i. **Intention-to-treat / treatment policy estimand**

The availability of outcome data would be summarised as described in section 4.1.3.

The characteristics of participants with and without a six month OHQ score would be explored. Characteristics to be summarised would have included baseline OHQ score, age, aetiology, gender, three month OHQ score (where available) and postural blood pressure drop (where available).

If more than 15% of participants had missing primary outcome data (OHQ score at six months), either overall or in any arm, then a sensitivity analysis of the primary outcome would be undertaken to explore the impact of departures from the MAR assumption, i.e. that the data are missing not at random (MNAR).

To do this we would implement reference-based multiple imputation (MI) using a jump-to-reference approach, following the guide proposed by Cro *et al* [10]. Missing data would be imputed assuming participants jump to behave like the usual care arm following their last observed timepoint. This would provide a conservative approach to missing data under a MNAR assumption. Note that any interim missing values would be imputed under a MAR assumption.

The variables to be included in the imputation model would be the same as those included in the primary analysis model. We would initially plan to impute 50 datasets, which would be very likely to be higher than the simple rule of thumb of one imputation per percent of missing data [11]. However, the number of imputations may have been increased if 50 imputations was not felt to provide adequate precision. Each imputed dataset would be analysed using the primary analysis model and Rubin’s rules used to combine treatment estimates across datasets to give a single value [12].

ii. **‘On-treatment’ / hypothetical estimand**

For the ‘on-treatment’ estimand we would implement controlled multiple imputation (MI) using a δ-based pattern-mixture approach, again following the guide proposed by Cro *et al* [8]. Briefly, missing primary outcome data would be imputed under the assumption that it is MAR, conditional on the other variables in the imputation model. The variables in the imputation model would be the same as those included in the primary analysis model. We would initially plan to impute 50 datasets, which would be very likely to be higher than the simple rule of thumb of one imputation per percent of missing data [11]. However, the number of imputations may have been increased if 50 imputations was not felt to provide adequate precision. A fixed value, δ, would then be added to the imputed values to increase the mean response beyond that predicted under MAR. Each imputed dataset would be analysed using the primary analysis model and Rubin’s rules used to combine treatment estimates across datasets to give a single value [12]. We planned to use the following values of δ: +0.5, +1.0, +1.5, +2.0 and +2.5, but bounded to lie in the range 0 to 10. For example, for δ = 2.0 implies a missing OHQ score would be 2.0 points higher than expected if the data were missing at random. We would also explore how extreme a value a value of δ (positive or negative) would be needed to change the interpretation of the results (i.e. from *p* < 0.05 to *p* ≥ 0.05 or vice-versa); this ‘tipping point’ value for δ would be reported and consideration given to how realistic or plausible this value would be in practice.

For each value of δ we would report the estimated mean difference in six-month OHQ between each experimental arm and control, along with 95% CIs and corresponding p-value, e.g. see Appendix Table 4 below.

**Appendix Table 4: Exploring the impact of data being MNAR for the ‘on-treatment’ analysis of OHQ data**

|  | Mean difference (MD) | |
| --- | --- | --- |
|  | Fludrocortisone - control | Midodrine - control |
|  | MD (95% CI); p-value | MD (95% CI); p-value |
| δ = 0 (i.e. MAR) |  |  |
| δ = 0.5 |  |  |
| δ = 1.0 |  |  |
| δ = 1.5 |  |  |
| δ = 2.0 |  |  |
| δ = 2.5 |  |  |
| δ = ? (‘tipping point*’) |  |  |

**may be different for each comparison*

We would also explore the possibility that data were only MNAR in the experimental arms by modifying the imputed values only for those allocated to conservative management plus fludrocortisone or midodrine. We would also determine the ‘tipping point’ value for δ in this scenario.

We did not anticipate missing baseline data for the variables included in the primary analysis model as these are either stratification factors and hence required for randomisation or required to confirm eligibility (i.e. the OHQ score). If however, the baseline OHQ score was missing we would use simple imputation and replace the missing value with the mean of all non-missing baseline values [13].

### **REferences**

1. Kaufmann H, Malamut R, Norcliffe-Kaufmann L, Rosa K, Freeman R: **The Orthostatic Hypotension Questionnaire (OHQ): validation of a novel symptom assessment scale**. *Clin Auton Res* 2012, **22**(2):79-90.
2. Frith J, Newton JL: **Validation of a questionnaire for orthostatic hypotension for routine clinical use**. *Geriatr Gerontol Int* 2016, **16**(7):785-790.
3. Nouri FM, Lincoln NB. **An extended activities of daily living scale for stroke patients.** Clinical Rehabilitation. 1987;**1**:301-5.
4. Bell ML, Fairclough DL. **Practical and statistical issues in missing data for longitudinal patient-reported outcomes.** *Statistical Methods in Medical Research* 2014, **23**(5):440-459.
5. Rodgers H, Howel D, Bhattarai N, Cant R, Drummond A, Ford GA, et al. **Evaluation of an Extended Stroke Rehabilitation Service (EXTRAS): A Randomized Controlled Trial and Economic Analysis.** *Stroke* 2019, **50**(12):3561-3568.
6. Spice CL, Morotti W, George S, Dent THS, Rose J, Harris S, Gordon CJ: **The Winchester falls project: a randomised controlled trial of secondary prevention of falls in older people**. *Age & Ageing* 2009, **38**(1):33-40.
7. Ciaschini PM, Straus SE, Dolovich LR, Goeree RA, Leung KM, Woods CR, Zimmerman GM, Majumdar SR, Spadafora S, Fera LA *et al*: **Community-based intervention to optimise falls risk management: a randomised controlled trial**. *Age Ageing* 2009, **38**(6):724-730.
8. Tomlinson CL, Stowe R, Patel S, Rick C, Gray R, Clarke CE: **Systematic Review of Levodopa Dose Equivalency Reporting in Parkinson’s Disease.** *Movement Disorders* 2010, **25**(15): 2649-2684.
9. **ICH E9 (R1) addendum on estimands and sensitivity analysis in clinical trials to the guideline on statistical principles for clinical trials.** EMA/CHMP/ICH/436221/2017.
10. Cro S, Morris TP, Kenward MG, Carpenter JR: **Sensitivity analysis for clinical trials with missing continuous outcome data using controlled multiple imputaion: A practical guide.** *Statistics in Medicine* 2020; **39**:2815-2842.
11. White IR, Royston P, Wood AM: **Multiple imputation using chained equations: issues and guidance for practice.** *Statistics in Medicine* 2011;**30**(4):377-399.
12. Rubin, DB: **Multiple Imputation for Nonreponse in Surveys.** *Wiley Series in Probability and Mathematical Statistics.* New York, NY: John Wiley & Sons; 1987.
13. White IR, Thompson SG: **Adjusting for partially missing** baseline **measurements in randomized trials.** *Statistics in Medicine* 2005;**24**(7):993-1007.

Document 2. Health Economics Analysis Plan

Author Tumi Sotire Health Economist

Reviewed by: Professor Luke Vale	Health Economist 30/07/2023

Outline of economic analysis

The key objective of this economic plan is to outline how an economic evaluation will be carried out as part of the CONFORM-OH trial. It was planned that the study will include a within-trial cost-utility analysis with a longer-term model-based extrapolation to estimate the incremental cost per quality-adjusted life-years (QALYs). QALYs will be derived based on responses to the EQ-5D-5L. The rationale behind using a model-based analysis and a trial-based analysis is that the results of the within-trial study might indicate that a treatment strategy for Orthostatic Hypotension is not cost-effective. Still, it is plausible that extrapolation might change this conclusion. However, the converse can also occur, where a strategy initially found to be cost-effective becomes cost-ineffective when longer-term costs and effects are considered. Therefore, an economic evaluation model was planned to extrapolate cost and QALYs beyond the 12-month trial follow-up period. Section 4 presents further details of this analysis.

The perspective of the CONFORM trial is that of the UK National Health Service (NHS) and Personal and Social Services (PSS). Healthcare resource use will be the main cost of this economic evaluation. The economic assessment will calculate the average cost to the NHS of interventions, conservative management plus Fludrocortisone and conservative management plus Midodrine. We will also calculate the average costs of the comparator, conservative management. A sensitivity analysis will explore a broader perspective considering the price falling on individuals or their families/carers. Within the sensitivity analysis, this cost will include direct, e.g., out-of-pocket costs of any medications or therapies purchased and indirect costs such as time off, paid work and travel, and costs to the NHS.

The within-trial economic evaluation was planned to report the following economic outcomes:

- NHS cost of managing individuals with Orthostatic Hypotension over 12 months.
- Direct and indirect costs fall on individuals who suffer from Orthostatic Hypotension and their families/carers over 12 months.
- QALYs estimated based on responses to the EQ-5D-5L administered at baseline, three, six-, and 12-months post-randomisation.

The incremental cost-effectiveness ratio (ICER) is the incremental cost per QALY gained for a more effective but more costly intervention than a less costly but less effective one.

- The model-based economic evaluation was planned to report the following economic outcomes:
- NHS cost of managing individuals with Orthostatic Hypotension over the estimated patient lifetime.
- Direct and indirect costs fall on individuals who suffer from O and their families/carers over the estimated patient lifetime.
- QALYs estimated based on responses to the EQ-5D-5L over the estimated patient lifetime.

The incremental cost-effectiveness ratio (ICER) is the incremental cost per QALY gained for a more effective but more costly intervention than a less costly but less effective on

CONFORM-OH has closed early as recruitment did not reach planned targets. This health economics analysis plan reports the originally planned Within-Trial Analysis (section 3); the plan for the economic analysis of the data given the early termination is reported in section 4.

Within – Trial analysis

Using the cost and effectiveness data derived from the trial allows the estimation of Fludrocortisone's cost-effectiveness compared to Midodrine and Conservative Management. The economic analysis will be an intention-to-treat (ITT) analysis.

Description of management strategies

The CONFORM protocol version 2.0 provides a more detailed description of the OH strategies. The CONFORM study is analysing the following management strategies.

- Conservative management - In this study, non-pharmacologic therapy is standard first-line care and represents the control arm of the trial.
- Fludrocortisone with conservative management -The typical starting dose will be 100 micrograms, and the highest amount used in this study will be 400 micrograms. The PI has judged participants to be frailer and may have a lower starting dose of 50 micrograms.
- Midodrine with conservative management - The recommended dose range is 2.5 milligrams three to 10 milligrams three times a day. Within this study, lower frequencies and doses may also be accepted.

The duration of follow-up for the three arms is 12 months from randomisation.

Format of the incremental analysis

Table 1 will present all analysis costs, QALYs, and the ICER. Treatments A-C do not a priori represent any of the interventions. Instead, the treatments are ordered from least costly (treatment A) to most costly (treatment C). The table assumes that treatment B is more costly and more effective than treatment A and that treatment C is more costly and more effective than treatment B. Thus, the calculation of ICERs compares treatment B to treatment A and treatment C. Should one or two treatments be more costly and no more effective (and possibly less effective), these treatments will be considered dominated and dropped from the analysis. For example, suppose treatment B is more costly and no more effective than treatment A. I. In this case, treatment A will dominate B. In this situation, the ICER would be calculated for treatment C compared with treatment A (assuming treatment C is more effective than treatment A).

| *Treatment Arm* | *Costs* | *Incremental Costs* | *Effects* | *Incremental Effects* | *ICER* |
| --- | --- | --- | --- | --- | --- |
| *A* | *A* |  |  |  |  |
| *B* | *B* | *∆ costs (B-A)* | *B* | *∆ effects (B-A)* | *∆ costs/∆ effects for treatment B vs treatment A* |
| *C* | *C* | *∆ costs (C-B)* | *C* | *∆ effects (C-B)* | *∆ costs/∆ effects*  *for treatment C vs treatment B* |

Table1:	Incremental cost-effectiveness of conservative management vs Fludrocortisone with conservative management vs Midodrine with conservative management

Estimation of costs

Costs and frequency of use of services and costs

When participants consent to enter the study, the researchers randomly allocate them into one of the three trial arms. Each of the three arms is associated with a different cost. For the within-trial analysis, we will base these costs on the interventions and healthcare services used during the 12-month follow-up period post-randomisation.

Costs of Interventions

Three management strategies are being evaluated as part of this study are:

- Conservative management only (non-pharmacologic treatments)
- Conservative management plus Fludrocortisone
- Conservative management plus Midodrine

Conservative management costs

We will base each form of conservative management on the data collected within the electronic case report form (e-CRF) and estimate it on an individual-participant level (micro-costing). The costs of conservative management will be a sum of every aspect, considering the mix of staffing overheads, disposable equipment, and reusable equipment.

The estimation of these costs will utilise the dummy tables presented in the Appendix, specifically Table 5.1.

We will record the length of time and the number of intervals for the conservative management interventions. The staff costs will consist of the grade of the health care professional per hour. These costs have arisen because health professionals' numbers, types, and grades will be assumed based on consultations with the research team. Total staff costs will be calculated by multiplying the cost per session by the number of sessions within a particular time horizon. The costs per session would be derived by multiplying the duration of a session in minutes by the cost of staff per minute. The cost of staff per minute will be gathered from the Unit costs of health and social care (15). Total staff costs will allow for the average expenses per patient per arm. The total overhead costs will be calculated by multiplying the costs per session by the number of sessions. The costs of overheads will be retrieved from the Unit costs of health and social care (15).

The research team will need to confirm what reusable and consumable equipment is required for each type of conservative management. First, the cost per session will be multiplied by the number of sessions to calculate the total disposable equipment costs. Next, the disposable equipment used will be multiplied by the cost per item to calculate the total equipment costs. The cost per item would be sourced from the NHS Supply Chain website (16). Finally, the number of reusable equipment used in a session will be multiplied by the cost per item, which will then be multiplied by the number of times equipment can be used, the life span in years, and the discount rate. This multiplication will be multiplied by the number of sessions to calculate total reusable costs.

NHS tariffs for conservative intervention will be used in sensitivity analysis, and the results compared to the micro-setting will be estimated using the methods described above.

Cost of Fludrocortisone

As described above, individuals will receive a starting dose of 10m micrograms daily; frailer patients may receive a starting dose of 50 micrograms. This decision will be based on the discretion of the clinician. Clinicians will be encouraged to titrate towards the highest clinically effective dose. As suggested by the British National Formulary (BNF), the highest dose used within this trial is 400 micrograms; however, most sites will use the maximum dose of 300 micrograms as this is the usual practice. The dummy tables used to estimate these costs are shown in the Appendix, Table 5.2.

The unit costs of the Fludrocortisone will be derived from the BNF (17). The BNF documents the costs per pack of Fludrocortisone and the number of tablets in a pack allowing for the cost per tablet to be calculated by dividing the number of tablets by the cost per pack. The number of tablets used per dose will be calculated by dividing the dose from the protocol by the mass of the tablet. The cost per tablet will be multiplied by the number of tablets used per dose to calculate the total daily costs. Total costs will be calculated by multiplying the total cost per day by the time horizon in days.

- cost per tablet = cost per pack/number of tablets in a pack
- number of tablets used per dose = dose from protocol/ mass of tablets
- total costs per day = cost per tablet X number of tablets used per dose
- total costs = total costs per day X by time horizon in days

In addition, the costs of the clinicians administering the therapy can be identified in the Unit costs of health and social care and NHS tariffs (15, 18). The staff costs for administering the medication will be getting the grade of the staff, which the research team will confirm. The hourly rate of this staff will be retrieved from the Unit cost of health and social care. The research team would estimate the mean time required to administer Fludrocortisone. Staff costs per session will be calculated by dividing the hourly staff rate by the time needed to help with the medication. Total staff costs will be calculated by multiplying staff costs per session by the number of sessions.

The estimates of the unit costs of Fludrocortisone will be added to the staff costs of administering. Total costs will be calculated for each participant, and an average cost per arm will be summarised as the average total resource use and total intervention cost per arm.

Cost of Midodrine

Participants who are randomised to Midodrine with conservative management will typically receive a starting dose of 2.5mmg TDS orally. Depending on the discretion of the clinician, lower doses and frequencies may be accepted. The maximum dose in this study is 10 mg. The costs of Midodrine will be estimated by the same method used for Fludrocortisone explained in the paragraph above, and the unit cost can be found in the BNF (19). The dummy tables used to estimate these costs are shown in the Appendix, Table 5.2.

*Costs of NHS Resources*

During the following-up period, participants may utilise additional healthcare resources. Examples of Primary care resources can include GP, nurse/or other healthcare professional consultations. Primary care consultations can occur in various places, healthcare providers' various places, healthcare ’providers' various places, healthcare providers' practices, the participant's home, and over the telephone (including phone calls to NHS call centres). Secondary care resource use can refer to A and E departments, outpatient visits, and hospital admissions, which could be a day or overnight.

The Health Utilisation Questionnaires are used to capture the use of primary care services, including the number of contacts with GP, nurses, and " other health professionals" "". We aim to capture all potential forms of primary care costs were captured. For example, primary care consultations may be face-to-face at the practice, at the '" 'participant's home, or by telephone. The differentiation is important as the cost of each of these varies (see below on derivation of unit costs). Secondary care refers to visits to an A & E department, outpatient clinics and hospital admissions as a day patient or as an inpatient (inpatient admissions are defined as where the person stays at least one night in the hospital).

Healthcare resource use is associated with different unit costs, regardless of whether these resources are primary or secondary. Details of resource use of primary and secondary care will be captured using the e-CRF and follow-up Health Utilisation Questionnaires at baseline, three months, six months, and 12 months after the trial. The dummy tables used to estimate these costs are shown in the Appendix, Table 5.3.

The number of face-to-face or phone appointments with their GP or nurse within a specific time horizon will be extracted from the e-CRF. The cost per minute and the mean length of consultation in minutes can be removed from the Unit cost of health and Social Care (15). The costs per consultation will be multiplied by the number of appointments within a time horizon to calculate the total staff costs per patient for each patient, thus allowing us to calculate the average staff per patient for each intervention arm. The dummy tables used to estimate these costs are shown in the Appendix, Table 5.3.

The ambulance service unit cost can be extracted from the NHS cost Schedule. The NHS cost Schedule lists four different types of ambulance services (18). The average unit costs will be calculated, and the average costs will be multiplied by the number of visits within a particular time horizon to calculate the total cost of ambulance services per patient. The average cost per patient would then be calculated for each other three intervention arms. The dummy tables used to estimate these costs are shown in the Appendix, Table 5.3.

An average of the unit costs of the Accident & Emergency currency descriptions with the highest number of data submissions (any currency description with a number of data submissions higher than 110 will be included in the average)(18). Accident and emergency services unit costs will be extracted from the NHS cost Schedule. The total cost will be calculated by multiplying the unit cost by the number of appointments documented in the e-CRF. The average Accident and emergency services cost will then be calculated for each of the three intervention arms. The dummy tables used to estimate these costs are shown in the Appendix, Table 5.3.

Cost of Private Services (non-NHS) Care

The cost of private services and the number of particular visits will be extracted from the e-CRF. Private services include things such as physiotherapy and acupuncture. First, the total cost for each private patient service will be calculated by multiplying the number of private service visits by the amount each patient paid for the private service they used. Then, the private costs will be grouped with the same theme. Finally, the average costs of private services for each of the three arms will be calculated. The dummy tables used to estimate these costs are shown in the Appendix, Table 5.3.

Cost of Personal Social Services

The e-CRF will allow us to determine what social benefit each participant receives. These benefits include Attendance Allowance, Pension Savings Credit, and Pensions Guarantee Credit; the cost of these benefits can be found on the the.Gov.UK website (20, 21). These costs will be multiplied by the length of the time horizon to calculate the total cost per participant. We will then be able to calculate the average cost of benefits per intervention. A sensitivity analysis would be used to see how the cost-effectiveness would vary if we used the lowest rate compared to the highest. The dummy tables used to estimate these costs are shown in the Appendix, Table 5.3.

The number of social service consultations a participant uses within a given time horizon can be derived from the e-CRF. The Unit Cost of Health and Social Care reports the cost of social care per hour (15). The research team would confirm the meantime one can expect for a social service consultation. Collecting data on the mean length of time will allow us to calculate the average cost of social service intervention. The average cost of social care can be multiplied by the number of social care appointments to calculate the total cost of social care. The total social costs would allow us to calculate the average cost of social per treatment arm.

Time and Travel Questionnaire

The time and travel questionnaire will assist in gathering information on the cost of accessing healthcare resources relating to Orthostatic Hypotension. The questionnaire captures information about the amount of time and money the participant and a 'participant's family member or carer have spent using health care services six months after. The time and travel questionnaire are split into three sections with 12 questions.

The three sections are:

- Hospital outpatient appointments
- Hospital admissions
- GP / Nurse/Walk-in Centre visits

For each of the three sections on the questionnaire, the cost of travel and the opportunity cost in monetary terms for each study participant and the people accompanying them to the appointments will be summated, and an average will be calculated for each treatment arm. In addition, the average cost and opportunity cost will be calculated per participant and the person accompanying the appointment. The dummy tables used to estimate these costs are shown in the Appendix, Table 5.4.

Hospital Outpatients Appointments

This set of questions referred to hospital appointments. Participants who did not have a hospital appointment were told to proceed to the next section. The questionnaire captured information about the mode of transport a participant used to go to a hospital appointment to their most recent. The cost of travel to this appointment for those who used public travel will be extracted from the questionnaire; the time spent travelling to this appointment and the length of time spent at this appointment will also be collected.

The travel costs of those who arrived and used ambulance services to attend the appointment will be derived from the average unit cost of ambulance services from the NHS reference list (18). The distance travelled to the appointment was also recorded for the participants that travelled by car, walked, or cycled. The cost per mile will be extracted from the advisory fuel rates.Gov.uk website (22). The price per mile will be multiplied by the distance travelled to the appointment to calculate the total cost travelled by car. For those that walked and cycled, the cost of travel was not recorded. Instead, the average trip price, the average time spent travelling to an appointment, and the average time spent during an appointment per participant will be calculated for all modes of transport.

Estimates of the opportunity costs of the appointment will be calculated from the questionnaire. Participants will be asked about their main activity if they were not attending the appointment. The national median hourly wage will be calculated using the Office of National Statistics data for the participants who would have been in paid work. This hourly rate will be multiplied by the time spent travelling to and attending the appointment to calculate the opportunity cost of paid work for each employed participant. The opportunity costs of being a homemaker will be calculated by multiplying the national minimum wage, which can be found on the Gov. UK website, by the time, spent travelling to and attending the appointment (23).

A similar process will be used to calculate the opportunity cost for that hourly rate for childcare will be collected from the Coram Childcare Survey 2022; this will be multiplied by the time spent travelling to and attending the appointment (24). The opportunity costs for caring for friends and family would be calculated using the same method mentioned above using the hourly rate of care derived from the UK Care Guide (25). The opportunity costs of those who are retired will be calculated similarly using the state pensions, which can be found on the UK government website (26). The student finance website will be used to provide data needed to calculate the opportunity cost of full-time education. (27).

Similar methods will be used to calculate the opportunity costs of unemployed people by deriving the average monthly allowance from universal credit (28). The standard monthly budget for universal income varies depending on age and marital status. For this trial, we will assume participants are single and older than 25. The opportunity costs for volunteering would be calculated using information from the Office for National Statistics (29). The Office for National Statistics will provide data on average expenditure on recreation and culture, which will be used to calculate the opportunity costs of leisure activities.

A similar process was conducted to calculate the opportunity costs of the family member, friend or carer accompanying the study.

Adverse Events

Information on adverse events, AEs, and serious adverse events (SAE) will be collected via the electronic e-CRF at any time during the trial. AEs may require additional medications or hospitalisation. The cost of the AE will be estimated by calculating the cost of the action taken to resolve the AE. The three possible actions that could be taken are reducing the dose and withdrawing the Investigational Medical Product. IMP or no action. The intervention cost will be calculated as described above until the resolution is conducted for participants who had a dose reduction or no longer received their IMP; as a result, AE Reducing the dose would alter the inputs needed to calculate the intervention costs from the resolution date. The resolution date allows us to calculate the cost of AE over a specific period. A withdrawal of the IMP at a given resolution date will result in only the cost-conservative management calculated from the resolution date. For participants that did not require any action due to the AE, the intervention cost was calculated similarly to the overhead cost of the intervention section. The total cost of adverse events for each intervention will be calculated, thus allowing us to calculate the average prices.

Estimation of total costs

The mean total cost of each intervention group will be calculated by summing the costs of interventions, NHS resources, Private Services, Personal Social Services, Time and Travel and Adverse Events for each participant and then dividing by the number of participants included in the analysis are added together. A sensitivity analysis will be conducted for each cost input to assess the effect of high and low estimates of costs.

Effectiveness measures

The effectiveness measure for the within-trial economic evaluation will be QALYs estimated from responses to the EQ-5D-5L data. The EQ-5D-5L measures health-related quality of life (HRQoL), which the National Institute of Health and Care Excellence (NICE) recommends. This general health questionnaire considers five domains, mobility, self-care, usual activities, pain/discomfort, and anxiety/ depression (31). Each of the five domains has five possible responses; there are, therefore, 3125 possible health states. These responses vary in severity from no, slight, moderate, severe, or extreme problems/inability to conduct usual activities). EQ-5D-5L values are converted into health state utility for each patient at each time point, estimated from a representative sample of the UK population (32). This process is followed by mapping the EQ-5D-5L descriptive data set onto the EQ-5D-3L value set described in the NICE guidelines using the mapping/crosswalk algorithm(31). Finally, we will conduct a sensitivity analysis to investigate the impact of using alternative crosswalk algorithms on the ICER results.

A proxy version of the EQ-5D-5L will be included for participants who cannot self-complete the questionnaires. EQ-5D–5L data will be collected at baseline, three, six and twelve months following randomisation. With the proxy version, the caregiver is asked to rate how they think the patient would rate their health-related quality of life if they could communicate. These responses will be converted into scores using population tariffs (31). The values from both participants and proxy values will be compared when both values are available to calculate QALY simultaneously. Following this, the variance between participant and proxy will be estimated. Where participant values are unavailable in the analysis, proxy values will be used.

The outcome data will then be converted into (QALYs) for each participant using the under-the-curve approach for baseline, three, six and twelve months. A QALY combines the quality of life and life expectancy into a single index (31). Standard deviations, standard errors and confidence intervals will be calculated using standard formulas for each of the mean utility measures. QALYs will be calculated by summing the area under the curve (AUC), connecting the mean utility measures at each time point for all three interventions. The AUC is calculated by summing up the areas of the shapes obtained from the linear relationship between utility scores during the study period (32,33).

Baseline means utility values may be imbalanced between treatment arms. Imbalanced utilities need to be considered when the mean differential QALYs are estimated. Estimation for mean differential QALYS can be achieved using multiple regression methods(34). This method controls for baseline utility values simultaneously, allowing for the Estimation of differential QALYs and the prediction of adjusted QALYS. The regression analysis provides unbiased estimates of differential QALYs between the trial's three arms, increasing the treatment effect's precision (35).

Cost-utility analysis

The cost-utility analysis will base on the incremental cost per QALY gained and will be presented as the incremental cost-effectiveness ratio (ICER) and can be calculated by dividing the change of the expenses by the change in effect (QALYs). The total cost and QALY will be estimated for each arm, which will then be expressed as point estimates of the mean incremental costs and effects (QALYs) and the incremental cost per QALY gained. The UK’s willingness to pay is approximately £20,000 per additional QALY; thus, if conservative management plus Fludrocortisone and conservative management plus Midodrine is not dominant, the ICER is within this threshold, the interventions could be deemed cost-effective by decision-makers. A dominant intervention is an intervention with a lower cost and better health outcomes than the comparator.

Adjusted analysis – seemingly unrelated regression (SUR)

Both unadjusted and adjusted analyses will be performed to estimate the cost-effectiveness of conservative management plus Fludrocortisone compared to conservative management plus Midodrine compared to conservative management alone. The mean incremental costs, effects and cost-effectiveness results will be presented as point estimates.

An adjusted analysis will be carried out using seemingly unrelated regression (SUR) for all comments to estimate the costs and effects of the three interventions and their cost-effectiveness. SUR allows one to simultaneously estimate the costs and effects for each individual, accounting for unobservable individual characteristics to affect both costs and effects, resulting in a potential correlation between two variables. Furthermore, the SUR permits the control for additional covariates such as age, baseline, severity and utility that could potentially affect costs and effects when appropriate. The covariates used in the SUR model will replicate the covariates used in the statistical analysis outlined in the Statistical Analysis Plan. We will analyse Stata using the seemingly unrelated regression command sureg (StataCorp LLC, Texas, USA).

Sensitivity analysis

A stochastic/probabilistic and deterministic sensitivity analysis will be conducted to characterise the uncertainty.

Stochastic sensitivity analysis

A stochastic sensitivity analysis will be conducted. Bootstrapping techniques will be utilised in this analysis (35). (Bootstrapping can be described as the non-parametric technique used to estimate the distributions of important statistics relevant to the model, such as the ICER.(36). Stochastic analysis would explore the potential effects of statistical imprecision of parameters used within the model on costs effects and cost-effectiveness estimates if the sampling process could be repeated many times. Random values will be selected from the cost and QALY data retrieved from the trial. When a random value is used for the bootstrap resample, it is returned to the original sample. Consequently. A bootstrapped dataset derived from the complete case data on costs and outcomes from each trial arm.

This process will be carried out many times (e.g., 1000, 5000, 10,000), creating a sample of bootstrapped mean costs and QALYs with their corresponding distributions. The bootstrapped distribution will be calculated using bootstrapped costs, QALYS, and other parametric statistics. The results from the bootstrapping will be presented as a cost-effectiveness plane to demonstrate the distribution of incremental costs and incremental effects, allowing us to identify the distribution of total incremental costs and incremental effects. The horizontal axis will represent the QALYs difference between the two interventions, and the vertical axis will represent the corresponding cost difference. The bootstrapped results will also be presented as cost-effectiveness acceptability curves (CEACs). CEACs enable us to identify the management strategy that maximises net benefits at each willingness to pay value for additional QALYs gained) (37).

Deterministic sensitivity analysis

Deterministic sensitivity analysis will be used to assess the effect of assumptions such as dose of medications, method of conservative management and unit costs on the results using best-case and worst-case scenario analysis. A societal perspective will be used to explore the impact of patient costs on cost-effectiveness results. The results from the deterministic analysis will determine how the analysis will be performed, including one-way, two-way or multiway analysis.

To identify the primary inputs that could be changed to make Fludrocortisone and Conservative Management more or less cost-effective relative to both Midodrine and Conservative Management and Conservative Management alone.

Intervention costs

Conservative management

As mentioned before, we will use micro-costing to calculate conservative cost management. We will use sensitivity analysis to explore how low and high estimates affect the ICER. In addition, we will also investigate how the type of conservative management can influence the ICER separately. This form of analysis will be achieved by one-way sensitivity analysis. We will use higher and lower values of the intervention cost using a 95% confidence interval. Multiway sensitivity analysis will allow us to assess the impact of variation in the costs of Fludrocortisone and Midodrine.

Fludrocortisone and Midodrine

As mentioned, certain assumptions about the pharmacological interventions, such as the dose strength and the number of tablets used in a day, will be tested using sensitivity analysis. In the sensitivity analysis, we will explore the effect of each medication’s high and low estimations on the ICER.

Cost of services

As mentioned previously, the assumptions about the cost of services would be assessed using sensitivity analysis. This analysis will determine non-NHS, private healthcare, and social care resources. For each cost, we will investigate the effect of using higher and lower cost estimates on the cost-effectiveness results, and whether the ICER goes above or below the threshold. Results from the sensitivity analysis will be presented using a tornado diagram for multiple one-way sensitivity analyses for higher and lower values of service cost.

Utility values

We will use EQ-5D-5L crosswalked to the EQ-5D-3L value set in the base case analysis (31). In sensitivity analyses, we will use alternative crosswalk algorithms (e.g., provided by van Hout et al.) and the population value set for the EQ-5D-5L to investigate the effect of these utility values on the ICER (38).

In a further sensitivity analysis, proxy values based on the degree of discordance between participants and the proxy values will be used in the sensitivity analysis. The reported proxy values will be replaced with alternative values estimated by converting the upper and lower confidence intervals of a paired t-test. This will compare the patient and proxy values into a % variation from the man score by applying that % variation of the observed score. This will mean that high and low values utilised in the sensitivity analysis will be a proxy score * (1+ x) or proxy score * (1-x), when x is the % variation.

Total costs and utility

To deal with missing costs and utility data, we will do multiple imputations in the sensitivity analysis. An ICER will be presented in addition to the base case analysis.

Handling Missing Data

Potential issues may arise when dealing with trial data, such as missing data and not adhering to the protocol. Therefore, a missing data analysis will be carried out.

This missing data analysis will primarily focus on missing data because the e-CRF and participant questionnaires are incomplete. As with most trial-based data collection, not all participants who enter the trial will complete all questionnaires and questions in each questionnaire. Therefore, missing data is expected. When we have a full trial dataset available, the imputation methods will be determined as the most appropriate, depending on the nature of the missing data.

Close Down Plan

Summary of Existing Economic Component

The health economics component for CONFORM-OH,-, had it been recruited to target, would have included a within-trial cost-utility analysis with longer-term model-based extrapolation. For both the within-trial and model-based analysis, the results will be reported as incremental costs, QALYs, and incremental costs per QALY gained.

The within-trial analysis cost and outcomes for the three trial intervention arms would have been based on data collected over the 12-month follow-up period. Costs were to fall on the NHS, personal social services (PSS), participants and their families. These costs were to be based upon the use of health and social services and, as part of the costs falling on patients and their families, the time and travel costs of accessing services. QALYs were to be based upon responses to the EQ-5D-5L collected from participants at baseline, three, six and 12 months.

The plan was also to use an economic model to extrapolate outcomes into the longer term. Outcomes of the model will be expressed in terms of costs to the NHS and PSS, QALYs, and incremental cost per QALY gained. The model will be structured as when there is a choice about whether or not to start medical treatment and which medical treatment to start, and it would follow individuals for the remainder of their life. The model was to be developed per the NICE reference case (398).

Close-down Plan

Given the likely data available from the trial, we no longer propose to conduct a complete within-trial economic evaluation of the model-based analysis. As an alternative, we will use the available trial data to:

1) Provide descriptive statistics (e.g. mean, medians and appropriate measures of variance around completion rates for each health economic data collection tool (i.e. the health service utilisation questionnaire and EQ-5D-5L completed at baseline, 3, 6, 12 months)

2) Provide descriptive statistics (e.g. mean, medians and appropriate measures of variance) for each area of resource use collected at each time point and overall as part of the trial and by group.

3) Provide descriptive statistics for the EQ-5D-5L utility score and EQ-5D-VAS (e.g. mean, medians and appropriate measures of variance) at each time point and overall as part of the trial overall and by group.

4) For each trial respondent, map changes by level on the EQ-5D-5L descriptive framework for responses to the EQ-5D-5L at baseline, 3 and 6 months. There will be insufficient data at 12 months to make a similar work useful.

Analyses 1-3 are consistent with the work proposed in Section 3 of the Health Economic Analysis Plan, Version 0.2. These analyses will provide information for any subsequent evidence synthesis project (e.g., a systematic review or modelling exercise). The data will also help plan future evaluations addressing the same or similar research questions. Specifically, question 1 will provide some information as to whether the tools used can be completed by this group of people (or their proxies). Question 2 will illustrate where resources are used, which may help design future data collection tools that balance data collection against respondent burden. Questions 3 and 4 will help inform whether the EQ-5D-5L can detect any changes in health status in this group of people.

In addition to the above, we propose using the data from the trial assembled from the literature and expert opinion to conduct an early economic evaluation model. This approach is used in other healthcare contexts to help design future primary research. This element aims to identify key uncertainties and estimate, using the methods of Value of Sample Information, the sample size required for a definitive trial to address the research question. Expected value of information will be calculated.

References

1. Freeman R, Wieling W, Axelrod FB, Benditt DG, Benarroch E, Biaggioni I, Cheshire WP, Chelimsky T, Cortelli P, Gibbons CH et al.: Consensus statement on the definition of orthostatic Hypotension, neurally mediated syncope and the postural tachycardia syndrome. Clinical Autonomic Research 2011, 21(2):69-72.
2. Zhou Y, Ke SJ, Qiu XP, Liu LB: Prevalence, risk factors, and prognosis of orthostatic Hypotension in diabetic patients: A systematic review and meta-analysis. Medicine (Baltimore) 2017, 96(36):e8004.
3. Velseboer DC, de Haan RJ, Wieling W, Goldstein DS, de Bie RM: Prevalence of orthostatic hypotension in Parkinson's disease: a systematic review and meta-analysis. Parkinsonism & Related Disorders 2011, 17(10):724-729.
4. Kaufmann H, Malamut R, Norcliffe-Kaufmann L, Rosa K, Freeman R: The Orthostatic Hypotension Questionnaire (OHQ): validation of a novel symptom assessment scale. Clin Auton Res 2012, 22(2):79-90.
5. Angelousi A, Girerd N, Benetos A, Frimat L, Gautier S, Weryha G, Boivin JM: Association between orthostatic Hypotension and cardiovascular risk, cerebrovascular risk, cognitive decline and falls as well as overall mortality: a systematic review and meta-analysis. J Hypertens 2014, 32(8):1562-1571.
6. Ricci F, Fedorowski A, Radico F, Romanello M, Tatasciore A, Di Nicola M, Zimarino M, De Caterina R: Cardiovascular morbidity and mortality related to orthostatic Hypotension: a meta-analysis of prospective observational studies. Eur Heart J 2015, 36(25):1609-1617.
7. McDonald C, Pearce M, Kerr SR, Newton J: A prospective study of the association between orthostatic hypotension and falls: definition matters. Age Ageing 2016.
8. Frith J, Elliott CS, Bashir A, Newton Jl: Public and patient research priorities for orthostatic hypotension. Age and Ageing 2014, 43(6):865-868.
9. Frith J, Parry SW: New Horizons in orthostatic hypotension. Age Ageing 2017, 46(2):168-174.
10. Brignole M, Moya A, de Lange FJ, Deharo J-C, Elliott PM, Fanciulli A, Fedorowski A, Furlan R, Kenny RA, Martín A et al: 2018 ESC Guidelines for the diagnosis and management of syncope. European Heart Journal 2018, 39(21):1883-1948
11. Logan IC, Witham MD: Efficacy of treatments for orthostatic hypotension: a systematic review. Age and Ageing 2012, 41(5):587-594
12. Mills PB, Fung CK, Travlos A, Krassioukov A: Nonpharmacologic management of orthostatic hypotension: a systematic review. Arch Phys Med Rehabil 2015, 96(2):366-375 e366.
13. Ong AC, Myint PK, Shepstone L, Potter JF: A systematic review of the pharmacological management of orthostatic hypotension. Int J Clin Pract 2013, 67(7):633-646.
14. Parsaik AK, Singh B, Altayar O, Mascarenhas SS, Singh SK, Erwin PJ, Murad MH: Midodrine for orthostatic hypotension: a systematic review and meta-analysis of clinical trials. J Gen Intern Med 2013, 28(11):1496-1503.
15. Unit Costs of Health and Social Care 2021 | PSSRU [Internet]. Pssru.ac.uk. 2022 [cited 12 July 2022]. Available from: <https://www.pssru.ac.uk/project-pages/unit-costs/unit-costs-of-health-and-social-care-2021/>
16. NHS Supply Chain. 2022 [cited 27 July 2022]. Available from: <https://www.supplychain.nhs.uk/>
17. Medicinal forms | Fludrocortisone acetate | Drugs | BNF content published by NICE [Internet]. Bnf.nice.org.uk. 2022 [cited 14 July 2022]. Available from: <https://bnf.nice.org.uk/drugs/fludrocortisone-acetate/medicinal-forms/>
18. England N. NHS England » National Cost Collection for the NHS [Internet]. England.nhs.uk. 2022 [cited 14 July 2022]. Available from: <https://www.england.nhs.uk/costing-in-the-nhs/national-cost-collection/>
19. Medicinal forms | Midodrine hydrochloride | Drugs | BNF content published by NICE [Internet]. Bnf.nice.org.uk. 2022 [cited 13 July 2022]. Available from: <https://bnf.nice.org.uk/drugs/midodrine-hydrochloride/medicinal-forms/>
20. Attendance Allowance [Internet]. GOV.UK. 2022 [cited 19 July 2022]. Available from: <https://www.gov.uk/attendance-allowance/what-youll-get>
21. Pension Credit [Internet]. GOV.UK. 2022 [cited 27 July 2022]. Available from: <https://www.gov.uk/pension-credit/what-youll-get>
22. Advisory fuel rates [Internet]. GOV.UK. 2022 [cited 28 July 2022]. Available from: <https://www.gov.uk/guidance/advisory-fuel-rates>
23. The National Minimum Wage in 2022. GOV.UK. 2022 [cited 1 August 2022]. Available from: <https://www.gov.uk/government/publications/the-national-minimum-wage-in-2022>
24. Familyandchildcaretrust.org. 2022 [cited 27 July 2022]. Available from: <https://www.familyandchildcaretrust.org/sites/default/files/Resource%20Library/Final%20Version%20Coram%20Childcare%20Survey%202022_0.pdf>
25. HOW MUCH DOES IN HOME CARE COST 2022? A definitive guide [Internet]. UK Care Guide. 2022 [cited 3 August 2022]. Available from: <https://ukcareguide.co.uk/home-care-costs/>
26. The new State Pension [Internet]. GOV.UK. 2022 [cited 2 August 2022]. Available from: <https://www.gov.uk/new-state-pension/what-youll-get>
27. Allingham T. This is how much uni REALLY costs you [Internet]. Save the Student. 2022 [cited 2 August 2022]. Available from: <https://www.savethestudent.org/student-finance/university-study-cost.html>
28. Universal Credit [Internet]. GOV.UK. 2022 [cited 2 August 2022]. Available from: <https://www.gov.uk/universal-credit/what-youll-get>
29. [Internet]. 2022 [cited 2 August 2022]. Available from: <https://www.ons.gov.uk/employmentandlabourmarket/peopleinwork/earningsandworkinghours/articles/billionpoundlossinvolunteeringeffort/>
30. Family spending workbook 1: detailed expenditure and trends - Office for National Statistics [Internet]. Ons.gov.uk. 2022 [cited 2 August 2022]. Available from: <https://www.ons.gov.uk/peoplepopulationandcommunity/personalandhouseholdfinances/expenditure/datasets/familyspendingworkbook1detailedexpenditureandtrends>
31. National Institute for Health and Care Excellence (NICE). Guide to the methods of technology appraisal 2013. 2013, National Institute for Health and Care Excellence London, UK
32. Devlin NJ, Shah KK, Feng Y, Mulhern B, Hout B. Valuing health-related quality of life: An EQ-5D-5L value set for England. Health Economics. 2018;27(1):7-22.
33. Manca A, Hawkins N, Sculpher MJ. Estimating mean QALYs in trial‐based cost‐effectiveness analysis: the importance of controlling for baseline utility. Health economics. 2005 May;14(5):487-96.
34. Hunter RM, Baio G, Butt T, Morris S, Round J, Freemantle N. An educational review of the statistical issues in analysing utility data for cost-utility analysis. Pharmacoeconomics. 2015 Apr;33(4):355-66.
35. Barber JA & Thompson S.G. Analysis of cost data in randomised trials: an application of the non‐parametric bootstrap. Statistics in Medicine, 2000: 19(23): pp. 3219-3236.
36. Probabilistic/Stochastic Sensitivity Analysis - YHEC - York Health Economics Consortium [Internet]. YHEC - York Health Economics Consortium. 2022 [cited 16 April 2022]. Available from: <https://yhec.co.uk/glossary/probabilistic-stochastic-sensitivity-analysis/>
37. Fenwick E., Claxton K., Sculpher M. Representing uncertainty: the role of cost-effectiveness acceptability curves. Health Econ, 2001: 10(8): pp. 779-8
38. Van Hout B, Janssen MF, Feng YS, Kohlmann T, Busschbach J, Golicki D, Lloyd A, Scalone L, Kind P, Pickard AS. Interim scoring for the EQ-5D-5L: mapping the EQ-5D-5L to EQ-5D-3L value sets. Value in health. 2012 Jul 1;15(5):708-15.
39. National Institute for Health and Care Excellence (NICE). Guide to the Methods of Technology Appraisal 2013. 2013, National Institute for Health and Care Excellence London, UK from the probabilistic sensitivity analysis outputs

Dummy Tables

6.1Conservative Management Costs

6.1.1 Education intervention

6.1.1.1 staff

| Staff | | Time involved | Cost per minute | Cost per consultations | Number of Consultations | Total staff costs* | Notes |
| --- | --- | --- | --- | --- | --- | --- | --- |
| Type | Grade |  |  |  |  |  |  |
| Staff member 1 |  |  |  |  |  |  |  |
| Staff member 2 |  |  |  |  |  |  |  |
| Totals |  |  |  |  |  |  |  |

* These are calculated

6.1.1.2 Overheads

| Overhead type | Unit cost | Cost per session* | Number of sessions* | Total overhead costs* | Note |
| --- | --- | --- | --- | --- | --- |
| Type | Cost per minute |  |  |  |  |
| Room cost |  |  |  |  |  |
| Heat power and light |  |  |  |  |  |
| Internet |  |  |  |  |  |
| Print outs |  |  |  |  |  |
| Totals |  |  |  |  |  |

* These are calculated

6.1.1.3 equipment disposable

| Equipment item | | Cost per item | Cost per session* | Number of sessions | Total Equipment costs* |
| --- | --- | --- | --- | --- | --- |
| Type | Number used |  |  |  |  |
| Equipment 1 |  |  |  |  |  |
| Equipment 2 |  |  |  |  |  |
| Totals |  |  |  |  |  |

* These are calculated

| Equipment item  Type | N0 used | times equipment can be used | Lifespan (years) | Discount rate | Cost per item per use | Cost per session* | Number of sessions* | Total reusable Equipment costs* |
| --- | --- | --- | --- | --- | --- | --- | --- | --- |
| Equipment 1 |  |  |  |  |  |  |  |  |
| Equipment 2 |  |  |  |  |  |  |  |  |
| Totals |  |  |  |  |  |  |  |  |

* These are calculated

6.1.2 Trigger avoidance

6.1.2.1 staff

| Staff | | Time involved | Cost per minute | Cost per Consultations | Number of Consultations | Total staff costs* | Notes |
| --- | --- | --- | --- | --- | --- | --- | --- |
| Type | Grade |  |  |  |  |  |  |
| Staff member 1 |  |  |  |  |  |  |  |
| Staff member 2 |  |  |  |  |  |  |  |
| Totals |  |  |  |  |  |  |  |

6.1.2.2 Overheads

| Overhead type | Cost per minute | Cost per session* | Number of sessions* | Total overhead costs* |
| --- | --- | --- | --- | --- |
| Room cost |  |  |  |  |
| Heat power and light |  |  |  |  |
| Internet |  |  |  |  |
| Print outs |  |  |  |  |
| Totals |  |  |  |  |

* These are calculated

6.1.2.3 equipment disposable

| Equipment item | | Cost per item | Cost per session* | Number of sessions | Total Equipment costs* |
| --- | --- | --- | --- | --- | --- |
| Type | Number used |  |  |  |  |
| Equipment 1 |  |  |  |  |  |
| Equipment 2 |  |  |  |  |  |
| Totals |  |  |  |  |  |

* These are calculated

6.1.2.4 equipment reusable

| Equipment item | Number used | Number of times equipment can be used | Lifespan (years) | Discount rate | Cost per item per use | Cost per session* | Number of sessions* | Total reusable Equipment costs* |
| --- | --- | --- | --- | --- | --- | --- | --- | --- |
| Equipment 1 |  |  |  |  |  |  |  |  |
| Equipment 2 |  |  |  |  |  |  |  |  |
| Totals |  |  |  |  |  |  |  |  |

6.1.3 Safe Standing

6.1.3.1 staff

| Staff | | Time involved | Cost per minute | Cost per Consultations | Number of Consultations | Total staff costs* |
| --- | --- | --- | --- | --- | --- | --- |
| Type | Grade |  |  |  |  |  |
| Staff member 1 |  |  |  |  |  |  |
| Staff member 2 |  |  |  |  |  |  |
| Totals |  |  |  |  |  |  |

* These are calculated

6.1.3.2 Overheads

| Overhead type | Cost per minute | Cost per session* | Number of sessions* | Total overhead costs* | Notes |
| --- | --- | --- | --- | --- | --- |
| Room cost |  |  |  |  |  |
| Heat power and light |  |  |  |  |  |
| Internet |  |  |  |  |  |
| Print outs |  |  |  |  |  |
| Totals |  |  |  |  |  |

* These are calculate

6.1.3.3 equipment disposable

| Equipment item | | Cost per item | Cost per Session* | Number of sessions | Total Equipment costs* |
| --- | --- | --- | --- | --- | --- |
| Type | Number used |  |  |  |  |
| Equipment 1 |  |  |  |  |  |
| Equipment 2 |  |  |  |  |  |
| Totals |  |  |  |  |  |

* These are calculated

6.1.3.4 equipment reusable

| Equipment item | Number used | Number of times equipment can be used | Lifespan (years) | Discount rate | Cost per item per use | Cost per session* | Number of sessions* | Total reusable Equipment costs* |
| --- | --- | --- | --- | --- | --- | --- | --- | --- |
| Equipment 1 |  |  |  |  |  |  |  |  |
| Equipment 2 |  |  |  |  |  |  |  |  |
| Totals |  |  |  |  |  |  |  |  |

* These are calculated

.1.4 Physical counter–manoeuvre

6.1.4.1 staff

| Staff | | Time involved | Cost per minute | Cost per Consultations | Number of Consultations | Total staff costs* |
| --- | --- | --- | --- | --- | --- | --- |
| Type | Grade |  |  |  |  |  |
| Staff member 1 |  |  |  |  |  |  |
| Staff member 2 |  |  |  |  |  |  |
| Totals |  |  |  |  |  |  |

* These are calculated

6.1.4.2 Overheads

| Overhead type | Cost per minute | Cost per session*** | Number of sessions* | Total overhead costs* |
| --- | --- | --- | --- | --- |
| Type |  |  |  |  |
| Room cost |  |  |  |  |
| Heat power and light |  |  |  |  |
| Internet |  |  |  |  |
| Print outs |  |  |  |  |
| Totals |  |  |  |  |

* These are calculated

6.1.4.3 equipment disposable

| Equipment item | | Cost per item | Cost per session* | Number of sessions | Total Equipment costs* |
| --- | --- | --- | --- | --- | --- |
| Type | Number used |  |  |  |  |
| Equipment 1 |  |  |  |  |  |
| Equipment 2 |  |  |  |  |  |
| Totals |  |  |  |  |  |

* These are calculated

6.1.4.4Equipment reusable

| Equipment item | Number used | Number of times equipment can be used | Lifespan (years) | Discount rate | Cost per item per use | Cost per session* | Number of sessions* | Total reusable Equipment costs* |
| --- | --- | --- | --- | --- | --- | --- | --- | --- |
| Type |  |  |  |  |  |  |  |  |
| Equipment 1 |  |  |  |  |  |  |  |  |
| Equipment 2 |  |  |  |  |  |  |  |  |
| Totals |  |  |  |  |  |  |  |  |

* These are calculated

6.1.5 Fluid and salt intake

6.1.5.1Staff

| Staff | | Time involved | Cost per minute | Cost per consultations | Number of Consultations | Total staff costs* |
| --- | --- | --- | --- | --- | --- | --- |
| Type | Grade |  |  |  |  |  |
| Staff member 1 |  |  |  |  |  |  |
| Staff member 2 |  |  |  |  |  |  |
| Totals |  |  |  |  |  |  |

* These are calculated

6.1.5.2 Overheads

| Overhead type | Cost per minute | Cost per session* | Number of sessions* | Total overhead costs* |
| --- | --- | --- | --- | --- |
| Room cost |  |  |  |  |
| Heat power and light |  |  |  |  |
| Internet |  |  |  |  |
| Print outs |  |  |  |  |
| Totals |  |  |  |  |

* These are calculated

6.1.5.3 equipment disposable

| Equipment item | | Cost per item | Cost per session* | Number of sessions | Total Equipment costs* |
| --- | --- | --- | --- | --- | --- |
| Type | Number used |  |  |  |  |
| Equipment 1 |  |  |  |  |  |
| Equipment 2 |  |  |  |  |  |
| Totals |  |  |  |  |  |

* These are calculated

6.1.5.4 equipment reusable

| Equipment item | Number used | Number of times equipment can be used | Lifespan (years) | Discount rate | Cost per item per use | Cost per session* | Number of sessions* | Total reusable Equipment costs* |
| --- | --- | --- | --- | --- | --- | --- | --- | --- |
| Equipment 1 |  |  |  |  |  |  |  |  |
| Equipment 2 |  |  |  |  |  |  |  |  |
| Totals |  |  |  |  |  |  |  |  |

* These are calculated

6.1.6 Compression hosiery

6.1.6.1 staff

| Staff | | Time involved | Cost per minute | Cost per Consultations | Number of Consultations | Total staff costs* |
| --- | --- | --- | --- | --- | --- | --- |
| Type | Grade |  |  |  |  |  |
| Staff member 1 |  |  |  |  |  |  |
| Staff member 2 |  |  |  |  |  |  |
| Totals |  |  |  |  |  |  |

* These are calculated

5.1.6.2 Overheads

| Overhead type | Cost per minute | Cost per session* | Number of sessions* | Total overhead costs* |
| --- | --- | --- | --- | --- |
| Room cost |  |  |  |  |
| Heat power and light |  |  |  |  |
| Internet |  |  |  |  |
| Print outs |  |  |  |  |
| Totals |  |  |  |  |

* These are calculated

6.1.6.3 equipment disposable

| Equipment item | | Cost per item | Cost per session* | Number of sessions | Total Equipment costs* |
| --- | --- | --- | --- | --- | --- |
| Type | Number used |  |  |  |  |
| Equipment 1 |  |  |  |  |  |
| Equipment 2 |  |  |  |  |  |
| Totals |  |  |  |  |  |

* These are calculated

6.1.6.4Equipment reusable

| Equipment item | Number used | Number of times equipment can be used | Lifespan (years) | Discount rate | Cost per item per use | Cost per session* | Number of sessions* | Total reusable Equipment costs* |
| --- | --- | --- | --- | --- | --- | --- | --- | --- |
| Equipment 1 |  |  |  |  |  |  |  |  |
| Equipment 2 |  |  |  |  |  |  |  |  |
| Totals |  |  |  |  |  |  |  |  |

* These are calculated

6.2 Pharmacological interventions

6.2.1 Fludrocortisone

| Staff Administering Fludrocortisone | Grade | Time involved | Cost per minute | Cost per Session | Number of sessions | Total staff costs* |
| --- | --- | --- | --- | --- | --- | --- |
| Staff Member 1 |  |  |  |  |  |  |
| Staff Member 2 |  |  |  |  |  |  |
| Total |  |  |  |  |  |  |

* These are calculated

6.2.2 Cost of medications

| Dose in Protocol | Strength of tablet according to BNF | Cost per pack | Number of tablets per pack | Frequency per day | Cost per tablet £ | Number of tables used per dose | Total cost per day | Time horizon in days | Total Costs |
| --- | --- | --- | --- | --- | --- | --- | --- | --- | --- |
| Typical dose is 100 micrograms | 100 micrograms | £8.06 | 30 | 1 | £0.27 | 1 | £0.27 | 365 | £98.06 |
| Starting dose 50 micrograms | 100 micrograms | £8.06 | 30 | 1 | £0.27 | 0.5 | £0.13 | 365 | £49.03 |
| Maximal dose of 300 micrograms | 100 micrograms | £8.06 | 30 | 1 | £0.27 | 3 | £0.81 | 365 | £294.19 |

6.2.3 Midodrine

| Staff Administering Fludrocortisone | Grade | Time involved | Cost per minute | Cost per session* | Number of sessions | Total staff costs * |
| --- | --- | --- | --- | --- | --- | --- |
| Staff Member 1 |  |  |  |  |  |  |
| Staff Member 2 |  |  |  |  |  |  |
| Total |  |  |  |  |  |  |

* These are calculated

6.2.4 Medications

| Dose in Protocol | Strength of tablet according to BNF | Cost per pack | Number of tablets per pack | Frequency per day | Cost per tablet £ | Number of tables used per dose | Total cost per day | Number of days | Total Costs* |
| --- | --- | --- | --- | --- | --- | --- | --- | --- | --- |
| The typical starting dose is 2.5mg |  |  |  |  |  |  |  |  |  |
| Maximum dose 10mg |  |  |  |  |  |  |  |  |  |

* These are calculated

6.3 Healthcare Resources

6.3.1 National Health Care Service

| Resource | Mean length of consultation in minutes | Number of appointments | Cost per minute | Cost per Consultation | Total staff cost* | Source /Notes |
| --- | --- | --- | --- | --- | --- | --- |
| GP at Practice | 9.22 |  | £4.23 | £39.00 |  | 42 weeks per year. 41.4 hours per week, including direct staff costs with qualifications PSSRU 2019/20 |
| GP on Phone | 4 |  | £3.88 | £15.520 |  | 44 weeks per year, 41.7 hours per week PSSRU 2019/20, including other costs per year |
| Nurse at GP practice | NA |  | £0.73 | £44 |  | 41.9 weeks per year 37.5 hours per week costs including qualifications PSSRU 2019/20,44 per hour |
| Nurse on the Phone | 6.56 |  | £1.19 | £7.80 |  | 42 Weeks per year, 37.5 hours per week |

* These are calculated

6.3.2 National Health Care Service (cont)

| Ambulance service | Unit costs FY19/20 | Number of visits | Total costs* | Source/ Cost |
| --- | --- | --- | --- | --- |
| Hear and treat or refer |  |  |  | National cost collection 2019/20 Report |
| See and treat or refer |  |  |  | National cost collection 2019/20 Report |
| See and treat and convey |  |  |  | National cost collection 2019/20 Report |
| Calls |  |  |  | National cost collection 2019/20 Report |

* These are calculated

| Accident and Emergency | Average unit cost | Number of consultations | Total costs* | Source/ notes |
| --- | --- | --- | --- | --- |
| Emergency Medicine, Category 3 Investigation with Category 1-3 Treatment Type 01 admitted |  |  |  | National reference costs 19/20 |
| Emergency Medicine, Category 3 Investigation with Category 1-3 Treatment Type 01 admitted |  |  |  | National reference costs 19/20 |
| Emergency Medicine, Category 2 Investigation with Category 4 Treatment Type 01 admitted |  |  |  | National reference costs 19/20 |
| Emergency Medicine, Category 2 Investigation with Category 4 Treatment Type 01 non admitted |  |  |  | National reference costs 19/20 |
| Emergency Medicine, Category 2 Investigation with Category 3 Treatment Type 01 admitted |  |  |  | National reference costs 19/20 |
| Emergency Medicine, Category 2 Investigation with Category 3 Treatment Type 01 non admitted |  |  |  |  |
| Emergency Medicine, Category 2 Investigation with Category 1 Treatment Type 01 admitted |  |  |  | National reference costs 19/20 |
| Emergency Medicine, No Investigation with No Significant Treatment Type 01 non admitted |  |  |  | National reference costs 19/20 |
| Emergency Medicine, No Investigation with No Significant Treatment Type 01 admitted |  |  |  | National reference costs 19/20 |
| Emergency Medicine, No Investigation with No Significant Treatment Type 01 unknown |  |  |  | National reference costs 19/20 |

* These are calculated

6.3.3 Private Service

| Private service | number of consultations | length of time per consultation | cost per consultation | Cost per minute of consultation | Total costs* | Source/ notes |
| --- | --- | --- | --- | --- | --- | --- |
| Private hospital |  |  |  |  |  |  |
| Physiotherapy |  |  |  |  |  |  |
| Spa |  |  |  |  |  |  |
| Acupuncture |  |  |  |  |  |  |

* These are calculated

6.3.4 Personal social service

| Type of service | number of time service used per month | Frequency | Cost of social services* | Total Costs* | Sources/ notes |
| --- | --- | --- | --- | --- | --- |
| Pensions savings credit |  |  |  |  |  |
| Pensions Guarantee credit |  |  |  |  |  |
| Attendance Allowance |  |  |  |  |  |
| Personal Independence Payment (Daily living) |  |  |  |  |  |
| Personal Independence Payment (Mobility |  |  |  |  |  |
| Employment and Support Allowance |  |  |  |  |  |
| Universal Credit |  |  |  |  |  |
| Carer’s Allowance |  |  |  |  |  |
| Other welfare Benefits |  |  |  |  |  |

6.3.5 Social workers and other community-based health services

| Type of service | Length of time per consultation | number of consultations | Cost per consultation | Total cost* | Sources/ Notes |
| --- | --- | --- | --- | --- | --- |

##### 6.4 Time and Travel Questionnaire

6.4.1 Most recent hospital appointment

| Mode of transport | Cost of fare | Distance Travelled | Time spent travelling to the appointment | Time spent at the appointment | Main Activity | Accompanied by the hospital | Main activity of the person who accompanied | Person who accompanied time spent travelling | Person who accompanied time spent at the appointment | How much did the person accompany pay for a ticket and fare | How miles did the person accompany travel |
| --- | --- | --- | --- | --- | --- | --- | --- | --- | --- | --- | --- |

6.4.2 Hospital admission

| Mode of transport | Cost of fare | Distance Travelled | Time spent travelling to the appointment | Time spent at the appointment | Main Activity | Accompanied by the hospital | Main activity of the person who accompanied | Person who accompanied time spent travelling | Person who accompanied time spent at the appointment | How much did the person accompany for a ticket and fare | How miles did the person accompany travel |
| --- | --- | --- | --- | --- | --- | --- | --- | --- | --- | --- | --- |

6.4. GP/Nurse / Walkin Centre

| Mode of transport | Cost of fare | Distance Travelled | Time spent travelling to the appointment | Time spent at the appointment | Main Activity | Accompanied by the hospital | Main activity of the person who accompanied | Person who accompanied time spent travelling | Person who accompanied time spent at the appointment | How much did the person accompany for a ticket and fare | How miles did the person accompany travel |
| --- | --- | --- | --- | --- | --- | --- | --- | --- | --- | --- | --- |
