## Supplementary tables for "Control, Fludrocortisone or Midodrine for the treatment of Orthostatic Hypotension (CONFORM-OH): Results from an internal pilot randomised controlled trial"

Table 1. Secondary outcomes. *Calculated as supine minus standing nadir

|  | **Outcome** | **Measurement** | **Timepoint** |
| --- | --- | --- | --- |
| Clinical effectiveness | Change in symptoms | Orthostatic Hypotension Questionnaire (OHQ) | 3 months  12 months |
|  | Daily function | Nottingham Extended Activities of Daily Living Scale (NEADL) questionnaire | 3 months  6 months  12 months |
|  | Quality of life | EQ-5D-5L questionnaire | 3 months  6 months  12 months |
|  | Falls and syncope | Monthly falls diary:   - Time to first fall - Number of falls - Number of fallers - Fall rate per person year - Fall related injuries - Number of syncopal events | Over 12 months |
|  | Blood pressure | Supine and standing BP measured according to usual clinical practice:   - Orthostatic systolic BP drop* - Orthostatic diastolic BP drop* - Standing nadir systolic BP - Standing nadir diastolic BP | 3 months  6 months  12 months |
| Safety | Side effects | Adverse event reporting | 12 months |
|  | Safety | - Serious adverse reactions and events - Hospitalisations - Deaths | 12 months |
| Cost effectiveness |  | - Health & social care use questionnaire - Time & travel questionnaire | 12 months |

Table 2: Screening and recruitment data by site

| Site | Months open | Number screened | Number eligible | % eligible (of screened) | Number recruited | % recruited (of eligible) |
| --- | --- | --- | --- | --- | --- | --- |
| Newcastle | 9 | 52 | 11 | 21% | 5 | 45% |
| Walsall | 7 | 79 | 21 | 27% | 6 | 29% |
| Norfolk and Norwich | 7 | 26 | 6 | 23% | 1 | 17% |
| Dumfries and Galloway | 7 | 10 | 4 | 40% | 1 | 25% |
| Lewisham and Greenwich | 6 | 16 | 3 | 19% | 0 | 0% |
| Gateshead | 5 | 3 | 0 | 0% | 0 | 0% |
| Devon and Exeter | 4 | 3 | 1 | 33% | 0 | 0% |
| Bath | 4 | 87 | 3 | 3% | 0 | 0% |
| Salford | 3 | 6 | 0 | 0% | 0 | 0% |
| **Total** | **52** | **282** | **49** | **17.4%** | **13** | **26.5%** |

Table 3: Screening data

| **Screened** | **N = 282** |
| --- | --- |
| **Eligibility status** |  |
| Eligible | 49 (17%) |
| Ineligible | 233 (83%) |
| **Reasons for ineligibility** | **N = 233** |
| Use of fludrocortisone or midodrine within the last six months | 120 (52%) |
| *Drug used* |  |
| Fludrocortisone | 45 (38%) |
| Midodrine | 64 (53%) |
| Both | 10 (8%) |
| Unknown | 1 (1%) |
| *Current or previous use* |  |
| Current | 116 (97%) |
| Previous | 3 (3%) |
| Not available | 1 (1%) |
| A known contra-indication to fludrocortisone or midodrine which outweighs the potential clinical benefit | 25 (11%) |
| *Drug contraindicated* |  |
| Midodrine | 19 (76%) |
| Both | 2 (8%) |
| Unknown | 4 (16%) |
| Inability to comply with the study procedures as specified in the schedule of events | 21 (9%) |
| Supine hypertension (where the risks of treatment outweigh the benefits) at baseline | 20 (9%) |
| Other: A score of <2 on the OHQ | 12 (5%) |
| Other: Too frail/pre-existing conditions | 7 (3%) |
| Other: No BP drop as per inclusion criteria | 6 (3%) |
| OH secondary to acute or reversible causes | 5 (2%) |
| Other: Unable to provide informed consent | 2 (1%) |
| Other: already started on other medication/ didn’t want to start new | 2 (1%) |
| Other: Unable to wait 4 weeks of lifestyle modification | 2 (1%) |
| Terminal illness or life expectancy <12 months | 1 (<0.5%) |
| Inability to communicate in English | 1 (<0.5%) |
| Other: Did not have OH on examination | 1 (<0.5%) |
| Other: Unknown reason | 3 (1%) |
| Eligibility status unknown | 5 (2%) |
| **Reasons for eligible patients not participating** | **N = 36** |
| Participant declined | 30 (83%) |
| I would find it difficult to participate (e.g. due to burden of assessments/questionnaires) | 10 (33%) |
| No reason given | 8 (27%) |
| I do not want to change my treatment plan | 5 (17%) |
| I see no benefit in the study to me | 2 (7%) |
| I do not want to be randomised to usual care | 2 (7%) |
| Other: Did not want to be randomised into either IMP | 2 (7%) |
| I am too busy | 1 (3%) |
| Participant relocated/unable to contact after screening | 3 (8%) |
| Patient not approached by doctor | 3 (8%) |

Table 4: Completion rates of outcome measures

|  | Conservative management | Conservative management plus fludrocortisone | Conservative management plus midodrine | Overall |
| --- | --- | --- | --- | --- |
| **Month 3^1^** | **Exp = 1** | **Exp = 4** | **Exp = 6** | **Exp = 11** |
| **BP^2^** | Exp = 2 | Exp = 4 | Exp = 6 | Exp = 12 |
| Collected | 1 (50%) | 4 (100%) | 6 (100%) | 11 (92%) |
| Not collected | 1 (50%) | 0 (0%) | 0 (0%) | 1 (8%) |
| ***How was the BP obtained?*** |  |  |  |  |
| Study visit | 0 (0%) | 4 (100%) | 6 (100%) | 10 (91%) |
| Other medical records | 1 (100%) | 0 (0%) | 0 (0%) | 1 (9%) |
| **OHQ** | 1 (100%) | 4 (100%) | 6 (100%) | 11 (100%) |
| **NEADL** | 1 (100%) | 4 (100%) | 6 (100%) | 11 (100%) |
| **Falls Diary** |  |  |  |  |
| Falls data collected | 0 (0%) | 4 (100%) | 6 (100%) | 10 (91%) |
| Falls diary completed | 0 (0%) | 2 (50%) | 5 (83%) | 7 (64%) |
| **Month 6^1^** | **Exp = 0** | **Exp = 4** | **Exp = 6** | **Exp = 10** |
| **BP^2^** | Exp = 2 | Exp = 4 | Exp = 6 | Exp = 12 |
| Collected | 0 (0%) | 4 (100%) | 6 (100%) | 10 (83%) |
| Not collected | 2 (100%) | 0 (0%) | 0 (0%) | 2 (17%) |
| ***How was the BP obtained?*** |  |  |  |  |
| Study visit | NA | 4 (100%) | 5 (83%) | 9 (90%) |
| Other medical records | NA | 0 (0%) | 1 (17%) | 1 (10%) |
| **OHQ** | NA | 4 (100%) | 6 (100%) | 10 (100%) |
| **NEADL** | NA | 4 (100%) | 6 (100%) | 10 (100%) |
| **Falls Diary** |  |  |  |  |
| Falls data collected | NA | 4 (100%) | 6 (100%) | 10 (100%) |
| Falls diary completed | NA | 2 (50%) | 4 (67%) | 6 (60%) |
| **Month 12^1^** | **Exp = 0** | **Exp = 4** | **Exp = 5** | **Exp = 9** |
| **BP^2^** | Exp = 2 | Exp = 4 | Exp = 5 | Exp = 11 |
| Collected | 0 (0%) | 4 (100%) | 5 (100%) | 9 (82%) |
| Not collected | 2 (100%) | 0 (0%) | 0 (0%) | 2 (18%) |
| ***How was the BP obtained?*** |  |  |  |  |
| Study visit | NA | 3 (75%) | 5 (100%) | 8 (89%) |
| Self-assessed at home | NA | 1 (25%) | 0 (0%) | 1 (11%) |
| **OHQ** | NA | 4 (100%) | 5 (100%) | 9 (100%) |
| **NEADL** | NA | 4 (100%) | 5 (100%) | 9 (100%) |
| **Falls Diary** |  |  |  |  |
| Falls data collected | NA | 4 (100%) | 5 (100%) | 9 (100%) |
| Falls diary completed | NA | 1 (25%) | 4 (80%) | 5 (56%) |

*^1^The number of expected CRFs at this visit (unless otherwise stated)*

*^2^Also expected for those that withdrew but allowed use of routinely collected medical data.*

Table 5: Summary of learning from feasibility components

| **Feasibility issues** | **Key findings & learning** |
| --- | --- |
| Permissions | Ethical permissions were delayed due to prioritisation of Covid-19 studies |
| Site opening | Clinical and research staff redeployed during and after COVID-19 |
| Eligible patient population | The biggest barrier to recruitment was the exclusion criteria, predominantly by excluding those already taking one of the trial interventions. Potential solutions include having a washout period before entering the trial, although this may not be acceptable to patients and clinicians and prior experience of treatment may introduce bias. |
| Acceptance rate | One third of eligible participants declined due to the burden of assessments. Given the level of frailty and multiple long-term conditions in this population this may be expected. Feedback from trial participants was that the number of assessments was not burdensome. |
| Intervention crossover | Given the low numbers of participants, the estimate is unlikely to be of a value. Participants and clinicians reported reluctance to be randomised to the conservative arm; had the trial continued this may have led to increased crossover from the control arm to one of the intervention arms. |
| Retention | Retention was 100% at the primary time point of six months for the intervention arms but was 0 of 3 for the control arm. Retention remained good (90%) for the intervention arms at 12 months. As described above, the control arm appears to have been a barrier to recruitment and retention, although numbers in all arms were small to make definitive conclusions. |
| Availability of outcome measures | With the exception of the falls diaries, completion rates of the outcomes was excellent. Of those remaining in the trial the primary outcome was available for all participants. Of secondary outcomes, with the exception of falls diaries, these were all available for all participants at all time points.  Return rates for falls diaries are known to be poor in falls studies. Strategies such as telephone and text reminders may help. One participant stated they would have preferred an online falls diary. In one participant who did not return their falls diary, a review of their medical records identified an injurious fall. Reviewing medical records will likely improve identification of injurious falls rather than relying on self-report. |

Table 6: Participant demographics and characteristics at baseline

|  | Conservative management | Conservative management plus fludrocortisone | Conservative management plus midodrine | Total |
| --- | --- | --- | --- | --- |
|  | (N = 3) | (N = 4) | (N = 6) | (N = 13) |
| **Age** |  |  |  |  |
| Mean (SD) | 75.3 (14.6) | 70.3 (12.0) | 75.0 (6.0) | 73.6 (9.6) |
| Median (Range) | 80 (59, 87) | 74 (53, 80) | 73 (70, 84) | 73 (53, 87) |
| <80 years | 1 (33%) | 3 (75%) | 4 (67%) | 8 (62%) |
| ≥80 | 2 (67%) | 1 (25%) | 2 (33%) | 5 (38%) |
| **Sex at birth** |  |  |  |  |
| Female | 2 (67%) | 1 (25%) | 4 (67%) | 7 (54%) |
| Male | 1 (33%) | 3 (75%) | 2 (33%) | 6 (46%) |
| **Ethnicity** |  |  |  |  |
| Any white background | 3 (100%) | 4 (100%) | 6 (100%) | 13 (100%) |
| **BMI** |  |  |  |  |
| No. with data | 3 (100%) | 3 (75%) | 6 (100%) | 12 (92%) |
| Mean (SD) | 22.5 (6.1) | 26.1 (2.5) | 23.8 (1.5) | 24.1 (3.3) |
| Median (Range) | 22.4 (16.4, 28.7) | 25.8 (23.7, 28.7) | 24.2 (21.7, 26.0) | 24.2 (16.4, 28.7) |
| **Disease aetiology** |  |  |  |  |
| Neurogenic OH | 1 (33%) | 1 (25%) | 3 (50%) | 5 (38%) |
| Non-neurogenic OH | 2 (67%) | 3 (75%) | 3 (50%) | 8 (62%) |
| **Medical history** |  |  |  |  |
| Diabetes | 0 (0%) | 0 (0%) | 0 (0%) | 0 (0%) |
| Pure Autonomic Failure | 0 (0%) | 0 (0%) | 0 (0%) | 0 (0%) |
| Parkinson's disease | 1 (33%) | 0 (0%) | 3 (50%) | 4 (31%) |
| **OHQ score** |  |  |  |  |
| Mean (SD) | 6.8 (1.7) | 6.8 (1.6) | 6.0 (1.6) | 6.4 (1.5) |
| Median (Range) | 7.5 (4.9, 8.0) | 6.7 (5.1, 8.8) | 5.9 (3.8, 8.7) | 6.2 (3.8, 8.8) |
| **NEADL score** |  |  |  |  |
| Mean (SD) | 13.7 (6.0) | 16.0 (9.4) | 14.2 (5.3) | 14.6 (6.4) |
| Median (Range) | 13 (8, 20) | 20 (2, 22) | 15 (6, 21) | 17 (2, 22) |
| **Blood pressure** |  |  |  |  |
| How was the BP obtained? |  |  |  |  |
| Study visit | 2 (67%) | 2 (50%) | 2 (33%) | 6 (46%) |
| Other medical records | 1 (33%) | 1 (25%) | 4 (67%) | 6 (46%) |
| Self-assessed at home | 0 (0%) | 1 (25%) | 0 (0%) | 1 (8%) |
| Supine Blood Pressure |  |  |  |  |
| Systolic |  |  |  |  |
| Mean (SD) | 134.7 (20.7) | 128.5 (26.2) | 141.3 (22.9) | 135.8 (22.3) |
| Median (Range) | 131 (116, 157) | 129 (96, 160) | 151 (102, 160) | 132 (96, 160) |
| Diastolic |  |  |  |  |
| Mean (SD) | 74.7 (4.0) | 70.5 (24.6) | 74.5 (10.6) | 73.3 (14.3) |
| Median (Range) | 77 (70, 77) | 69 (42, 102) | 79 (53, 80) | 77 (42, 102) |
| Lowest standing BP |  |  |  |  |
| Systolic |  |  |  |  |
| Mean (SD) | 88.3 (23.6) | 88.8 (10.6) | 105.0 (18.4) | 96.2 (18.3) |
| Median (Range) | 80 (70, 115) | 93 (73, 96) | 105 (80, 130) | 94 (70, 130) |
| Diastolic |  |  |  |  |
| Mean (SD) | 60.0 (20.9) | 61.5 (13.0) | 65.3 (8.4) | 62.9 (12.2) |
| Median (Range) | 50 (46, 84) | 62 (46, 77) | 66 (55, 76) | 60 (46, 84) |
| Postural blood pressure drop |  |  |  |  |
| Systolic |  |  |  |  |
| Mean (SD) | 46.3 (4.5) | 39.8 (19.6) | 36.3 (18.2) | 39.7 (15.9) |
| Median (Range) | 46 (42, 51) | 34 (23, 68) | 34.5 (11, 64) | 39 (11, 68) |
| Diastolic |  |  |  |  |
| Mean (SD) | 14.7 (19.6) | 9.0 (23.6) | 9.2 (10.4) | 10.4 (15.9) |
| Median (Range) | 20 (-7, 31) | -1 (-6, 44) | 7.5 (-6, 22) | 7 (-7, 44) |
| **Conservative measures advised at baseline** |  |  |  |  |
| Trigger avoidance | 2 (67%) | 3 (75%) | 5 (83%) | 10 (77%) |
| Hydration | 3 (100%) | 4 (100%) | 6 (100%) | 13 (100%) |
| Bolus water drinking | 1 (33%) | 2 (50%) | 3 (50%) | 6 (46%) |
| Salt intake | 3 (100%) | 2 (50%) | 2 (33%) | 7 (54%) |
| Caffeine intake | 1 (33%) | 0 (0%) | 1 (17%) | 2 (15%) |
| Physical counter-manoeuvres | 3 (100%) | 2 (50%) | 3 (50%) | 8 (62%) |
| Provision of compression stockings (below knee) | 0 (0%) | 1 (25%) | 0 (0%) | 1 (8%) |
| Provision of compression stockings (full length) | 0 (0%) | 0 (0%) | 0 (0%) | 0 (0%) |
| Abdominal compression | 0 (0%) | 0 (0%) | 0 (0%) | 0 (0%) |
| Medication review | 1 (33%) | 2 (50%) | 3 (50%) | 6 (46%) |
| Other - Advised to consider stockings from GP | 1 (33%) | 1 (25%) | 2 (33%) | 4 (31%) |

Figure 1: CONSORT flow diagram

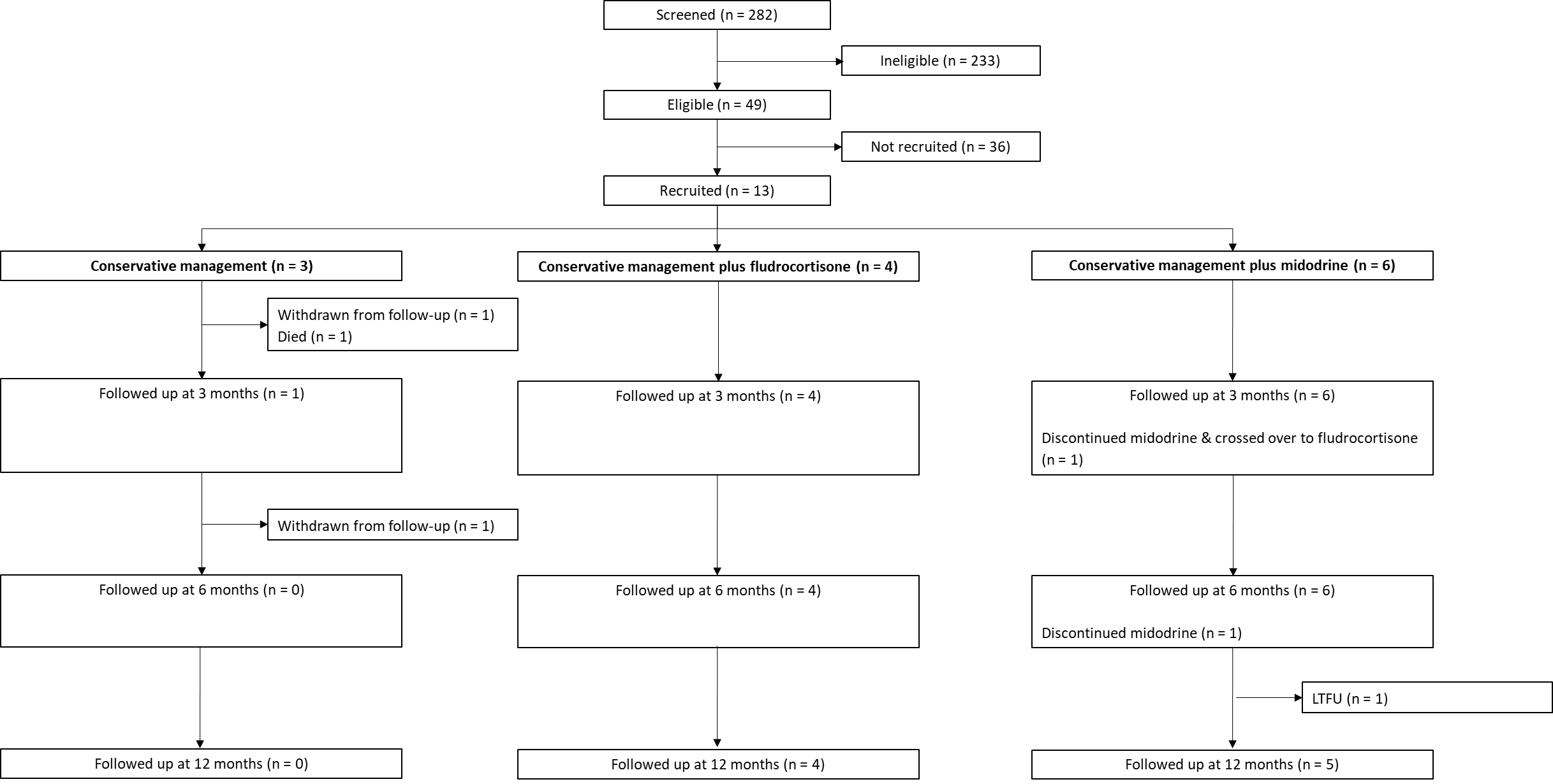

LTFU: Lost to follow-up
